# Modelling the cost-effectiveness of TasP and PrEP in female sex workers in Cotonou, Benin

**DOI:** 10.1101/2023.12.06.23299621

**Authors:** F Cianci, L Geidelberg, KM Mitchell, L Kessou, A Mboup, S Diabate, L Behanzin, FA Guedou, DM Zannou, N Geraldo, E Goma-Matsétsé, K Giguère, M Aza-Gnandji, M Diallo, R Kêkê, M Bachabi, K Dramane, C Lafrance, D Affolabi, MP Gagnon, F Gangbo, R Silhol, F Terris-Prestholt, MC Boily, M Alary, P Vickerman

## Abstract

**Introduction:** Treatment as prevention (TasP) and pre-exposure prophylaxis (PrEP) could impact the HIV epidemic among Female Sex Workers (FSW) but their cost-effectiveness is uncertain in this group. This study aims to model the cost-effectiveness of TasP and PrEP among FSW in Cotonou, Benin.

**Methods:** A demonstration study assessed TasP and PrEP use among FSW in Cotonou. A dynamic HIV transmission model was developed to estimate the impact of this intervention and published elsewhere. Incremental economic costs of the study were collected prospectively capturing both provider and FSW costs. The incremental cost-effectiveness ratio per HIV infection and disability-adjusted life years (DALY) averted were estimated over a 20 year time horizon with costs converted to USD 2020 and both costs and DALYs discounted at 4.5% per year. Different cost scenarios were modelled to investigate the cost-effectiveness of the intervention as delivered by the government reflecting current day implementation and resource costs.

**Results:** The mean provider annual economic cost per FSW on TasP was $646, with an initiation cost of $347 and mean annual user costs were $16. The mean initiation costs for PrEP were $268, mean annual provider costs were $359-$499 and annual user cost $15-$21 depending on adherence level. TasP was found to be cost saving for all cost scenarios examined compared to routine HIV care for FSW. PrEP was not cost-effective for any cost scenario, population coverage or adherence level examined.

**Conclusion:** Our results support TasP but not PrEP for FSW in our setting. A streamlined, outreach delivery model with reduced costs should be investigated to assess its cost-effectiveness in this setting.

## Introduction

Both treatment as prevention (TasP) and pre-exposure prophylaxis (PrEP) have emerged as safe and effective strategies for HIV prevention [1, 2]. Studies of TasP, the initiation of antiretroviral therapy (ART) immediately following a HIV diagnosis and irrespective of CD4 count, have consistently reported positive results with efficacy as high as 96% in reducing HIV transmission in serodiscordant heterosexual couples [1] and zero transmission in gay serodiscordant couple[3, 4]. Studies of oral PrEP have examined the use of tenofovir disoproxil fumarate (TDF) or TDF and emtricitabine (FTC) use in various groups and settings with mixed results ranging from 86% effectiveness in men who have sex with men (MSM) in high income countries [5, 6] to 62% in heterosexual adults in Botswana and 49% in persons who inject drugs (PWID) in Thailand [7, 8]. Studies of PrEP use in women have shown more heterogeneous results with some studies showing effectiveness in women in African countries [2, 7] and others finding none [9, 10], mainly due to poor adherence. There are concerns that lower PrEP adherence levels have greater consequences in terms of efficacy in women [11] when compared to MSM probably due to the lower concentration of drug levels found in vaginal tissue than rectal tissue after TDF administration [12]. The important link between effectiveness and adherence highlights the potential difficulty in targeting more marginalised populations that may find it harder to achieve high adherence, including female sex workers (FSW) [13, 14].

A number of combined HIV prevention programmes providing ART and health promotion and education activities for FSW in low- and middle-income countries have shown good results in terms of reducing risk behaviour, HIV/STI prevalence and cost-effectiveness [15-22]. However, the TasP/PrEP demonstration projects conducted in South Africa and Benin in 2014-2016were the first to examine the specific impact of TasP and PrEP among FSW [23, 24].

In Benin, as in many West African countries, the HIV epidemic remains concentrated in key populations, with an estimated HIV prevalence amongst FSW in 2015 of 16% compared to 1% in the general population [25]. More recent studies show a reduction in HIV prevalence to 8% in FSW and 0.9% in the general population [26, 27].

A previous modelling analysis estimated that the Benin TasP/PrEP study for FSWs prevented 8% (95% uncertainty interval (UI): 6–12) and 7% (95% UI: 3–11) of HIV infections among FSW and the whole population, respectively, in Cotonou over its 2 years of implementation [24]. The study found that TasP had greater impact on HIV infections averted than PrEP, due to greater observed adherence and retention to TasP among FSW infected with HIV than FSW on PrEP. This study aims to estimate the cost-effectiveness of the TasP/PrEP demonstration study among FSW in Benin using these model-based impact estimates [24]. We also aimed to determine the cost-effectiveness of a less intensive intervention as would be delivered by the Ministry of Health of Benin, both at the time of the intervention and using current day drug costs.

## Methods

### The setting

The TasP/PrEP demonstration project aimed to assess the feasibility and usefulness of integrating TasP/PrEP into the combination prevention package offered to professional FSW in Benin. FSW in Cotonou, the economic centre of Benin, were recruited in the field by outreach workers and invited to attend a dedicated FSW clinic run from the local STI clinic (DIST) where they were screened for HIV, syphilis, gonorrhoea, chlamydia and hepatitis B. FSW willing to participate in the study were recruited to either the TasP or PrEP arm of the study according to their HIV status. Screening began in September 2014 and was completed in December 2015. Follow-up ended on December 31, 2016. Women were reviewed at the clinic every three months during the study period to assess clinical status and adherence. Further details and results have been published previously [28].

### Participant details

A total of 105 women were recruited to the TasP arm of the study (median age 35 years) and 256 to the PrEP arm (median age 31 years). Follow-up varied from 12 to 24 months and overall retention rate for TasP was 59%, and 47% for PrEP [28]. Optimal adherence to PrEP, measured through tenofovir plasma concentration above 35 ng/mL, combining all study visits was 37% [29]. There were two seroconversions during the study period.

### Model structure, parameterisation and analysis

We developed a dynamic compartmental model of sexual HIV transmission representing the open and growing heterosexual adult (aged 15-59 years old) population of Cotonou including professional and part-time FSW, their clients, and the male and female low-risk population. A description of the model was previously published [24] (Supplementary Material (SM) Figure S1 and S2).

The model reflects HIV disease progression through four stages characterised by decreasing CD4 cell count levels, as well as the HIV treatment cascade including testing, PrEP (professional FSW only), ART, and ART dropout. FSW can initiate PrEP into either perfectly-, intermediate- or non-adherent categories, reflecting >40, 0.31-40 and <0.31 ng/ml blood plasma tenofovir concentration, respectively. We assumed that perfect, intermediate and non-adherent categories represented 90-95%, 30-49% and 0% reductions in per sex-act transmission probability, respectively [24]. Individuals on ART have reduced disease progression and HIV-related mortality compared to those not on treatment, while those who have ceased ART have HIV progression and mortality equivalent to ART-naïve individuals. Those who drop out of ART can re-initiate ART. HIV-negative individuals can become infected with HIV at a per-capita force of infection rate, which depends on the annual number of commercial and non-commercial partnerships, number of sex acts per partnership, levels of condom use, PrEP use, and HIV prevalence, disease stage (e.g. primary or AIDS stage) and ART status in their partners. The model structure and equations are shown in the Supplementary material.

Fixed estimates and prior ranges were defined for parameters from a variety of sources in Cotonou, including Benin census data [30-33], population size estimate studies [34, 35], integrated bio-behavioural surveys among FSW and clients [15, 25, 35-37], household surveys [38, 39] government reports [40] and HIV natural history studies [41-50]. Within a Bayesian framework, the model was calibrated to the TasP/PrEP demonstration study data on uptake, adherence, retention and coverage for ART and PrEP (2014-2017) [28], as well as HIV prevalence (1993-2017) and ART coverage (2000-2017) by risk group [24]. Key parameter values and sources are available in Supplementary material Table S1A-1F and S2.

Model calibration resulted in 111 posterior parameter sets out of 79 million which well captured available HIV epidemiological and intervention data by risk groups in Cotonou, as reported in a previous publication [24] (Supplementary materials Section 6-8). The TasP/PrEP study intervention scenario was simulated to reflect the uptake, adherence and retention of FSW to PrEP and TasP observed during the 2-year study [24, 28], and various other long-term PrEP and TasP intervention scenarios, as well as a counterfactual scenario which assumed no PrEP or TasP for FSW but with government provided ART (i.e HIV+ professional FSW who had a CD4 cell count < 500 cells/ml blood) [24, 28] (Supplementary material Section 9). For all scenarios, incident HIV infections were counted in the intervention and counterfactual. Disability-adjusted life-years (DALYs) were also derived by counting the number of individuals in each risk group and HIV stage at the mid-point of the year (to estimate life-years), and then multiplying these by the mean DALY weight [51] for each stage of HIV infection. Epidemiological impact outcomes were calculated over 20 years by comparing incident HIV infections and DALYs in the intervention and counterfactual scenarios, to generate infections averted and DALYs averted respectively.

### Cost analysis

An incremental cost analysis was conducted from a societal perspective. Data on costs incurred by the FSW to attend the clinic were collected prospectively at each visit through interview including direct transport and food costs incurred to attend the visit and indirect costs due to loss of earnings, as well as direct and indirect costs of other medical visits incurred between study visits.

To estimate the health provider’s incremental costs, data on cost and resources used to deliver TasP/PrEP intervention were collected prospectively. A micro-costing approach was used and research costs were excluded where possible. Data on capital costs such as buildings and equipment, start-up costs such as staff training and recurrent costs such as medications, laboratory tests, staff salaries and overhead costs were collected from project records. (Supplementary materials Table S4A and 4B). Costs were collected in West African Francs (FCFA), and inflated to USD2020 using Benin’s consumer price indices [52]. Capital costs were annualised over expected life span using Central Bank West African States (BCEAO) Benin’s Central Bank discount rate (4.5%[53]). Start-up costs were annualised over two years.

Data on numbers of visits and patients seen at the DIST were collected from clinic records to inform fixed cost allocation rules. Data on resources used such as medications and laboratory tests used during each visit were collected from individual case report forms (CRF) completed for each woman’s visit. Estimates of staff time spent on intervention related activities were collected through four different methods. First, a timesheet was completed for each FSW visit detailing the start and end time of interactions with each different staff member. A detailed timesheet was also completed by each staff member two days per month with start and end time of each activity during and between patient visits. Two time in motion studies were also conducted at the start and end of the study period. Lastly, staff were asked to estimate proportion of time spent on clinical and administrative duties related to the intervention. These methods were used to estimate a mean, minimum and maximum time spent by each staff member on specific tasks both during and between visits. Allocation rules for joint costs were developed through staff interviews and clinic utilization data.

The total economic cost for the intervention was calculated. A fixed cost per visit was calculated and included capital costs, start-up costs, total screening visit costs of women who were not enrolled into the study, utilities and staff time spent on administrative tasks. The cost of four outreach community workers was also included in this fixed cost; given the group nature of their work it was not possible to include them in the micro-costing. The variable cost for each visit for each woman was calculated by applying unit costs to all resources used during each visit. Variable costs were then added to fixed costs to estimate the cost of each visit type (Month 3 (M3), Month 6 (M6) etc) for each woman. The base case mean, base case minimum (participant with lowest costs) and base case maximum (participant with highest costs) cost of initiation on treatment (Screening visit+ recruitment visit+ day 14 visit) and of the annual cost of care (M3+M6+M9+M12) for full attendance were then calculated. The mean proportion of visits attended by the women in the TasP arm of the study was estimated and applied to the full annual cost of care to estimate mean annual cost of care. In the PrEP arm, women were stratified according to low/medium/high adherence to PrEP based on blood drug levels and the proportion of visits attended in each group was then applied to the mean annual cost of care.

### Ministry of health scenario

To estimate the cost of the intervention to the Ministry of Health (MOH) in Benin, costs and procedures were altered to reflect a programme that would be implemented by the government (Supplementary materials Table S4A). Capital costs were kept the same. Staff salaries were replaced with public sector salaries. Some of the support staff were removed and staff time spent on administrative tasks was reduced to reflect the reduced visit schedule. In this scenario, women in the TasP arm of the study were seen every six months instead of quarterly. In the PrEP arm, women were still seen quarterly for a rapid HIV test and medication dispensation. Patient level costs were also reduced to reflect the reduced number of visits attended.

### Cost-effectiveness analysis

For each scenario, the number of initiations and person-years on PrEP (only professional FSW) and ART (all groups) were counted annually. Annual initiations and person-years were multiplied by their estimated mean as well as minimum and maximum initiation and annual costs. The annual costs for each scenario were summed over 20 years from 2015 to 2035. Both costs and impact outcomes were discounted annually at using Benin’s Central bank inflation rate (4.5%) [53], and 10% in sensitivity analysis.

To estimate the cost-effectiveness of the PrEP/TasP study, epidemiological and cost outcomes were compared to those of the counterfactual scenario (no PrEP or TasP but standard care in FSW). Further impact scenarios examined different coverage levels achieved. Therefore, long term TasP intervention with 80% or 87% of HIV-infected FSW on ART were compared with the counterfactual scenario of standard ART care for HIV positive FSW. TasP was introduced as National policy in Benin in 2017 and we aimed to determine the cost-effectiveness of adding PrEP to this by simulating interventions of PrEP at different coverage levels combined with TasP (80% coverage), and compared it to TasP alone (80% coverage); this estimated the impact of long-term PrEP incremental to long-term TasP.

Cost-effectiveness was calculated as the mean (minimum and maximum) cost (USD) per infection or DALY averted (incremental cost-effectiveness ratio or ICER). We compared the cost-effectiveness of interventions against willingness-to-pay thresholds of $108 [54] and $1219 per DALY averted (1x GDP of Benin [55]).

### Sensitivity analysis

Sensitivity analyses were performed to test the robustness of our findings by exploring the effects of changes in key parameters on the ICER for TasP and PrEP. A worst case cost scenario was developed where all cost and resource inputs in the cost model with a degree of uncertainty were changed to their maximum possible value (e.g. highest staff salary within a range, maximum time spent on administrative task, minimum life span of capital inputs, joint cost allocation rules increased by 20%, etc). Details can be found in Supplementary materials Table S4C. Similarly, a best case scenario was also developed reducing all costs and inputs to the minimum value of their estimated range (Supplementary Materials Table S4C).

Drug costs decreased considerably since 2016, therefore, further sensitivity analyses were undertaken to evaluate the cost-effectiveness of PrEP and TasP if we assumed current drug and laboratory costs of implementing TasP and PrEP rather than the higher study costs. The scaled back intervention as described for the Ministry of Health scenario was used for this purpose.

#### Cost scenario A

Drug costs were reduced from $17 per month to $5 for PrEP and from $17 per month to $9 for TasP (EFV+3TC+TDF) to reflect change in drug costs between 2016 and 2021.

#### Cost scenario B

Unit laboratory investigation costs were noted to be considerably higher than in other published studies. Therefore, laboratory investigation costs were reduced by 50%, alongside drug cost reductions detailed above to reflect likely costs in 2021.

### Ethical Approval

The study protocol received approval from the ethics committee of the CHU de Québec– Université Laval, Québec, Canada, and the Benin National Ethics Committee for Health Research. All participants provided written consent in a free and informed manner and could decline or withdraw from the study at any time.

## RESULTS

The mean initiation cost per woman on TasP was $347, the mean provider annual economic cost was $646 and mean annual patient cost per woman $16 (Table 1). The initiation costs per woman on PrEP were lower at $268, the mean provider annual economic cost was $359-499 per woman depending on adherence levels to PrEP. The best and worst case scenario costs are also shown in Table 1 as minimum and maximum costs.

**Table 1.**
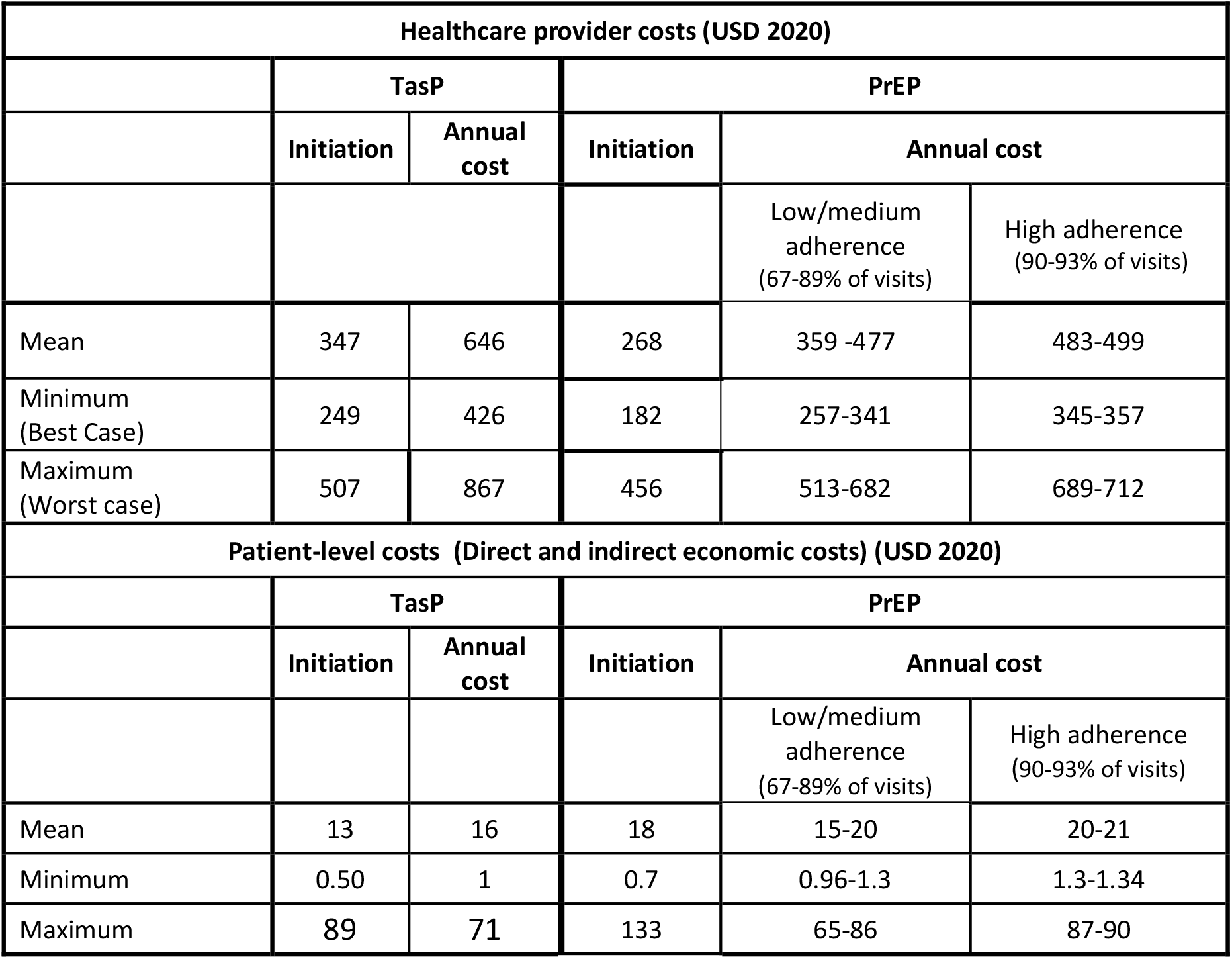
Mean initiation and annual cost per woman on TasP and PrEP in Benin, (USD 2020)

The fixed cost per visit (i.e. total fixed costs divided by total number of visits for both TasP and PrEP) was high at $57 mainly due to the high costs of non-clinical staff time, which accounted for 26% and 35% of total economic costs for TasP and PrEP respectively. The other main cost drivers were drug costs which accounted for 30% of total economic costs and laboratory costs which were 30% and 16% of total costs for the TasP and PrEP groups, respectively (Table 2).

**Table 2.**
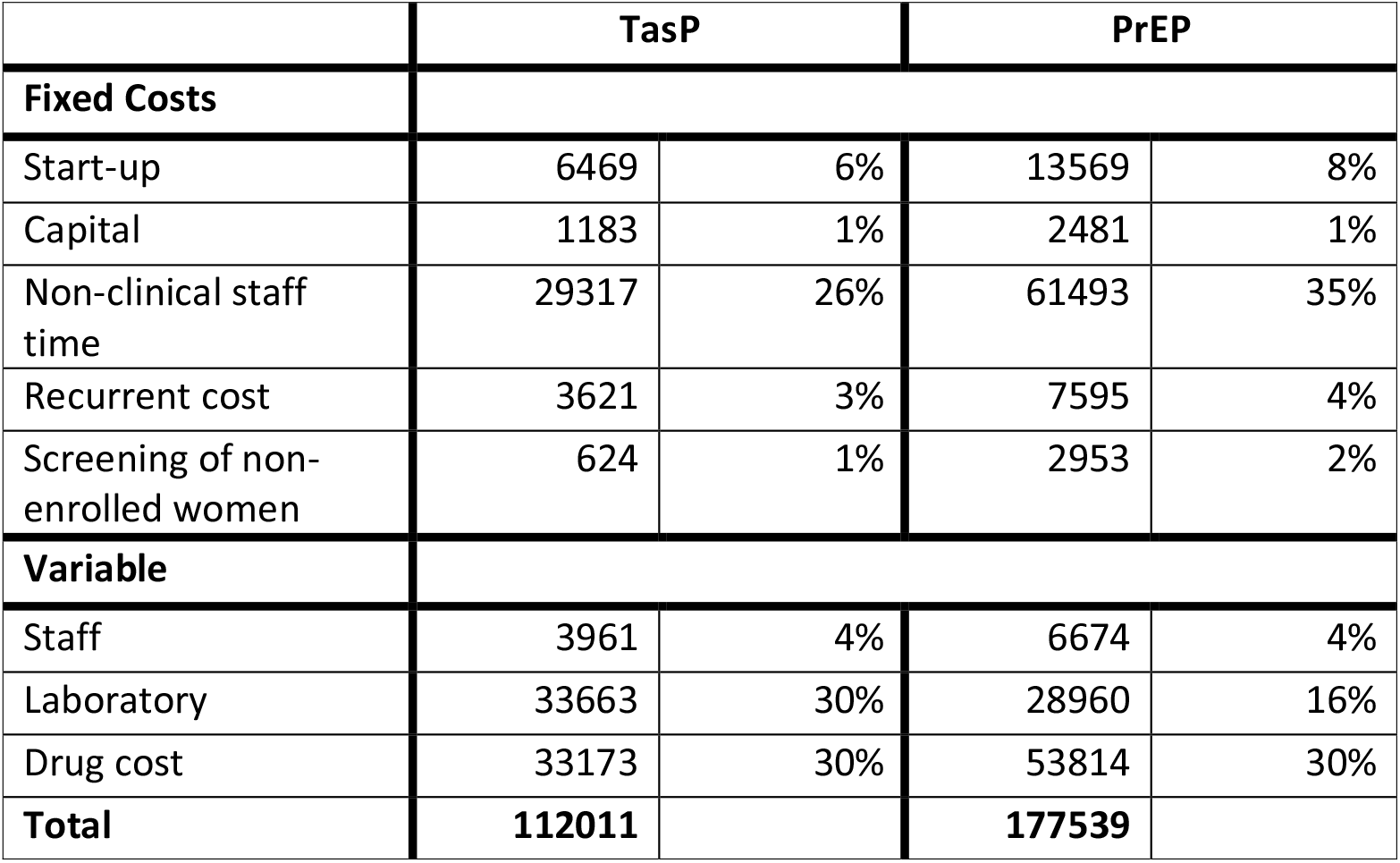
Breakdown of total economic costs for TasP/PrEP intervention over 2 years (USD 2020)

When the MOH scenario was considered, the initiation and mean annual costs per woman on TasP were reduced by about 37% to $220 and $415 respectively (Table 3). This cost reduction was mainly driven by reduced fixed visit cost and laboratory investigations. The mean annual cost for PrEP decreased less, by 18% to between $295-392, reflecting the continued quarterly visits and therefore the higher fixed costs.

**Table 3.**
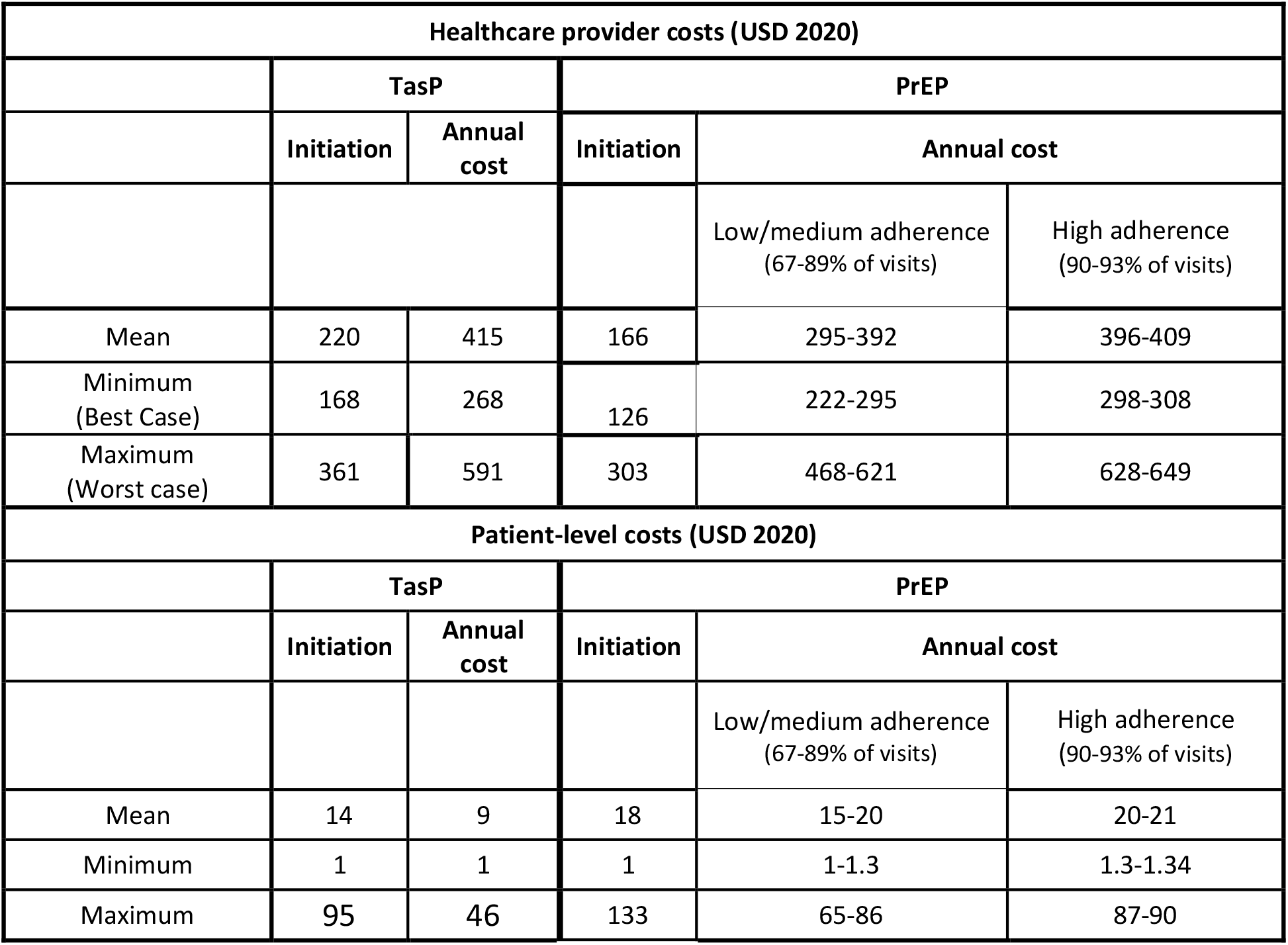
Ministry of Health scenario costs for mean initiation annual cost per woman on TasP and PrEP in Benin (USD 2020).

Details of the modelled impact estimates used in the cost-effectiveness analysis are reported elsewhere [24] and summary figures can be found in Supplementary Material Figure S14-16.

Figure 1 presents the absolute costs and the costs per HIV infection and DALY averted (over 20 years) of TasP and PrEP given over the two year study period assuming different cost scenarios when compared to standard ART care for FSW. TasP is cost saving in every cost scenario ranging from savings of $141,828 (95% UI $187,815-$77,690) using base case 2016 drug costs to $169,490 (95%UI $126,219-$188,390) using scenario B 2021 drug costs and reduced laboratory costs.

**Figure 1.**
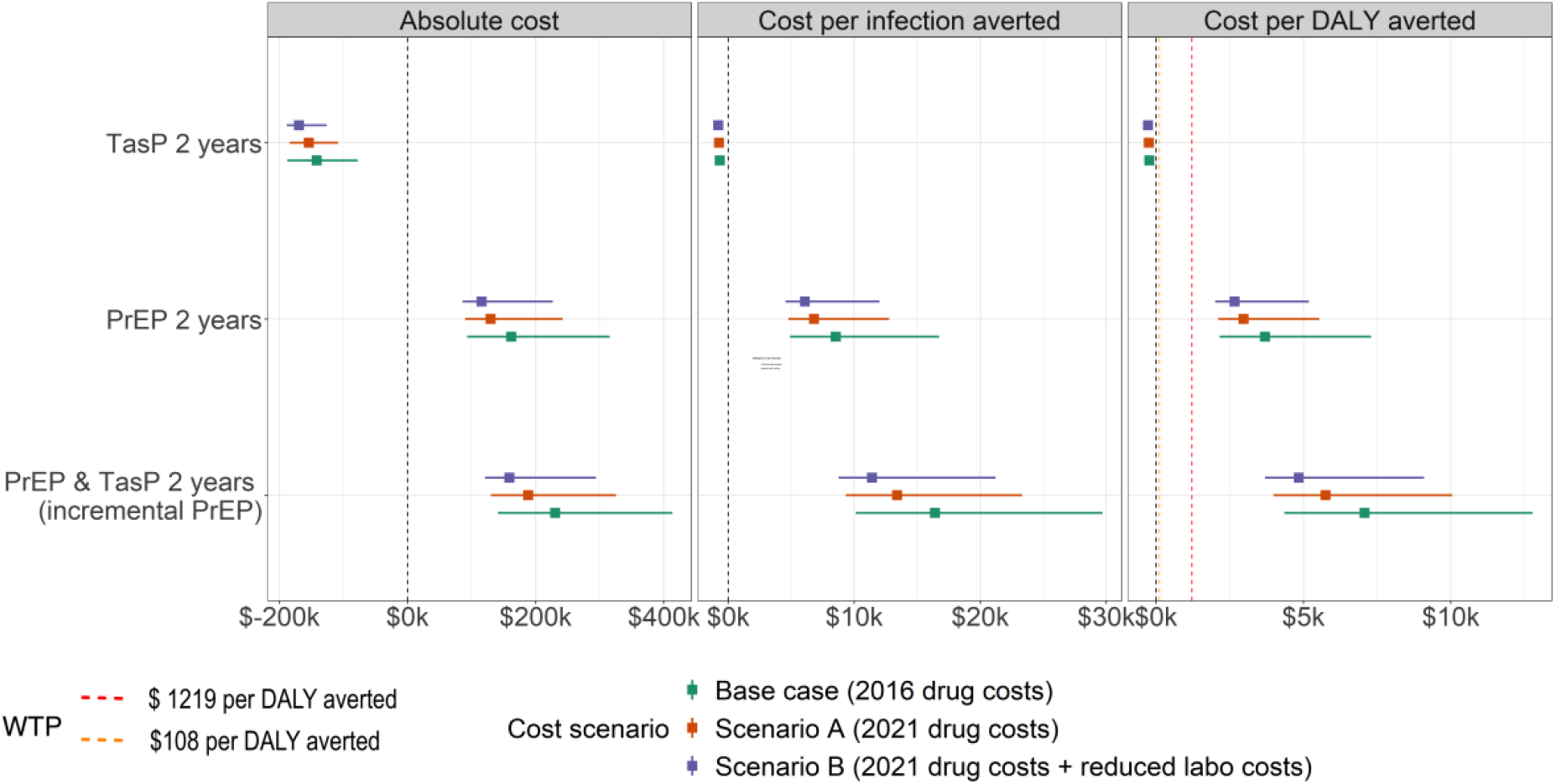
Cost and cost-effectiveness of two year TasP/PrEP as delivered by the study for 2 years when compared to standard ART care for HIV-positive FSW (USD 2020).

In contrast, PrEP was not cost-effective with respect to either of the cost-effectiveness willingness-to-pay thresholds examined, when compared to no PrEP for any cost scenario. The ICERs range from $3,697 (95%UI $2,156, $7,297) per DALY averted using 2016 base case drug costs to $2,975 (95%UI $2,107,$5,539) per DALY averted with current drug costs in Scenario A. When compared to two years of TasP, the incremental cost-effectiveness of PrEP is even less cost-effective at $4,844 (95%UI $3,700-$9,094) per DALY averted using Scenario B 2021 drug costs and reduced laboratory costs.

Figure 2 shows that TasP becomes more cost saving for higher coverage levels, with savings of $282,012 (95%UI $132,207-$363,084) over 20 years at 80% coverage of TasP among HIV-positive FSW and $422,875 (95%UI $265,939-$524,840) if 87% coverage is achieved, when using base case drug costs in 2016. Interestingly, TasP is more cost saving using base case costs from 2016 than when assuming current cheaper drug and laboratory costs in Scenario A and B. This is because the ART costs are reduced, and so the saving achieved for each HIV infection averted becomes less.

**Figure 2.**
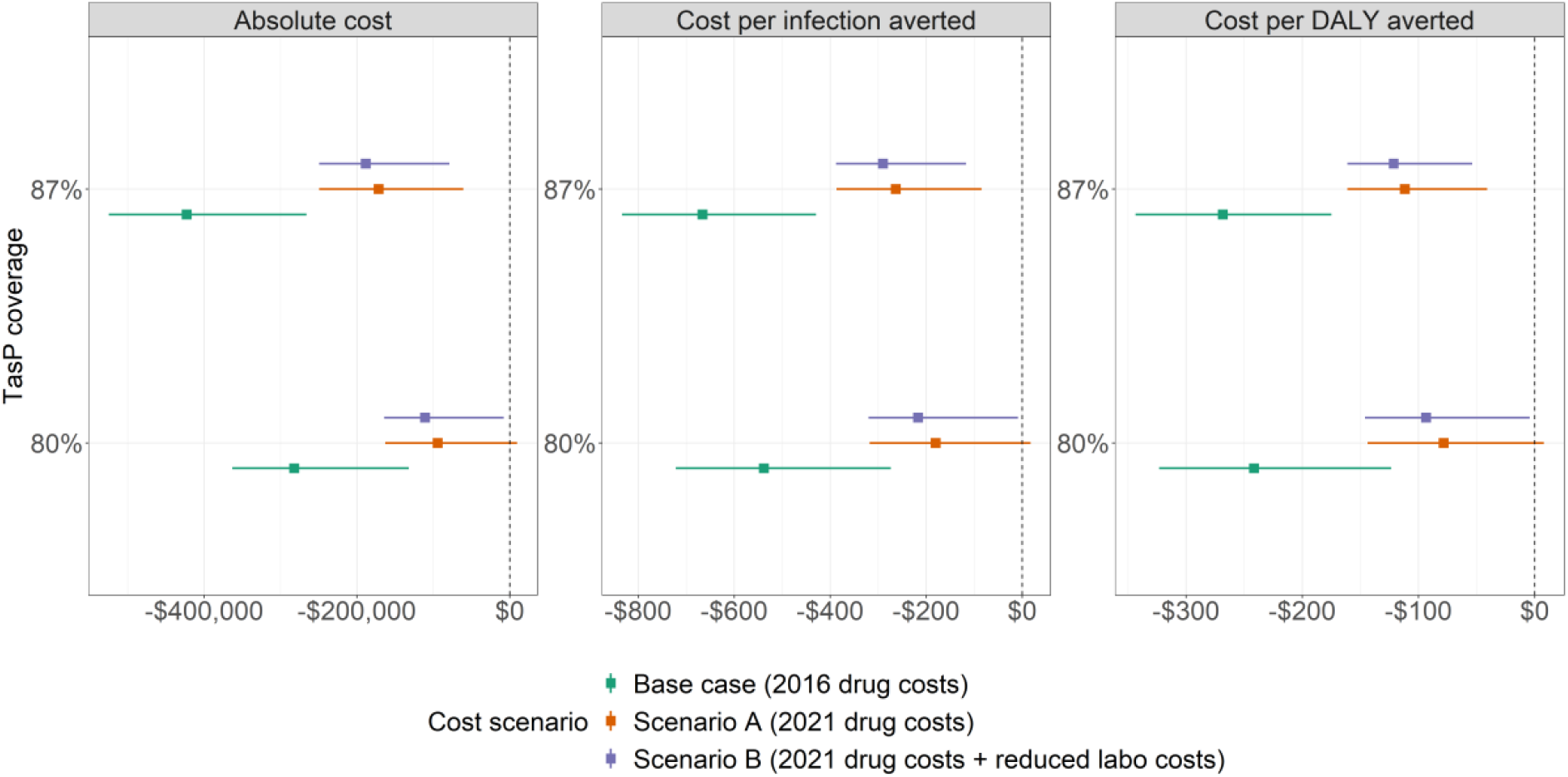
Cost and cost-effectiveness of TasP at 80% and 87% coverage among HIV-positive FSW as delivered by Ministry of Health over 20 years when compared to standard ART care in HIV positive FSW (USD 2020).

Figure 3 examines the cost-effectiveness of different population coverages of PrEP incremental to TasP alone at 80% of HIV positive FSW over 20 years. Again, PrEP is not cost-effective in any cost scenario examined.

**Figure 3.**
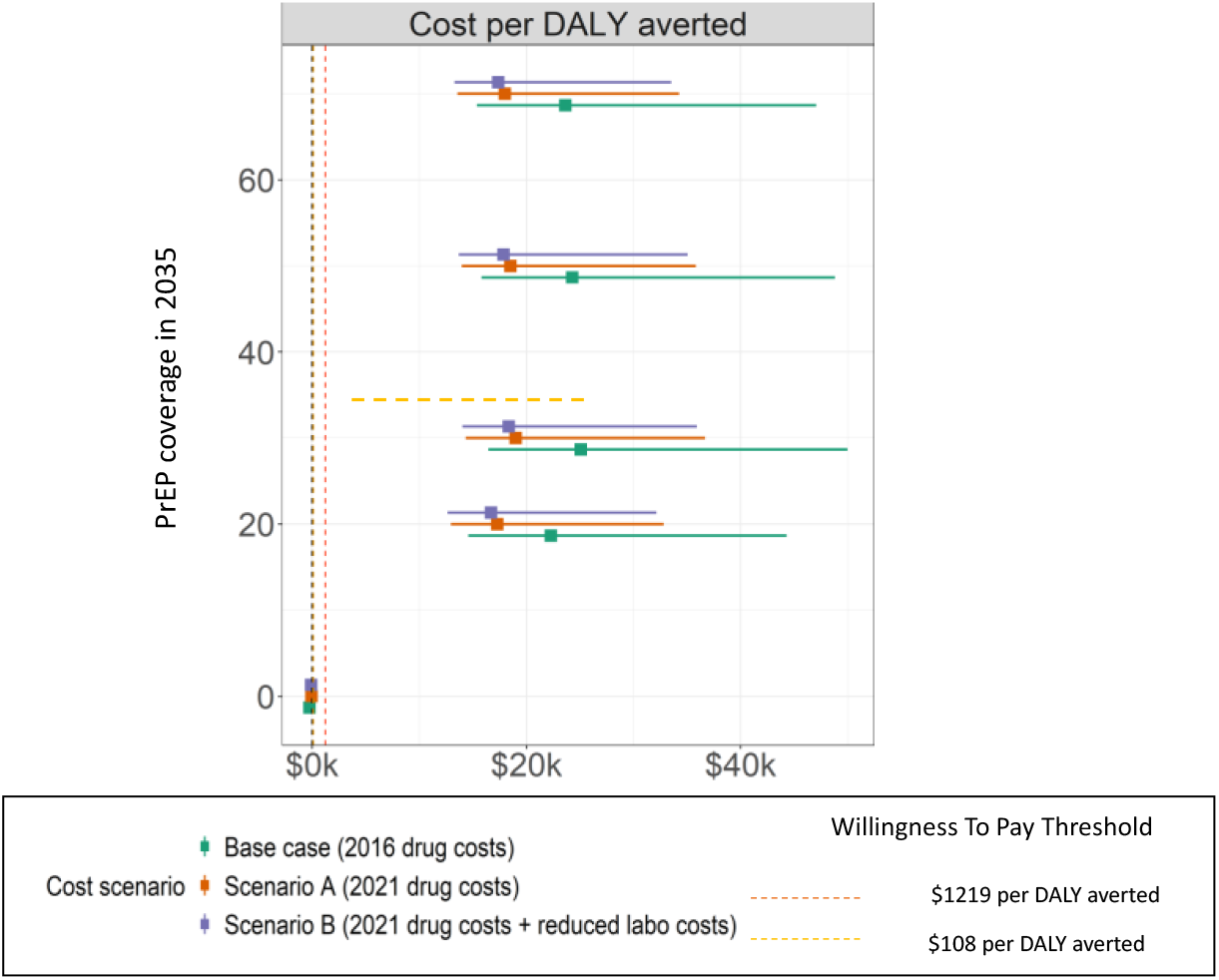
Incremental cost and cost-effectiveness of delivering PrEP at different coverage levels among HIV-negative FSW by Ministry of Health over 20 years compared to 80% coverage of TasP among HIV + positive FSW alone (USD2020)

## Discussion

Our cost-effectiveness analysis confirmed that TasP provision to FSW is a cost saving intervention in the Beninese context. This is because scaling up ART in FSWs reduces their HIV-related morbidity and also prevents new infections, so reducing the costs of future ART in FSWs, their clients and the general population. Interestingly, more is saved if the costs of ART are higher. In contrast, PrEP among FSWs was not cost-effective at any modelled coverage or for any realistic reductions in PrEP or ART costs when compared to standard care. A number of reasons explain these results. Firstly, TasP has immediate health benefits on the morbidity of individuals, whereas PrEP does not, and due to the higher coverage, adherence and retention levels achieved with ART a greater number of infections are averted compared to PrEP. Secondly, HIV incidence is relatively low among FSWs in Cotonou, and so the potential impact and cost-effectiveness of PrEP are lower than they could be for a higher incidence setting[56]. Lastly, our intervention costs were high due to both a resource intensive intervention and high unit costs. However, PrEP was still not cost-effective when cheaper costs and less visits were assumed. PrEP was even less cost effective when compared to TasP alone. In this scenario, where higher ART coverage results in even lower HIV incidence, the impact of any additional preventive measure is reduced and so PrEP becomes less cost-effective.

### Comparison to other studies

There is a paucity of published data on the cost-effectiveness of TasP and PrEP use in FSW. Jamieson et al, found a cost of $858 per HIV infection averted by delivering PrEP to FSW in South Africa, a country with a much higher prevalence and incidence of HIV than in Benin [57]. The unit costs of providing PrEP in that study were based on previously published studies and were much lower than those estimated in our study at $134 per FSW in the first year. This is because unit costs for all laboratory tests and drugs ($3.85) were much lower than those in our study ($17). A number of studies have reported costs of PrEP provision but did not estimate cost-effectiveness. Eakle et al found annual costs of PrEP delivery in South Africa in 2016 of $126 and $406 for TasP per person year, using data from a demonstration project among FSW [23]. Again, these lower costs are mainly due to much lower drug and test costs. In Zimbabwe in 2018, Mangenah et al, used real costs to estimate the cost of PrEP initiation for key populations including adolescent girls at $238 per person initiated which is similar to our study costs[58]. One other study in Kenya in 2014 found the annual cost of PrEP provision to a FSW to be $602. Their study assumed eleven yearly visits which is nearly double the number of visits as our study[59].

### Strengths and limitations

The main strength of our study is that it uses detailed programmatic costs and outcomes collected prospectively in the field to inform the cost-effectiveness analysis. We were also able to develop a plausible cost model to reflect intervention delivery by the Ministry of Health for both TasP and PrEP, making our results more applicable and relevant to a real world setting.

However, the impact projections were based on modelling, and as such, is based on assumptions that have uncertainty. To mitigate this issue, we incorporated known uncertainties in our analysis and conducted a best and worst case sensitivity analysis to test our cost assumptions, which did not significantly affect our results. Another limitation is the time lapsed since the study data collection. During this time, unit costs have reduced and the knowledge surrounding real life delivery and efficacy of PrEP in vulnerable groups has evolved. Our scenario analysis examined the effects of possible reductions in drug and laboratory costs to reflect more recent prices and given that these were two of the main cost drivers, it is likely that our modified costs are a close reflection of what the annual cost of care could be currently. However, even with these price reductions and assuming a simplified MOH delivery model, the provision of PrEP to FSW in this setting is unlikely to be cost-effective.

## Conclusion

Our results further support the roll out of TasP among FSW in Benin but raise important questions regarding the delivery of PrEP among FSW where incidence is low due to other successful interventions, even when important cost drivers like drugs and laboratory tests are reduced to levels seen in other African countries. For PrEP to be cost-effective in such settings, an outreach delivery model resulting in lower fixed costs combined with greatly reduced drug and laboratory costs should be considered.

## Data Availability

All data produced in the present study are available upon reasonable request to the authors

## References

1. Cohen MS, Chen YQ, McCauley M, Gamble T, Hosseinipour MC, Kumarasamy N, et al. Antiretroviral Therapy for the Prevention of HIV-1 Transmission. N Engl J Med. 2016;375(9):830-9. Epub 2016/07/19. doi: 10.1056/NEJMoa1600693. PubMed PMID: 27424812; PubMed Central PMCID: PMCPMC5049503.

2. Baeten JM, Donnell D, Ndase P, Mugo NR, Campbell JD, Wangisi J, et al. Antiretroviral prophylaxis for HIV prevention in heterosexual men and women. N Engl J Med. 2012;367(5):399-410. Epub 2012/07/13. doi: 10.1056/NEJMoa1108524. PubMed PMID: 22784037; PubMed Central PMCID: PMCPMC3770474.

3. Rodger AJ, Cambiano V, Bruun T, Vernazza P, Collins S, Degen O, et al. Risk of HIV transmission through condomless sex in serodifferent gay couples with the HIV-positive partner taking suppressive antiretroviral therapy (PARTNER): final results of a multicentre, prospective, observational study. Lancet. 2019;393(10189):2428-38. Epub 2019/05/06. doi: 10.1016/s0140-6736(19)30418-0. PubMed PMID: 31056293; PubMed Central PMCID: PMCPMC6584382.

4. Bavinton BR, Pinto AN, Phanuphak N, Grinsztejn B, Prestage GP, Zablotska-Manos IB, et al. Viral suppression and HIV transmission in serodiscordant male couples: an international, prospective, observational, cohort study. Lancet HIV. 2018;5(8):e438-e47. Epub 2018/07/22. doi: 10.1016/s2352-3018(18)30132-2. PubMed PMID: 30025681.

5. Molina JM, Capitant C, Spire B, Pialoux G, Cotte L, Charreau I, et al. On-Demand Preexposure Prophylaxis in Men at High Risk for HIV-1 Infection. N Engl J Med. 2015;373(23):2237-46. Epub 2015/12/02. doi: 10.1056/NEJMoa1506273. PubMed PMID: 26624850.

6. McCormack S, Dunn DT, Desai M, Dolling DI, Gafos M, Gilson R, et al. Preexposure prophylaxis to prevent the acquisition of HIV-1 infection (PROUD): effectiveness results from the pilot phase of a pragmatic open-label randomised trial. Lancet. 2016;387(10013):53-60. Epub 2015/09/14. doi: 10.1016/s0140-6736(15)00056-2. PubMed PMID: 26364263; PubMed Central PMCID: PMCPMC4700047.

7. Thigpen MC, Kebaabetswe PM, Paxton LA, Smith DK, Rose CE, Segolodi TM, et al. Antiretroviral preexposure prophylaxis for heterosexual HIV transmission in Botswana. N Engl J Med. 2012;367(5):423-34. Epub 2012/07/13. doi: 10.1056/NEJMoa1110711. PubMed PMID: 22784038.

8. Choopanya K, Martin M, Suntharasamai P, Sangkum U, Mock PA, Leethochawalit M, et al. Antiretroviral prophylaxis for HIV infection in injecting drug users in Bangkok, Thailand (the Bangkok Tenofovir Study): a randomised, double-blind, placebo-controlled phase 3 trial. Lancet. 2013;381(9883):2083-90. Epub 2013/06/19. doi: 10.1016/s0140-6736(13)61127-7. PubMed PMID: 23769234.

9. Van Damme L, Corneli A, Ahmed K, Agot K, Lombaard J, Kapiga S, et al. Preexposure prophylaxis for HIV infection among African women. N Engl J Med. 2012;367(5):411-22. Epub 2012/07/13. doi: 10.1056/NEJMoa1202614. PubMed PMID: 22784040; PubMed Central PMCID: PMCPMC3687217.

10. Marrazzo JM, Ramjee G, Richardson BA, Gomez K, Mgodi N, Nair G, et al. Tenofovir-based preexposure prophylaxis for HIV infection among African women. N Engl J Med. 2015;372(6):509-18. Epub 2015/02/05. doi: 10.1056/NEJMoa1402269. PubMed PMID: 25651245; PubMed Central PMCID: PMCPMC4341965.

11. Sheth AN, Rolle CP, Gandhi M. HIV pre-exposure prophylaxis for women. J Virus Erad. 2016;2(3):149-55. Epub 2016/08/03. PubMed PMID: 27482454; PubMed Central PMCID: PMCPMC4967966.

12. Patterson KB, Prince HA, Kraft E, Jenkins AJ, Shaheen NJ, Rooney JF, et al. Penetration of tenofovir and emtricitabine in mucosal tissues: implications for prevention of HIV-1 transmission. Sci Transl Med. 2011;3(112):112re4. Epub 2011/12/14. doi: 10.1126/scitranslmed.3003174. PubMed PMID: 22158861; PubMed Central PMCID: PMCPMC3483088.

13. Glick JL, Russo RG, Huang AK, Jivapong B, Ramasamy V, Rosman LM, et al. ART uptake and adherence among female sex workers (FSW) globally: A scoping review. Glob Public Health. 2022;17(2):254-84. Epub 2020/12/11. doi: 10.1080/17441692.2020.1858137. PubMed PMID: 33301704; PubMed Central PMCID: PMCPMC8190161.

14. Chou R, Evans C, Hoverman A, Sun C, Dana T, Bougatsos C, et al. Preexposure Prophylaxis for the Prevention of HIV Infection: Evidence Report and Systematic Review for the US Preventive Services Task Force. Jama. 2019;321(22):2214-30. Epub 2019/06/12. doi: 10.1001/jama.2019.2591. PubMed PMID: 31184746.

15. Béhanzin L, Diabaté S, Minani I, Boily MC, Labbé AC, Ahoussinou C, et al. Decline in the prevalence of HIV and sexually transmitted infections among female sex workers in Benin over 15 years of targeted interventions. J Acquir Immune Defic Syndr. 2013;63(1):126-34. Epub 2013/01/23. doi: 10.1097/QAI.0b013e318286b9d4. PubMed PMID: 23337368; PubMed Central PMCID: PMCPMC3805545.

16. Rachakulla HK, Kodavalla V, Rajkumar H, Prasad SP, Kallam S, Goswami P, et al. Condom use and prevalence of syphilis and HIV among female sex workers in Andhra Pradesh, India - following a large-scale HIV prevention intervention. BMC Public Health. 2011;11 Suppl 6(Suppl 6):S1. Epub 2012/03/02. doi: 10.1186/1471-2458-11-s6-s1. PubMed PMID: 22376071; PubMed Central PMCID: PMCPMC3287547.

17. Huet C, Ouedraogo A, Konaté I, Traore I, Rouet F, Kaboré A, et al. Long-term virological, immunological and mortality outcomes in a cohort of HIV-infected female sex workers treated with highly active antiretroviral therapy in Africa. BMC Public Health. 2011;11:700. Epub 2011/09/16. doi: 10.1186/1471-2458-11-700. PubMed PMID: 21917177; PubMed Central PMCID: PMCPMC3191514.

18. Pickles M, Boily MC, Vickerman P, Lowndes CM, Moses S, Blanchard JF, et al. Assessment of the population-level effectiveness of the Avahan HIV-prevention programme in South India: a preplanned, causal-pathway-based modelling analysis. Lancet Glob Health. 2013;1(5):e289-99. Epub 2014/08/12. doi: 10.1016/s2214-109x(13)70083-4. PubMed PMID: 25104493.

19. Vickerman P, Terris-Prestholt F, Delany S, Kumaranayake L, Rees H, Watts C. Are targeted HIV prevention activities cost-effective in high prevalence settings? Results from a sexually transmitted infection treatment project for sex workers in Johannesburg, South Africa. Sex Transm Dis. 2006;33(10 Suppl):S122-32. Epub 2006/06/01. doi: 10.1097/01.olq.0000221351.55097.36. PubMed PMID: 16735954.

20. Fung IC, Guinness L, Vickerman P, Watts C, Vannela G, Vadhvana J, et al. Modelling the impact and cost-effectiveness of the HIV intervention programme amongst commercial sex workers in Ahmedabad, Gujarat, India. BMC Public Health. 2007;7:195. Epub 2007/08/09. doi: 10.1186/1471-2458-7-195. PubMed PMID: 17683595; PubMed Central PMCID: PMCPMC1999496.

21. Burgos JL, Gaebler JA, Strathdee SA, Lozada R, Staines H, Patterson TL. Costeffectiveness of an intervention to reduce HIV/STI incidence and promote condom use among female sex workers in the Mexico-US border region. PLoS One. 2010;5(6):e11413. Epub 2010/07/10. doi: 10.1371/journal.pone.0011413. PubMed PMID: 20617193; PubMed Central PMCID: PMCPMC2894974.

22. Cremin I, McKinnon L, Kimani J, Cherutich P, Gakii G, Muriuki F, et al. PrEP for key populations in combination HIV prevention in Nairobi: a mathematical modelling study. Lancet HIV. 2017;4(5):e214-e22. Epub 2017/02/25. doi: 10.1016/s2352-3018(17)30021-8. PubMed PMID: 28233660.

23. Eakle R, Gomez GB, Naicker N, Bothma R, Mbogua J, Cabrera Escobar MA, et al. HIV pre-exposure prophylaxis and early antiretroviral treatment among female sex workers in South Africa: Results from a prospective observational demonstration project. PLoS Med. 2017;14(11):e1002444. Epub 2017/11/22. doi: 10.1371/journal.pmed.1002444. PubMed PMID: 29161256.

24. Geidelberg L, Mitchell KM, Alary M, Mboup A, Béhanzin L, Guédou F, et al. Mathematical Model Impact Analysis of a Real-Life Pre-exposure Prophylaxis and Treatment-As-Prevention Study Among Female Sex Workers in Cotonou, Benin. J Acquir Immune Defic Syndr. 2021;86(2):e28-e42. Epub 2020/10/27. doi: 10.1097/qai.0000000000002535. PubMed PMID: 33105397; PubMed Central PMCID: PMCPMC7803451.

25. Programme national de lutte contre le sida (PNLS). Enquête de surveillance de deuxième génération relative aux IST, VIH et SIDA au Bénin (ESDG-2015): Professionnelles de Sexe & Serveuses de Bar et Restaurants. . In: Benin. MoH, editor. 2015.

26. Burrows D, McCallum, L., Parsons, D. & Falkenberry, H. (Global summary of findings of an assessment of HIV services packages for key populations in six regions. . APMG Health, Washington, DC., 2019.

27. UNAIDS. HIV and AIDS estimates Benin 2020. Available from: https://www.unaids.org/en/regionscountries/countries/benin.

28. Mboup A, Béhanzin L, Guédou FA, Geraldo N, Goma-Matsétsé E, Giguère K, et al. Early antiretroviral therapy and daily pre-exposure prophylaxis for HIV prevention among female sex workers in Cotonou, Benin: a prospective observational demonstration study. J Int AIDS Soc. 2018;21(11):e25208. Epub 2019/07/11. doi: 10.1002/jia2.25208. PubMed PMID: 31291057; PubMed Central PMCID: PMCPMC6287093.

29. Mboup A, Béhanzin L, Guédou F, Giguère K, Geraldo N, Zannou DM, et al. Comparison of adherence measurement tools used in a pre-exposure prophylaxis demonstration study among female sex workers in Benin. Medicine (Baltimore). 2020;99(21):e20063. Epub 2020/06/03. doi: 10.1097/md.0000000000020063. PubMed PMID: 32481273; PubMed Central PMCID: PMCPMC7249870.

30. Institut National de la statistique et d’analyse économique du Bénin (INSAE). Quatrième Recensement Général de la Population et de l’Habitation. https://www.insae-bj.org/publications/nos-bases-de-donnees. : 2013.

31. Institut National de la statistique et d’analyse économique du Bénin (INSAE). Recensement General de la Population et de l’Habitation 3eme Edition. 2002.

32. (INSAE); INdlsedaédB. Recensement General de la Population et de l’Habitation 2eme Edition. 1992.

33. Institut National de la statistique et d’analyse économique du Bénin (INSAE). Recensement General de la Population et de l’Habitation 1er Edition. 1979.

34. Projet Équité en Santé Sexuelle et Santé. Cartographie des Travailleuses de Sexe des communes de Cotonou, Abomey-Calavi, Sèmè-Podji et Parakou: Rapport Synthèse du Mapping. Benin. 2013-2014.

35. Lowndes CM, Alary M, Meda H, Gnintoungbé CAB, Mukenge-Tshibaka L, Adjovi C, et al. Role of core and bridging groups in the transmission dynamics of HIV and STIs in Cotonou, Benin, West Africa. Sex Transm Infect. 2002;78 Suppl 1(Suppl 1):i69–i77. doi: 10.1136/sti.78.suppl_1.i69. PubMed PMID: 12083450.

36. Programme national de lutte contre le sida (PNLS). Enquête de surveillance de deuxième génération des IST/VIH/SIDA au Bénin (ESDG-2012): Camionneurs et Clients des Travailleuses du sexe. 2012.

37. Alary M, Mukenge-Tshibaka L, Bernier F, Geraldo N, Lowndes CM, Meda H, et al. Decline in the prevalence of HIV and sexually transmitted diseases among female sex workers in Cotonou, Benin, 1993-1999. Aids. 2002;16(3):463-70. Epub 2002/02/09. doi: 10.1097/00002030-200202150-00019. PubMed PMID: 11834959.

38. Béhanzin L, Diabaté S, Minani I, Lowndes CM, Boily M-C, Labbé A-C, et al. Decline in HIV Prevalence among Young Men in the General Population of Cotonou, Benin, 1998–2008. PLOS ONE. 2012;7(8):e43818. doi: 10.1371/journal.pone.0043818.

39. Institut National de la Statistique et de l’Analyse Économique (INSAE). Enquête Démographique et de Santé du Bénin 2011-2012. . 2013.

40. Ministry of Health Benin. Rapport de l’audit de la file active des personnes vivant avec le VIH au Bénin (Audit report). 2017.

41. French N, Mujugira A, Nakiyingi J, Mulder D, Janoff EN, Gilks CF. Immunologic and clinical stages in HIV-1-infected Ugandan adults are comparable and provide no evidence of rapid progression but poor survival with advanced disease. J Acquir Immune Defic Syndr. 1999;22(5):509-16. Epub 2000/08/29. doi: 10.1097/00126334-199912150-00013. PubMed PMID: 10961614.

42. Lawn SD, Myer L, Orrell C, Bekker LG, Wood R. Early mortality among adults accessing a community-based antiretroviral service in South Africa: implications for programme design. Aids. 2005;19(18):2141-8. Epub 2005/11/15. doi: 10.1097/01.aids.0000194802.89540.e1. PubMed PMID: 16284464.

43. Glynn JR, Sonnenberg P, Nelson G, Bester A, Shearer S, Murray J. Survival from HIV-1 seroconversion in Southern Africa: a retrospective cohort study in nearly 2000 goldminers over 10 years of follow-up. Aids. 2007;21(5):625-32. Epub 2007/02/23. doi: 10.1097/QAD.0b013e328017f857. PubMed PMID: 17314525.

44. Todd J, Glynn JR, Marston M, Lutalo T, Biraro S, Mwita W, et al. Time from HIV seroconversion to death: a collaborative analysis of eight studies in six low and middleincome countries before highly active antiretroviral therapy. AIDS (London, England). 2007;21 Suppl 6(Suppl 6):S55–S63. doi: 10.1097/01.aids.0000299411.75269.e8. PubMed PMID: 18032940.

45. Lavreys L, Baeten JM, Chohan V, McClelland RS, Hassan WM, Richardson BA, et al. Higher set point plasma viral load and more-severe acute HIV type 1 (HIV-1) illness predict mortality among high-risk HIV-1-infected African women. Clin Infect Dis. 2006;42(9):1333-9. Epub 2006/04/06. doi: 10.1086/503258. PubMed PMID: 16586394.

46. May M, Wood R, Myer L, Taffé P, Rauch A, Battegay M, et al. CD4(+) T cell count decreases by ethnicity among untreated patients with HIV infection in South Africa and Switzerland. J Infect Dis. 2009;200(11):1729-35. Epub 2009/10/24. doi: 10.1086/648096. PubMed PMID: 19848608; PubMed Central PMCID: PMCPMC3056076.

47. Morgan D, Mahe C, Mayanja B, Okongo JM, Lubega R, Whitworth JA. HIV-1 infection in rural Africa: is there a difference in median time to AIDS and survival compared with that in industrialized countries? AIDS. 2002;16(4):597-603. Epub 2002/03/02. doi: 10.1097/00002030-200203080-00011. PubMed PMID: 11873003.

48. Pantazis N, Morrison C, Amornkul PN, Lewden C, Salata RA, Minga A, et al. Differences in HIV Natural History among African and Non-African Seroconverters in Europe and Seroconverters in Sub-Saharan Africa. PLOS ONE. 2012;7(3):e32369. doi: 10.1371/journal.pone.0032369.

49. Peters PJ, Karita E, Kayitenkore K, Meinzen-Derr J, Kim DJ, Tichacek A, et al. HIV- infected Rwandan women have a high frequency of long-term survival. Aids. 2007;21 Suppl 6:S31-7. Epub 2008/01/11. doi: 10.1097/01.aids.0000299408.52399.e1. PubMed PMID: 18032936.

50. Van der Paal L, Shafer LA, Todd J, Mayanja BN, Whitworth JA, Grosskurth H. HIV- 1 disease progression and mortality before the introduction of highly active antiretroviral therapy in rural Uganda. Aids. 2007;21 Suppl 6:S21-9. Epub 2007/12/06. doi: 10.1097/01.aids.0000299407.52399.05. PubMed PMID: 18032935.

51. GBD 2016 Disease and Injury Incidence and Prevalence Collaborators. Global, regional, and national incidence, prevalence, and years lived with disability for 328 diseases and injuries for 195 countries, 1990–2016: a systematic analysis for the Global Burden of Disease Study 2016. Lancet. 2017;390:1211–59. doi: 10.1016/S0140-6736(17)32154-2.

52. The World Bank. World development Indicators, . Available from: https://databank.worldbank.org/source/world-development-indicators.

53. Central Bank of West African States. Main Indicators and Interest rates 2016 [November 2016]. Available from: https://www.bceao.int/en/content/main-indicators-and-interest-rates.

54. Ochalek J LJ, Claxton KP. Cost per DALY averted thresholds for low- and middleincome countries: evidence from cross country data. 2015.

55. Bertram MY LJ, De Joncheere K, Edejer T, Hutubessy R, Kieny MP, Hill SR. . Costeffectiveness thresholds: pros and cons. . Bulletin World Health Organization [Internet]. 2016; 94(12):[925-30 pp.]. Available from: https://www.who.int/bulletin/volumes/94/12/15-164418/en/.

56. Terris-Prestholt F. Mathematical Modelling to Estimate the Impact and Costeffectiveness of TasP, PrEP and Condom Promotion for Serodiscordant Couples in Nigeria. HIV Research for Prevention, South Africa2014.

57. Jamieson L, Gomez GB, Rebe K, Brown B, Subedar H, Jenkins S, et al. The impact of self-selection based on HIV risk on the cost-effectiveness of preexposure prophylaxis in South Africa. Aids. 2020;34(6):883-91. Epub 2020/02/01. doi: 10.1097/qad.0000000000002486. PubMed PMID: 32004205.

58. Mangenah C, Nhamo D, Gudukeya S, Gwavava E, Gavi C, Chiwawa P, et al. Efficiency in PrEP Delivery: Estimating the Annual Costs of Oral PrEP in Zimbabwe. AIDS Behav. 2022;26(1):161-70. Epub 2021/08/29. doi: 10.1007/s10461-021-03367-w. PubMed PMID: 34453240; PubMed Central PMCID: PMCPMC8786759.

59. Chen A KG, Mwai D, Dutta A. Cost of providing oral preexposure prophylaxis to prevent HIV infection among sex workers in Kenya. . Health Policy Project, 2014.

