## supplementary materials for "Modelling the cost-effectiveness of TasP and PrEP in female sex workers in Cotonou, Benin"

### Section 1: Model structure

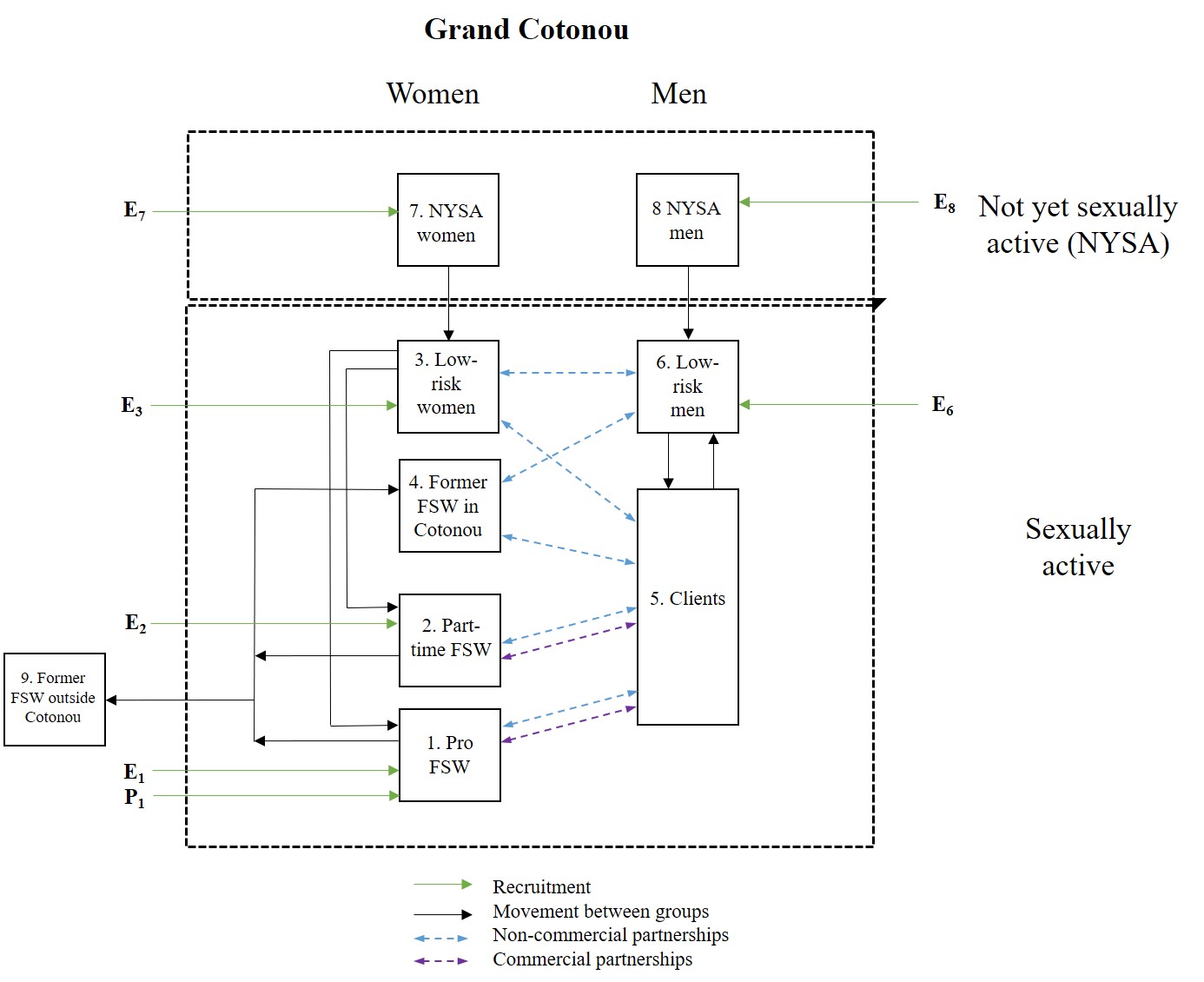

**Figure S1.** Recruitment, movement and sexual mixing between groups, represented by green, black and blue lines respectively. Groups within the dashed rectangles form the sexual transmission network of Grand Cotonou. $E_{i}$ represents the numbers of HIV- individuals in group i recruited to the model (Equation 2a); $P_{1}$ represents the number of HIV+ professional FSW recruited (Equation 2b). Commercial partnerships are only formed between professional and part-time FSW and clients. NYSA = not yet sexually active. FSW = female sex worker.

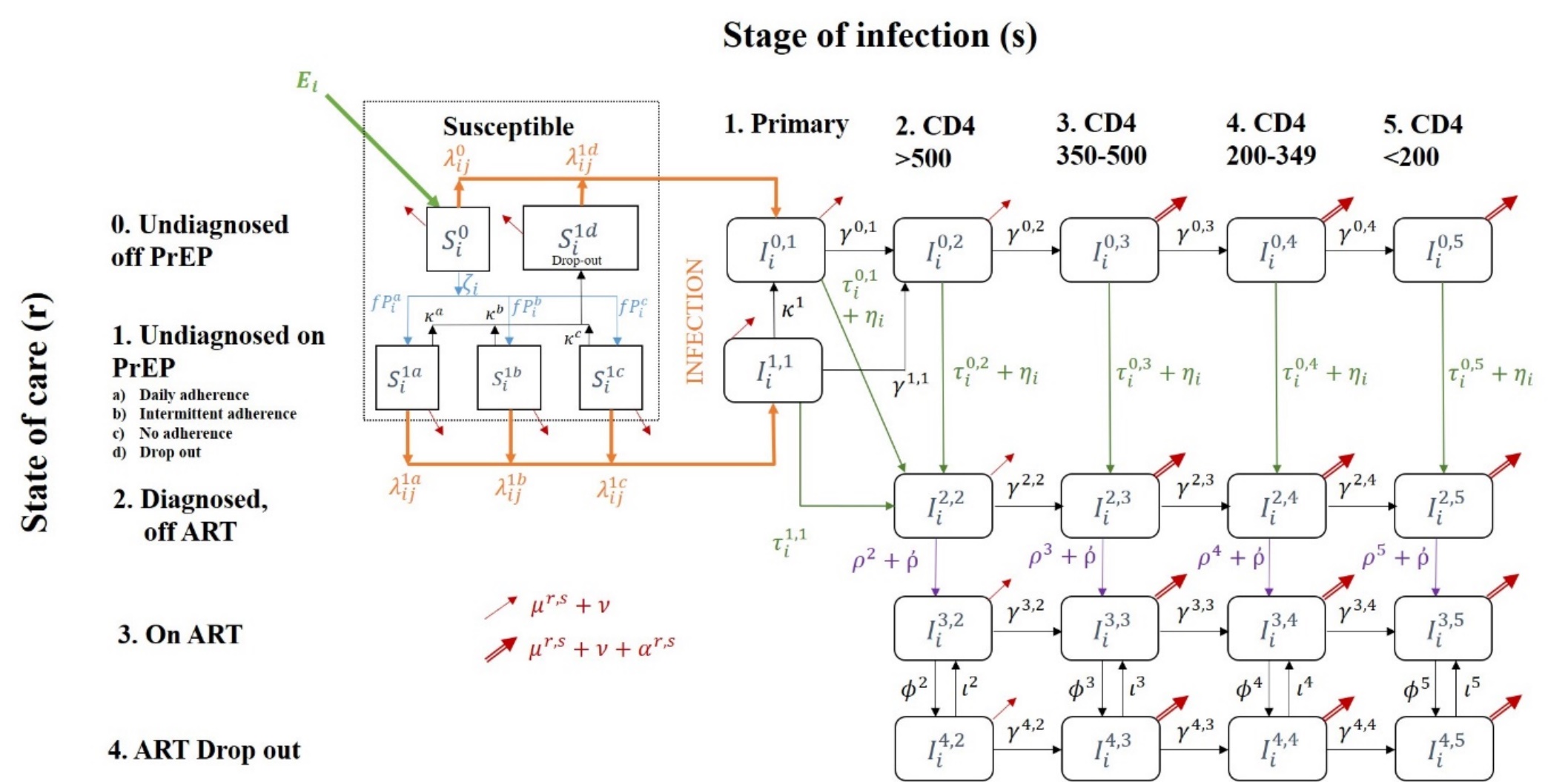

F**igure S2.** The model flow diagram, representing progression through state of care (r) and stage of infection (s). PrEP initiation, HIV testing, and ART initiation are represented by blue, green and purple arrows respectively. Red arrows represent mortality and ageing; double red arrow indicates increased mortality due to HIV infection. Orange arrows represent HIV infection. All risk groups are represented by these compartments, but some compartments will always be empty for some groups (i.e. “on PrEP” for groups who are not professional FSW). Not shown in diagram is recruitment of HIV+ professional FSW into compartments $I^{01}$ - $I^{05}$ (Equations 9, 11-14).

#### Model description

We developed a deterministic model of sexual HIV transmission and HIV interventions (PrEP, ART, condom use) in the heterosexual adult population of Grand Cotonou (abbreviated GC; comprising Cotonou Centrale, Abomey-Calavi and Seme-Kpodji). The model represents an open and growing population aged 15-59 years old, stratified in 9 risk groups (subscript *i*): 2 types of active female sex workers (professional, part-time), their male clients, 2 types of former FSWs (those who remain and those who leave GC), low-risk women and men, not yet sexually active (NYSA) women and men.

New susceptible individuals can join the population at time varying rates that reflect changes in the total population size of Cotonou. They are distributed between six risk groups (NYSA women & men, low-risk women & men and professional & low-level FSWs) according to relative population sizes. $E_{i}$ represents the number of new susceptible recruits joining the population in each group *i*. NYSA women and men enter the sexually active but susceptible low-risk population at per capita rates $u_{7}$ and $u_{8}$ determined by the average age at sexual debut, respectively. Professional and part-time FSWs are recruited directly from the low-risk women population of GC at low-risk population HIV prevalence at per capita rates $v_{1,3}$and $v_{2,3}$_,_ respectively, or from outside of GC (from both inside and outside Benin) at rates $E_{1}$ and $E_{2}$ respectively, reflecting HIV prevalence of neighbouring countries at time *t*. Professional and part-time FSW cease sex work and join the former FSW categories at rates $u_{1}$ and $u_{2}$, reflecting duration in sex work, respectively; the factor $p_{foreign}$ determines the fraction that leaves GC upon ceasing sex work. Low-risk population males become clients at a per capita rate $u_{6}$ and return at per capita rate $u_{5}$, reflecting duration of paying for sex. Individuals leave the modelled population due to aging, HIV-related and unrelated mortality. Movement between groups applies equally to all care states and stages of infection.

Apart from a fraction of professional FSW, all individuals enter the population in the susceptible off PrEP category ($S_{i}^{0}$). Susceptible professional FSWs can initiate PrEP at per capita rate $\zeta_{i}$, depending on background testing rate $\tau_{i}\left( t \right)$ of group *i* at time *t*, testing rate due to the PrEP/TasP study $\eta_{i}\left( t \right)$, PrEP offering rate $\xi_{i}(t)$ and PrEP acceptance rate $\sigma_{i}$, and the fraction $fP_{i}^{a}$, $fP_{i}^{b}$, $fP_{i}^{c}$, that are initially perfectly, intermediately and non-adherent (though receiving pills), moving into states $S_{i}^{1a}$, $S_{i}^{1b}$ and $S_{i}^{1c}$ respectively. The data indicates decreasing PrEP adherence over time, thus PrEP users can move into lower adherence categories (at rates $\psi^{a}$ and $\psi^{b}$ moving from perfect to intermediate and from intermediate to non adherence respectively) or cease to use PrEP and move to the dropout PrEP category, at per capita rates $\kappa^{a}$, $\kappa^{b}$, $\kappa^{c}$. Professional FSWs can retire from sex work and move to the former FSW groups inside or outside GC; in this case they do not move to their associated PrEP category, but rather into their susceptible off PrEP category ($S_{i}^{0}$).

Sexually active susceptible individuals are assumed to get infected at a per capita force of infection $\lambda_{ij}^{rs}$ that depends on their PrEP status, number and type of sexual partners, HIV prevalence among partners, sexual mixing patterns between risk groups, the fraction of sex acts effectively protected by condoms, the levels of PrEP use and efficacy (related to adherence status), the partner's infectiousness (i.e., varying by disease stage, ART treatment). The protective effect of male circumcision on HIV acquisition risk was captured in the per-act HIV transmission probability since more than 99% of men in GC are circumcised.^1,2^ Only vaginal sex was modelled, as the available data suggested no meaningful frequency of oral or anal sex.^1-14^ Infected individuals are represented in the model as $I_{i}^{r,s}$, for risk group *i*, care state *r* and stage of infection *s*.

Following infection, the model represents the course of disease progression stratified by CD4 cell count levels and different levels of engagement in the care and treatment cascade. Untreated undiagnosed HIV-positive individuals progress through a short highly infectious primary infection followed by 4 longer disease stages defined based on CD4 cell counts, at rates $\gamma^{r,s}$ that reflect care status (*r*) and stage of infection (*s*). Individuals in infected stages CD4 350-500, CD4 200-349 and CD4 <200 (*s* = 3, 4, 5 respectively) die at AIDS mortality rate $\alpha^{rs}$.

When infected, individuals on PrEP enter a distinct primary phase of infection compared to those not on PrEP to reflect the fact that they are tested at an increased rate. These individuals leave this compartment at testing rate $\tau_{i}^{1,1}\left( t \right)$, dropout of PrEP at rate $\kappa^{1}$, or progress to CD4 >500 at rate $\gamma_{i}^{1,1}$.

All undiagnosed individuals in these 5 stages are tested for HIV at a per capita rate $\tau_{i}^{rs}$, depending on group *i*, care status *r* and stage of infection *s*, moving to the diagnosed off ART category, who experience the same disease progression as undiagnosed individuals. When modelling the intervention, individuals experience an increased testing rate of $\eta_{i}\left( t \right)$.

Diagnosed individuals initiate treatment at a per capita rate $\rho_{i}^{s}$ that depends on group *i*, calendar year (due to changes in eligibility criteria) and stage of infection *s*. When modelling the intervention, individuals experience an increased ART initiation rate of $ῤ\left( t \right)$. Compared to those not on ART, treated individuals have slower rates of disease progression and reduced HIV-related mortality, represented by factors ${prog}_{ART}$ and ${mort}_{ART}$ respectively. Finally, some individuals may experience a therapeutic failure or discontinue ART, at rate $\varphi^{s}$, in which case the disease follows its natural progression unless these individuals re-initiate treatment at rate $\iota_{i}^{s}$.

Individuals leave the population at per capita rates $\nu$, $\mu^{rs}$ and $\alpha^{rs}$, which represent aging, background and AIDS-related mortalities, respectively. The full system of ODEs is presented in Section 2.

HIV was seeded at the initial prevalence $\pi_{i}$, and the system of ODEs was solved numerically from 1986 – 2035 using the lsoda algorithm (Petzhold and Hindmarsh) with a variable time step.^15^ The model was coded in R, using “odin”,^16^ a wrapper around the deSolve package for solving ordinary differential equations. ^17^

### Section 2: Equations

#### S2a: Ordinary differential equations (ODEs)

Figure S2 shows the care state *r* and stage of infection *s* categories for each group *i*. $S_{i}^{r}$ and $I_{i}^{r,s}$ represent susceptible and infected individuals respectively.

Testing/ART care-state (*r*):

0. Off PrEP, undiagnosed
1. On PrEP, undiagnosed
   a. Perfect adherence (daily dosing)
   b. Intermediate adherence
   c. Non-adherence (undetectable)
   d. PrEP Dropout
2. Diagnosed, off ART
3. Diagnosed, on ART
4. ART dropout

Stage of infection (*s*):

1. Primary phase
2. CD4 >500 cells/μl
3. CD4 350-500 cells/μl
4. CD4 200-349 cells/μl
5. CD4 <200 cells/μl

|  | $N\left( t \right)=\sum_{i=1}^{i=8} \sum_{r=0}^{r=4} \left( S_{i}^{r}\left( t \right)+\sum_{s=1}^{s=5} I_{i}^{rs}(t) \right)$ | 1 |
| --- | --- | --- |

N(t) is the total population of GC at time *t*; and $S_{i}^{r}\left( t \right)$ and $I_{i}^{rs}\left( t \right)$ are the susceptible and infected populations respectively, of group *i,* care state *r* and disease state *s*. Note that N(t) does not include former FSWs outside of GC (*i*=9).

The following equations describe the recruitment of new individuals ($E(t)$) into the population. Due to the population growth of GC, new people are added to the population every timestep dependent on the growth rate $\varepsilon(t)$.

|  | $E_{i}\left( t \right)=\left( \varepsilon\left( t \right)+\mu_{i}+\nu\right) N\left( t \right) \omega_{i}, i\neq1$ | 2a |
| --- | --- | --- |

Where $E(t)$ is the number of new susceptible people entering the sexually active population into group *i*; $\varepsilon(t)$ is the total population growth rate; $\mu_{i}$ is the natural mortality rate; $\nu$ is the rate of exit due to ageing; $N\left( t \right)$ is the total population size at time *t* as defined in Equation 1; $\omega_{i}$ is the proportion of new people distributed into group *i*.

Professional FSW

In order for the numbers of professional FSWs to be maintained, they have an extra term to account for those who retire from sex work and leave Grand Cotonou. Additionally, professional FSWs enter the model at a prevalence of the neighbouring countries to Benin (Nigeria, Togo and Ghana) at time *t*:

|  | $F_{i}\left( t \right)=\left( \varepsilon\left( t \right)+\mu_{i}+\nu\right) N\left( t \right) \omega_{i}+\frac{1}{dur_{PFSW}}*N_{PFSW}\left( t \right)*frac_{FSWforeign}, i=1$ | 2bi |
| --- | --- | --- |
|  | $E_{i}\left( t \right)=F_{i}\left( t \right)*\left( 1-prev_{FSW_{foreign}}\left( t \right) \right), i=1$ | 2bii |
|  | $P_{i}\left( t \right)=F_{i}\left( t \right)*prev_{FSW_{foreign}}\left( t \right), i=1$ | 2biii |
|  | $F_{i}\left( t \right)=0, i\neq1$ | 2biv |
|  | $P_{i}\left( t \right)=0, i\neq1$ | 2bv |

Where $F_{i}\left( t \right)$ is the total number of new individuals in group *i* entering the model at time *t*, $E_{i}\left( t \right)$ is the number of susceptible individuals in group *i* entering the model at time *t*, $P_{i}\left( t \right)$ is the number of infected individuals in group *i* entering the model at time *t*, $dur_{PFSW}$ is the duration of sex work for professional FSWs, $N_{PFSW}(t)$ is the number of professional FSW at time *t*, $frac_{FSWforeign}$ is the fraction of FSW that are foreign-born and $prev_{FSW_{foreign}}(t)$ is the prevalence of FSWs in neighbouring countries.

**Susceptible:**

**Susceptible off PrEP:**

| $\frac{dS_{i}^{0}(t)}{dt}= E_{i}(t)-S_{i}^{0}\left( t \right)\left( \sum_{j} \left( \lambda_{ij}^{0}\left( t \right) \right)+\mu_{i}+\zeta_{i}+\nu\right)+T_{i}^{0}$ | 3 |
| --- | --- |

Where $E(t)$ is the number of new susceptible people entering the sexually active population into group *i* as described in Equation 2; where $\mu_{i}$ is the background mortality rate of group *i*; $\zeta_{i}$ is the PrEP initiation rate; $\lambda_{ij}^{0}\left( t \right)$ is the force of infection from group *j* to group *i*;$\nu$ is the rate of exit due to ageing; $T_{i}^{as}$ is the sum of the movement of people to and from group *i*.

The PrEP initiation rate $\zeta_{i}$ is composed of background testing rate $\tau_{i}\left( t \right)$ of group *i* at time *t*, testing rate due to PrEP/TasP intervention $\eta_{i}\left( t \right)$, PrEP offering rate $\xi_{i}(t)$ and PrEP acceptance rate $\sigma_{i}$.

| $\zeta_{i}=\left( \tau_{i}\left( t \right)+\eta_{i}\left( t \right) \right)*\xi_{i}(t)*\sigma_{i}$ | 4 |
| --- | --- |

**Susceptible on PrEP (perfect adherence):**

| $\frac{dS_{i}^{1a}(t)}{dt}=\zeta_{i}\left( t \right) {fP}_{i}^{a}S_{i}^{0}\left( t \right)-S_{i}^{1a}\left( t \right)\left( \sum_{j} \left( \lambda_{ij}^{1a}\left( t \right) \right)+\mu_{i}+ \kappa_{i}^{a}+\nu\right)+T_{i}^{1a}$ | 5 |
| --- | --- |

Where ${fP}_{i}^{a}$ is the fraction of individuals in the perfect PrEP adherence category and $\kappa_{i}^{r}$ is rate of PrEP dropout.

**Susceptible on PrEP (intermediate adherence):**

| $\frac{dS_{i}^{1b}(t)}{dt}= \zeta_{i}{\left( t \right) {fP}_{i}^{b} S}_{i}^{0}\left( t \right)-S_{i}^{1b}\left( t \right) \left( \sum_{j} \left( \lambda_{ij}^{1b}\left( t \right) \right)+\mu_{i}+\kappa_{i}^{b}+\nu\right)+T_{i}^{1b,0}$ | 6 |
| --- | --- |
| Where ${fP}_{i}^{b}$ is the fraction of individuals in the intermediate PrEP adherence category. |  |

**Susceptible on PrEP (non-adherence):**

| $\frac{dS_{i}^{1c}(t)}{dt}= \zeta_{i}{\left( t \right) {fP}_{i}^{c} S}_{i}^{0}\left( t \right)-S_{i}^{1c}\left( t \right)\left( \sum_{j} \left( \lambda_{ij}^{1c}\left( t \right) \right)+\mu_{i}+ \kappa_{i}^{c}+\nu\right)+T_{i}^{1c,0}$ | 7 |
| --- | --- |

Where ${fP}_{i}^{c}$ is the fraction of individuals in the non-adherence to PrEP category.

**Susceptible PrEP drop-out:**

| $\frac{dS_{i}^{1d}(t)}{dt}= \kappa_{i}^{a}\left( t \right)S_{i}^{1a}\left( t \right)+\kappa_{i}^{b}\left( t \right)S_{i}^{1b}\left( t \right)+\kappa_{i}^{c}\left( t \right)S_{i}^{1c}\left( t \right)-S_{i}^{1d}\left( t \right)\left( \sum_{j} \left( \lambda_{ij}^{1d}\left( t \right) \right)+\mu_{i}+\nu\right)+T_{i}^{1c,0}$ | 8 |
| --- | --- |

Note that $\lambda_{ij}^{0}\left( t \right)$ and $\lambda_{ij}^{1d}\left( t \right)$ are equivalent, as those who have dropped out of PrEP do not have any PrEP protection; the PrEP dropout group is however retained for the purposes of determining cost-effectiveness.

**Primary infection:**

As detailed in Equation 2biii, the number of new infected professional FSWs entering the model depends on a changing prevalence in neighbouring countries over time ($P_{i}\left( t \right)=F_{i}\left( t \right)*prev_{FSW_{foreign}}\left( t \right)$ for i=1). There are no infected individuals entering the model from any other group ($P_{i}\left( t \right)=0$ for i≠1). The infected professional FSWs entering the model are divided into the 5 stages of infection in the undiagnosed care state, proportional to the relative duration of each disease state ($d^{s}$).

**Primary infection off PrEP**

| $\frac{dI_{i}^{0,1}\left( t \right)}{dt}=d^{1} P_{i}\left( t \right)+S_{i}^{0}\left( t \right)\sum_{j} \left( \lambda_{ij}^{0}\left( t \right) \right)+S_{i}^{1d}\left( t \right)\sum_{j} \left( \lambda_{ij}^{1d}\left( t \right) \right)-I_{i}^{0,1}\left( t \right) \left( \gamma^{0,1}+\tau_{i}^{0,1}\left( t \right)+\eta_{i}\left( t \right)+ \mu_{i}+\nu\right)+I^{1,1}\left( t \right)\kappa^{1}+T_{i}^{0,1}\left( t \right)$ | 9 |
| --- | --- |

Where $\gamma^{rs}$ is disease progression from care state *r* and disease stage *s*; $\tau_{i}^{as}\left( t \right)$ is testing rate; $\alpha_{i}^{r}$ is mortality from AIDS in care state *r*; $\kappa^{1}$ is the PrEP dropout rate for infected individuals.

**Primary infection on PrEP**

| $\frac{dI_{i}^{1,1}\left( t \right)}{dt}=S_{i}^{1a}\left( t \right)\sum_{j} \left( \lambda_{ij}^{1a}\left( t \right) \right)+S_{i}^{1b}\left( t \right)\sum_{j} \left( \lambda_{ij}^{1b}\left( t \right) \right)+S_{i}^{1c}\left( t \right)\sum_{j} \left( \lambda_{ij}^{1c}\left( t \right) \right)-I_{i}^{1,1}\left( t \right) \left( \gamma^{1,1}+\kappa^{1}+\tau_{i}^{1,1}\left( t \right)+ \mu_{i}+\nu\right)+T_{i}^{1,1}(t)$ | 10 |
| --- | --- |

**Not on PrEP, undiagnosed infection:**

**CD4 > 500 off PrEP**

| $\frac{dI_{i}^{0,2}\left( t \right)}{dt}=d^{2} P_{i}\left( t \right)+{\gamma^{0,1} I}_{i}^{0,1}\left( t \right)+ {\gamma^{1,1} I}_{i}^{1,1}\left( t \right)-I_{i}^{0,2}(t)\left( \gamma^{0,2}+\tau_{i}^{0,2}\left( t \right)+\eta_{i}\left( t \right)+\mu_{i}+\nu\right)+T_{i}^{0,2}(t)$ | 11 |
| --- | --- |

**CD4 350 - 500 off PrEP**

| $\frac{dI_{i}^{0,3}\left( t \right)}{dt}=d^{3} P_{i}\left( t \right)+{\gamma^{0,2} I}_{i}^{0,2}\left( t \right)-I_{i}^{0,3}(t)\left( \gamma^{0,3}+\tau_{i}^{0,3}\left( t \right)+\eta_{i}\left( t \right)+\alpha_{i}^{0,3}+\mu_{i}+\nu\right)+T_{i}^{0,3}(t)$ | 12 |
| --- | --- |

**CD4 200 - 349 off PrEP**

| $\frac{dI_{i}^{0,4}\left( t \right)}{dt}=d^{4} P_{i}\left( t \right)+{\gamma^{0,3} I}_{i}^{0,3}\left( t \right)-I_{i}^{0,4}(t)\left( \gamma^{0,4}+\tau_{i}^{0,4}\left( t \right)+\eta_{i}\left( t \right)+\alpha_{i}^{0,4}+\mu_{i}+\nu\right)+T_{i}^{0,4}(t)$ | 13 |
| --- | --- |

**CD4 < 200 off PrEP**

| $\frac{dI_{i}^{0,5}\left( t \right)}{dt}=d^{5} P_{i}\left( t \right)+{\gamma^{0,4} I}_{i}^{0,4}\left( t \right)-I_{i}^{0,5}(t)\left( \tau_{i}^{0,5}\left( t \right)+\eta_{i}\left( t \right)+\alpha_{i}^{0,5}+\mu_{i}+\nu\right)+T_{i}^{0,5}(t)$ | 14 |
| --- | --- |

**Diagnosed infection, not on ART:**

**CD4 > 500 diagnosed, off ART**

| $\frac{dI_{i}^{2,2}\left( t \right)}{dt}=\left( \tau_{i}^{0,1}\left( t \right)+\eta_{i}\left( t \right) \right)I^{0,1}\left( t \right)+\tau_{i}^{1,1}\left( t \right)I^{1,1}\left( t \right)+\left( \tau_{i}^{0,2}\left( t \right)+\eta_{i}\left( t \right) \right) I^{0,2}(t)-I_{i}^{2,2}(t)\left( \gamma^{2,2}+\rho_{i}^{2}\left( t \right)+ῤ_{i}(t)+\mu_{i}+\nu\right)+T_{i}^{2,2}(t)$ | 15 |
| --- | --- |

Where $\rho_{i}^{s}\left( t \right)$ is rate of ART uptake in group *i*, disease state *s*.

**CD4 350 - 500 diagnosed, off ART**

| $\frac{dI_{i}^{2,3}\left( t \right)}{dt}={\gamma^{2,2} I}_{i}^{2,2}(t)+\left( \tau_{i}^{0,3}\left( t \right)+\eta_{i}\left( t \right) \right)I^{0,3}(t)-I_{i}^{2,3}(t)\left( \gamma^{2,3}+\rho_{i}^{3}\left( t \right)+ῤ_{i}(t)+\alpha_{i}^{0,3}+ \mu_{i}+\nu\right)+T_{i}^{2,3}(t)$ | 16 |
| --- | --- |

**CD4 200 - 349 diagnosed, off ART**

| $\frac{dI_{i}^{2,4}\left( t \right)}{dt}={\gamma^{2,3} I}_{i}^{2,3}(t)+\left( \tau_{i}^{0,4}\left( t \right)+\eta_{i}\left( t \right) \right)I^{0,4}(t)-I_{i}^{2,4}(t)\left( \gamma^{2,4}+\rho_{i}^{4}\left( t \right)+ῤ_{i}(t)+\alpha_{i}^{0,4}+ \mu_{i}+\nu\right)+T_{i}^{2,4}(t)$ | 17 |
| --- | --- |

**CD4 < 200 diagnosed, off ART**

| $\frac{dI_{i}^{2,5}\left( t \right)}{dt}={\gamma^{2,4} I}_{i}^{2,4}(t)+\left( \tau_{i}^{0,5}\left( t \right)+\eta_{i}\left( t \right) \right)I^{0,5}(t)-I_{i}^{2,5}(t)\left( \rho_{i}^{5}\left( t \right)+ῤ_{i}(t)+\alpha_{i}^{0,5}+ \mu_{i}+\nu\right)+T_{i}^{2,5}(t)$ | 18 |
| --- | --- |

**Diagnosed infection, on ART:**

**CD4 >500 diagnosed, on ART**

| $\frac{dI_{i}^{3,2}\left( t \right)}{dt}=\left( \rho_{i}^{2}\left( t \right)+ῤ_{i}(t) \right)I_{i}^{2,2}\left( t \right)+\iota_{i}^{2}I_{i}^{4,2}(t)-I_{i}^{3,2}(t)\left( \gamma^{3,2}+\varphi_{i}^{2}\left( t \right)+ \mu_{i}+\nu\right)+T_{i}^{3,2}(t)$ | 19 |
| --- | --- |

Where $\iota_{i}^{s}\left( t \right)$ is rate of ART re-initialisation in group *i*, disease state *s*; $\varphi_{i}^{s}$ is rate of ART dropout.

**CD4 350 - 500 diagnosed, on ART**

| $\frac{dI_{i}^{3,3}\left( t \right)}{dt}=\gamma^{3,2} I_{i}^{3,2}(t)+\left( \rho_{i}^{3}\left( t \right)+ῤ_{i}(t) \right)I_{i}^{2,3}\left( t \right)+\iota_{i}^{3}I_{i}^{4,3}(t)-I_{i}^{3,3}(t)\left( \gamma^{3,3}+\varphi_{i}^{3}\left( t \right)+\alpha_{i}^{3,3}+ \mu_{i}+\nu\right)+T_{i}^{3,3}(t)$ | 20 |
| --- | --- |

**CD4 200 - 349 diagnosed, on ART**

| $\frac{dI_{i}^{3,4}\left( t \right)}{dt}=\gamma^{3,3}I_{i}^{3,3}\left( t \right)+\left( \rho_{i}^{4}\left( t \right)+ῤ_{i}(t) \right)I_{i}^{2,4}\left( t \right)+\iota_{i}^{4}I_{i}^{4,4}(t)-I_{i}^{3,4}(t)\left( \gamma^{3,4}+\varphi_{i}^{4}\left( t \right)+\alpha_{i}^{3,4}+\mu_{i}+\nu\right)+T_{i}^{3,4}(t)$ | 21 |
| --- | --- |

**CD4 < 200 diagnosed, on ART**

| $\frac{dI_{i}^{3,5}\left( t \right)}{dt}=\gamma^{3,4}I_{i}^{3,4}\left( t \right)+\left( \rho_{i}^{5}\left( t \right)+ῤ_{i}(t) \right)I_{i}^{2,5}\left( t \right)+\iota_{i}^{5}I_{i}^{4,5}(t)-I_{i}^{3,5}(t)\left( \varphi_{i}^{5}\left( t \right)+\alpha_{i}^{3,5}+\mu_{i}+\nu\right)+T_{i}^{3,5}(t)$ | 22 |
| --- | --- |

**Diagnosed infection, ART dropout / therapeutic failure:**

**CD4 > 500 ART drop out / therapeutic failure**

| $\frac{dI_{i}^{4,2}\left( t \right)}{dt}=\varphi_{i}^{2}\left( t \right) I_{i}^{3,2}(t)-I_{i}^{4,2}(t)\left( \gamma^{4,2}+\iota_{i}^{2}\left( t \right)+\mu_{i}+\nu\right)+T_{i}^{4,2}(t)$ | 23 |
| --- | --- |

**CD4 350 - 500 ART drop out / therapeutic failure**

| $\frac{dI_{i}^{4,3}\left( t \right)}{dt}={\gamma^{4,2} I_{i}^{4,2}(t)+\varphi}_{i}^{3}\left( t \right)I_{i}^{3,3}(t)-I_{i}^{4,3}(t)\left( \gamma^{4,3}+\iota_{i}^{3}\left( t \right)+\alpha_{i}^{0,3}+ \mu_{i}+\nu\right)+T_{i}^{4,3}(t)$ | 24 |
| --- | --- |

**CD4 200 - 349 ART drop out / therapeutic failure**

| $\frac{dI_{i}^{4,4}\left( t \right)}{dt}={\gamma^{4,3} I_{i}^{4,3}(t)+\varphi}_{i}^{4}\left( t \right)I_{i}^{3,4}(t)-I_{i}^{4,4}(t)\left( \gamma^{4,4}+\iota_{i}^{4}\left( t \right)+\alpha_{i}^{0,4}+ \mu_{i}+\nu\right)+T_{i}^{4,4}(t)$ | 25 |
| --- | --- |

**CD4 < 200 ART drop out / therapeutic failure**

| $\frac{dI_{i}^{4,5}\left( t \right)}{dt}={\gamma^{4,4} I_{i}^{4,4}(t)+\varphi}_{i}^{5}\left( t \right)I_{i}^{3,5}(t)-I_{i}^{4,5}(t)\left( \iota_{i}^{5}\left( t \right)+\alpha_{i}^{0,5}+ \mu_{i}+\nu\right)+T_{i}^{4,5}(t)$ | 26 |
| --- | --- |

#### S2b: Linear interpolation of parameters

We assume that certain parameters vary over time. For example, the fraction of sex acts that are protected by a condom will generally increase over time. Estimates for this fraction will be taken from surveys (e.g. 2008 and 2011). We linearly interpolate between these years in order to get a value for every year. We assume all parameters to remain fixed at their 2015 value when simulating the epidemic beyond this year.

For non-commercial condom use ($f_{ij}^{N}\left( t \right)$), we calculate its value over time as follows:

$$f_{ij}^{N}\left( t \right)=\left\{ \begin{aligned} f_{ij1998}^{N}, t<2008 \\ f_{ij1998}^{N}+zt, 2008\leq t\leq2011 \\ f_{ij2008}^{N}, t>2011 \end{aligned} \right.$$

Where *z* is the annual increase in condom use, calculated as follows:

$$z= \frac{f_{ij2008}^{N}-f_{ij1998}^{N}}{2011-2008}$$

#### S2c: Entering the population

New people entering the population are distributed into the 9 groups at fractions determined by the parameter $\omega_{i}$.

$$\omega_{PFSW}= frac_{F}*frac_{FPFSW}*frac_{FSWforeign}$$

$$\omega_{LFSW}= frac_{F}*frac_{FPFSW}*ratio_{LFSW}*frac_{FSWforeign}$$

$$\omega_{GPF}= frac_{F}-\omega_{PFSW}-\omega_{LFSW}-\omega_{VF}$$

$$\omega_{GPM}=\left( 1-frac_{F} \right)*frac_{MSA}$$

$$\omega_{VF}=frac_{F}*\left( 1-frac_{FSA} \right)$$

$$\omega_{VM}=\left( 1-frac_{F} \right)*(1-frac_{MSA})$$

$$\omega_{C}=\omega_{FFSWinCot}=\omega_{FFSWoutCot}=0$$

$$\sum_{i=1}^{i=9} \omega_{i}=1$$

Where $\omega_{i}$ is the fraction of new individuals entering group *i*; PFSW, LFSW, GPF, GPM, VF, VM, C, FFSWinCot and FFSWoutCot represent professional female sex workers, part-time female sex workers, low risk women and men, not yet sexually active women and men, clients, former female sex workers in GC and former female sex workers outside of GC respectively; $frac_{F}$ is the fraction of individuals that are female; $frac_{FPFSW}$ is the fraction of women that are professional sex workers; $ratio_{LFSW}$ is the ratio of number of part-time sex workers to professional sex workers; $frac_{FSWforeign}$ is the fraction of sex workers that are non-Beninese; $frac_{MSA}$ is the fraction of men that are sexually active; $frac_{FSA}$ is the fraction of women that are sexually active.

#### S2d: Turnover:

Turnover between compartments is a critical component of the model, spreading infections from high risk to low risk groups, and is also important for maintaining the correct demographic patterns in the population. Turnover is represented by two separate parameters, moving out ($u_{i}$) and moving in ($v_{ij}$). The former is the rate of leaving a group *i*, thus need only apply to the group in question. The latter is the rate of entering a group *i* from group *j* which capture the correct prevalences of care-state *r* and stage of disease *s*.

Turnover applies equally to both susceptible and infected states (whereby $X = S$ and $X = I$ respectively), and applies to all care states and stages of infection. Thus, the movement into states $S_{i}^{0}$ and $I_{i}^{4,5}$ will arrive from states $S_{j}^{0}$ and $I_{j}^{4,5}$ respectively. As such, when considering the movement of people from group *j* to group *i*, it will be at the HIV prevalence of group *j*. Note however that professional FSWs on PrEP do not move to their associated PrEP category, but rather into the susceptible off PrEP category ($S_{i}^{0}$) of the destination group.

Movements in and out are combined into the turnover $T_{i}^{rs}(t)$ as follows:

$$T_{i}^{r,s}\left( t \right)=-X_{i}^{r,s}\left( t \right)*u_{i}+\sum_{j=1}^{j=9} v_{ij}*X_{j}^{r,s}(t)$$

Where $T_{i}^{r,s}\left( t \right)$ is the turnover of group *i*, care-state *r* and disease state *s*;$X_{i}^{r,s}\left( t \right)$ is the current number of individuals in the state, $u_{i}$ is the rate of leaving of group *i*; $v_{ij}$ is the rate of entering in group *i* from group *j*.

Groups (*i*):

1. Professional FSW
2. Part-time FSW
3. Low-risk women
4. Former FSW in GC
5. Clients
6. Low-risk men
7. Not yet sexually active women
8. Not yet sexually active men
9. Former FSW outside model

*Not yet sexually active men and women*

Not yet sexually active (NYSA) men and women enter the sexually active population and do not go back. NYSA women (*i* = 7) move into the low-risk women category (*i* = 3); NYSA men (*i* = 8) move into the low-risk men category (*i* = 6).

Rate of leaving NYSA woman group ($u_{7})$:

$$u_{7}=\frac{1}{\chi_{7}-15}$$

Rate of leaving NYSA men group ($u_{8})$:

$$u_{8}=\frac{1}{\chi_{8}-15}$$

Where $u_{7}$ and $u_{8}$ are the rate leaving of NYSA women and men groups respectively; $\chi_{i}$ is the average age of sexual debut of group *i*. 15 is the youngest age in the model, thus $\chi_{8}-15$ represents the length of time spent NYSA.

This results in the movement of NYSA women and men to be expressed as follows:

$$T_{7}^{r,s}(t)=-X_{7}^{r,s}\left( t \right)*u_{7}$$

$$T_{8}^{r,s}(t)=-X_{8}^{r,s}\left( t \right)*u_{8}$$

Where $T_{7}^{r,s}(t)$ and $T_{7}^{r,s}(t)$ are the movements of NYSA women and men respectively of care state *r* and stage of infection *s*; $X_{i}^{r,s}$ is either a susceptible (X = S) or infected (X = I) individual in group *i*, care state *r* and stage of infection *s* at time *t*.

*Professional and part-time female sex workers*

Professional and part-time FSWs are recruited from either outside of sexual network of GC, or recruited from the low-risk population. Both FSW groups retire to former FSW groups (both inside and outside GC).

Rate of leaving sex work is equal to the inverse of the duration of sex work ($\frac{1}{dur_{PFSW}}$ and $\frac{1}{dur_{LFSW}}$ for professional and part-time FSW respectively).

Rate of leaving professional sex work ($u_{1})$:

$$u_{1}=\frac{1}{dur_{PFSW}}$$

Rate of leaving part-time sex work ($u_{2}$):

$$u_{2}=\frac{1}{dur_{LFSW}}$$

Where $dur_{PFSW}$ and $dur_{LFSW}$is the duration of professional and part-time sex work respectively.

The movement rates of FSWs from the low-risk women group are calculated as follows:

Rate of entering professional sex work from low-risk women:

$$v_{1,3}=\left( \frac{1}{dur_{PFSW}}+\mu_{F}+\nu\right) \frac{N_{1}(0)}{N_{3}(0)}\left( 1-frac_{FSWforeign} \right)$$

Rate of entering part-time sex work from low-risk women:

$$v_{2,3}=\left( \frac{1}{dur_{LFSW}}+\mu_{F}+\nu\right) \frac{N_{2}(0)}{N_{3}(0)}\left( 1-frac_{FSWforeign} \right)$$

Where $\mu_{F}$ is female baseline mortality; $\nu$ is the rate of ageing; $N_{i}(0)$ is the initial population size of group *i*; $frac_{FSWforeign}$ is the fraction of sex workers that are non-Beninese. If $frac_{FSWforeign}=0$, all FSWs are recruited from the low-risk women group of GC; if $frac_{FSWforeign}=1$, all FSWs are recruited from outside GC within the rate of entry $E_{1}$ and $E_{2}$ for professional and part-time sex workers respectively. The ratio of professional FSW and part-time FSW to low-risk women ($\frac{N_{1}(0)}{N_{3}(0)}$ and $\frac{N_{2}(0)}{N_{3}(0)}$ respectively) is required to ensure that the approximately the same numbers of FSWs that retire from sex work are recruited from the low-risk women group. $\mu_{F}$ and $\nu$ are also required to recruit FSWs at the rate at which they age and die.

Total movement for professional and part-time FSW ($T_{1}^{r,s}\left( t \right)$ and $T_{2}^{r,s}\left( t \right)$ respectively), including leaving and entering, are thus expressed as follows:

$$T_{1}^{r,s}\left( t \right)=-X_{1}^{r,s}\left( t \right)*u_{1}+v_{1,3}*X_{3}^{r,s}(t)$$

$$T_{2}^{r,s}\left( t \right)=-X_{2}^{r,s}\left( t \right)*u_{2}+v_{2,3}*X_{3}^{r,s}(t)$$

Where $u_{1}$ and $u_{2}$ are rates of leaving professional and part-time sex work respectively, which is multiplied by the number of individuals in each care state *r* and stage of infection *s* at time *t* ($X_{1}^{r,s}\left( t \right)$ and $X_{2}^{r,s}\left( t \right)$ respectively); $v_{1,3}$ and $v_{2,3}$ are the rates of entering professional and part-time sex work respectively, which are multiplied by the number of individuals in the low-risk women ($X_{3}^{r,s}(t)$) of care state *r* and stage of infection *s*.

*Low-risk women*

The rate of leaving of low-risk women group ($u_{3}$) is the combination of the rates $v_{1,3}$ and $v_{2,3}$ as calculated in equations X and X:

$$u_{3}=\left( \frac{1}{dur_{PFSW}}+\mu_{F}+\nu\right) \frac{N_{1}(0)}{N_{3}(0)}\left( 1-frac_{FSWforeign} \right)+\left( \frac{1}{dur_{LFSW}}+\mu_{F}+\nu\right) \frac{N_{2}(0)}{N_{3}(0)}\left( 1-frac_{FSWforeign} \right)$$

The rate of entering of low-risk women group is equal to the rate of leaving of the NYSA women group as shown in the following equation:

$$v_{3,7}=\frac{1}{\chi_{7}-15}$$

Total movement for low-risk womens ($T_{3}^{r,s}\left( t \right)$), including leaving and entering, is expressed as follows:

$$T_{3}^{r,s}\left( t \right)=-X_{3}^{r,s}\left( t \right)*u_{3}+v_{3,7}*X_{7}^{r,s}(t)$$

Where $X_{3}^{r,s}\left( t \right)$ is the number of low-risk womens individuals in care state *r* and stage of infection *s* at time *t*; $u_{3}$ is the leaving rate of low-risk women; $v_{3,7}$ is the entering rate of low-risk women from the NYSA women group; $X_{7}^{r,s}(t)$ is the number of NYSA women in the same care state and stage of infection.

*Former FSWs inside and outside Grand Cotonou*

Professional and part-time FSWs retire to become former FSWs, either inside or outside of GC (as shown in equations X and X). The parameter $frac_{FSWforeign}$ represents the fraction of FSW that were foreign-born, which determines how many FSW that leave GC upon retirement.

Rate of entering former FSW in GC from professional FSW category:

$$v_{4,1}=\frac{1}{dur_{PFSW}} \left( 1-frac_{FSWforeign} \right)$$

Rate of entering former FSW in GC from part-time FSW category:

$$v_{4,2}=\frac{1}{dur_{LFSW}} \left( 1-frac_{FSWforeign} \right)$$

Rate of entering former FSW outside GC from professional FSW category:

$$v_{9,1}=\frac{1}{dur_{PFSW}} frac_{FSWforeign}$$

Rate of entering former FSW outside GC from part-time FSW category:

$$v_{9,2}=\frac{1}{dur_{LFSW}} frac_{FSWforeign}$$

Total movement for former FSW inside ($T_{4}^{r,s}\left( t \right)$) and outside GC ($T_{9}^{r,s}\left( t \right)$) are calculated as follows:

$$T_{4}^{r,s}\left( t \right)=v_{4,1}*X_{1}^{r,s}\left( t \right)+ v_{4,2}*X_{2}^{r,s}(t)$$

$$T_{9}^{r,s}\left( t \right)=v_{9,1}*X_{1}^{r,s}\left( t \right)+ v_{9,2}*X_{2}^{r,s}(t)$$

Where $X_{1}^{r,s}\left( t \right)$ and $X_{2}^{r,s}\left( t \right)$ represent the number of professional and part-time FSWs respectively, in care state *r* and stage of infection *s* at time *t*.

*Clients*

Clients are recruited only from the low-risk men population, at a rate inversely proportional to the duration of being a client ($\frac{1}{dur_{client}}$).

Rate of leaving client group ($u_{5}$):

$$u_{5}=\frac{1}{dur_{client}}$$

In order to keep the proportion of men that are clients approximately the same over time, the rate of entering the client group from the low-risk men group ($v_{5,6}$) scales the rate of leaving of the client group by the fraction of men that are clients at seeding ($\frac{N_{5}(0)}{N_{6}(0)}$). The background male mortality and ageing rates ($\mu_{M}$ and $\nu$ respectively) are included to replace the clients that are leaving due to death or ageing. The rate of entering the client category from the low-risk men ($v_{5,6}$) is calculated as follows:

$$v_{5,6}=\left( \frac{1}{dur_{client}}+\mu_{M}+\nu\right)\frac{N_{5}(0)}{N_{6}(0)}$$

Total movement for clients is thus calculated as follows:

$$T_{5}^{r,s}\left( t \right)=-X_{5}^{r,s}\left( t \right)*u_{5}+v_{5,6}*X_{6}^{r,s}(t)$$

Where $X_{6}^{r,s}(t)$ is the number of low-risk men in care state *r* and stage of infection *s* at time *t*.

*Low-risk men*

Low-risk men leave at rate $u_{6}$, which is calculated as follows, as in the following equation:

$$u_{6}=\left( \frac{1}{dur_{client}}+\mu_{M}+\nu\right)\frac{N_{5}(0)}{N_{6}(0)}$$

Rate of entering low-risk men from the client category ($v_{6,5}$) is calculated as follows:

$$v_{6,5}=\frac{1}{dur_{client}}$$

Rate of entering low-risk men from the NYSA men category ($v_{6,8}$) is calculated as follows:

$$v_{6,8}=\frac{1}{\chi_{8}-15}$$

Where $\chi_{8}$ is the average age of sexual debut of NYSA men.

Total movement for the low-risk men category ($T_{6}^{r,s}\left( t \right)$) combines the turnover with clients with the movement from NYSA men, and is calculated as follows:

$$T_{6}^{r,s}\left( t \right)=-X_{6}^{r,s}\left( t \right)*u_{6}+v_{6,5}*X_{5}^{r,s}(t) +v_{6,8}*X_{8}^{r,s}(t)$$

Where $X_{5}^{r,s}(t)$ and $X_{8}^{r,s}(t)$ are the numbers of clients and NYSA men respectively, in care state *r* and stage of infection *s*.

#### S2e: Testing rates

Household, IBBA and DHS surveys provided estimates on the probability of being tested in the last year for FSWs and men and women in GC. After interpolating over time yearly estimates of testing coverage (see parameter table), every time step we estimate the rate of undergoing testing with the following equation:

$$\tau_{i}^{r,s}=-ln(1-ptest_{i})$$

Where$\tau_{i}^{r,s}$ is the rate of testing of group *i*; $ptest_{i}$ is the probability of being tested last year of group *i*.

There are two cases when this is different:

1. Individuals in $I_{i}^{0,5}$ have an increased testing rate ($\tau_{i}^{0,5}$) as calculated below

$$\tau_{i}^{0,5}=-\ln\left( 1-ptest_{i} \right)*RR_{testCD4200}$$

Where $RR_{testCD4200}$ is the increase in testing due to a CD4 count <200, which is associated with symptoms of AIDS.

2. For individuals on PrEP but infected in primary phase ($I^{1,1}$), testing rate $\tau_{i}^{1,1}$ is fixed to 4 (year^-1^), as those under PrEP surveillance are tested every 3 months.

$$\tau_{i}^{1,1}=4$$

We assume no difference in test sensitivity by PrEP status or stage of infection.

Furthermore, increased testing in the TasP/PrEP intervention is reflected in increased testing when simulating the intervention scenario. This additional testing rate, represented by $\eta_{i}\left( t \right)$, is only applied during the 2 years of the study, or for longer when simulating an intervention scale up.

#### S2f: ART initiation rates

ART initiation rate is represented by $\rho_{i}^{s}$, varying by group *i* and stage of infection *s*. Eligibility of ART has historically depended on CD4 count according to national and WHO recommendations; the rates of ART initiation in the model reflect this. Prior to 2012, only those with a CD4 count below 350 were eligible for ART (s=4,5); in 2015 this was extended to those below 500 (s=3,4,5), and in 2016 all stages of infection were eligible.

Increased ART initation due to the TasP/PrEP intervention is also represented in the model, with the parameter $ῤ_{i}\left( t \right)$, which applies only to professional FSWs.

The assumed proportion of all pFSW on ART that are virally suppressed was a weighted average of observed viral suppression among pFSW on TasP (87%), and the assumed viral suppression among pFSW outside of TasP (75%).

#### S2g: Force of infection:

The force of infection ($\lambda_{\mathrm{ij}}^{r}\left( t \right)$) is defined as the per capita rate at which susceptible individuals of risk group *i* and care state *r* are infected by individuals of risk group *j*. It is derived from the total risk of infection from commercial (C) and non-commercial partnerships (N), all care states *r* and stages of infection *s*, including sex acts with and without protection from condoms and PrEP.

$$\lambda_{ij}^{r}\left( t \right)=p_{ij}^{C} c_{i}^{C}\sum_{r=0}^{r=4} \sum_{s=1}^{s=5} \left( \frac{I_{j}^{r,s}\left( t \right)}{\sum_{r=0}^{r=4} \sum_{s=1}^{s=5} \left( I_{j}^{r,s}\left( t \right)+S_{j}^{r,s}\left( t \right) \right)}\left( 1-\left( 1-\left( 1-e_{P}^{r} \right) \beta_{ij} R^{r,s} \right)^{n_{ij}^{C}\left( t \right) \left( 1-f_{ij}^{C}\left( t \right) \right)}\times\left( 1-\left( 1-{ec}_{ij} \right) \left( 1-e_{P}^{r} \right) \beta_{ij} R_{j}^{r,s} \right)^{n_{ij}^{C}\left( t \right) f_{ij}^{C}\left( t \right)} \right) \right)+p_{ij}^{N} c_{i}^{N}\sum_{r=0}^{r=4} \sum_{s=1}^{s=5} \left( \frac{I_{j}^{r,s}\left( t \right)}{\sum_{r=0}^{r=4} \sum_{s=1}^{s=5} \left( I_{j}^{r,s}\left( t \right)+S_{j}^{r,s}\left( t \right) \right)}\left( 1-\left( 1-\left( 1-e_{P}^{r} \right) \beta_{ij} R^{r,s} \right)^{n_{ij}^{N}\left( t \right) \left( 1-f_{ij}^{N}\left( t \right) \right)}\times\left( 1-\left( 1-{ec}_{ij} \right) \left( 1-e_{P}^{r} \right) \beta_{ij} R_{j}^{r,s} \right)^{n_{ij}^{N}\left( t \right) f_{ij}^{N}\left( t \right)} \right) \right)$$

Where $p_{ij}^{C}$ and $p_{ij}^{N}$ are the probability of sexual contact between groups *i* and *j* for commercial and non-commercial partnerships respectively; $c_{i}^{C}$ and $c_{i}^{N}$ are the rates of new partner acquisition for commercial and non-commercial partnerships respectively; $\beta_{ij}$ and $\beta_{ij}$ are the commercial and non-commercial probability of HIV transmission per sex act from *j* to *i* respectively; $R_{j}^{r,s}$ is a matrix containing the scaling factors for risk of transmission based on care state *r* and stage of infection *s* of transmitter *j*; $n_{ij}^{C}\left( t \right)$ and $n_{ij}^{N}\left( t \right)$ are the commercial and non-commercial number of sex acts per partnership, $f_{ij}^{C}\left( t \right)$ and $f_{ij}^{N}\left( t \right)$ are the proportion of commercial and non-commercial sex acts protected by a condom, ${ec}_{ij}$ is the condom efficacy, and $e_{P}^{r}$ is PrEP efficacy of care state *r* (perfect, intermediate or non-adherence to PrEP). For individuals who are not using PrEP, or who are not adherent, efficacy of PrEP is assumed to be 0.

The total force of infection experienced by individuals of group *i*, and care state *r* $\left( {\lambda tot}_{i}^{r}\left( t \right) \right)$is the sum of the forces of infection contributed by each type of partnership they have.

$${\lambda tot}_{i}^{r}\left( t \right)=\sum_{j} \lambda_{\mathrm{ij}}^{r}\left( t \right)$$

#### S2h: Balancing of partnerships

*Commercial partnerships*

The total number of commercial partnerships as declared by women (*i* = 1, 2) must match the total number of commercial partnerships as declared by men (*i* = 5), such that the following equation is satisfied:

$$c_{1}^{C}N_{1}+c_{2}^{C}N_{2}=c_{5}^{C}N_{5}$$

Where $c_{i}^{C}$ is the commercial partner change rate of group *i*; $N_{i}$ is the total number of individuals in group *i*. We use two methods (one of which is randomly chosen in each simulation) in order to ensure that this equation is fulfilled:

1. Either we change the partner change rate of the professional FSWs every timestep in order to balance

$$c_{1}^{C'}=\frac{c_{5}^{C}N_{5}-c_{2}^{C}N_{2}}{N_{1}}$$

Where $c_{1}^{C'}$ is the balanced partner change rate of professional FSWs, which will be applied in the force of infection.

2. Or we change the partner change rate of the clients every timestep in order to balance

$$c_{5}^{C'}=\frac{c_{1}^{C}N_{1}+ c_{2}^{C}N_{2}}{N_{5}}$$

Where $c_{5}^{C'}$ is the balanced partner change rate of clients, which will be applied in the force of infection.

*Non-commercial partnerships*

The number of non-commercial partnerships as declared by women (*i* = 1, 2, 3, 4) must match the number of partnerships as declared by men (*i* = 5, 6), such that the following equation is satisfied:

$$\sum_{i=1}^{i=4} c_{i}^{N}N_{i}=\sum_{i=5}^{i=6} c_{i}^{N}N_{i}$$

$$c_{1}^{N}N_{1}+c_{2}^{N}N_{2}+c_{3}^{N}N_{3}+c_{4}^{N}N_{4}=c_{5}^{N}N_{5}+c_{6}^{N}N_{6}$$

We change the partner change rate of low-risk women (i=3) and former FSW (i=4) in GC every timestep to balance partnerships:

$$c_{3}^{N'}=c_{4}^{N'}=\frac{c_{5}^{N}N_{5}+c_{6}^{N}N_{6}-c_{1}^{N}N_{1}-c_{2}^{N}N_{2}}{N_{3}+N_{4}}$$

Low-risk women and former FSW in GC share the same non-commercial partner change rate ($c_{3}^{N'}$ and $c_{4}^{N'}$ respectively).

*Sex acts per partnership*

This parameter is equal between each pairing, i.e. professional FSW have the same number of sex acts per commercial partnerships with clients, as clients have with professional FSW. The value of this parameter is drawn from a range whose uncertainty encompasses estimates from professional FSW and client data.

#### S2i: Probability of sexual contact

The contact matrices $M_{ij}^{C}$ and $M_{ij}^{N}$ determine whether commercial and non-commercial partnerships are formed between groups *i* and *j*.

$$M_{ij}^{C}=$$

|  | | j | | | | | | | | |
| --- | --- | --- | --- | --- | --- | --- | --- | --- | --- | --- |
|  |  | Pro FSW | Part-time FSW | Low-risk women | Former FSW in Cotonou | Client | Low-risk men | NYSA women | NYSA men | Former FSW not in Cotonou |
| i | Pro FSW | 0 | 0 | 0 | 0 | 1 | 0 | 0 | 0 | 0 |
|  | Part-time FSW | 0 | 0 | 0 | 0 | 1 | 0 | 0 | 0 | 0 |
|  | Low-risk women | 0 | 0 | 0 | 0 | 0 | 0 | 0 | 0 | 0 |
|  | Former FSW in Cot | 0 | 0 | 0 | 0 | 0 | 0 | 0 | 0 | 0 |
|  | Client | 1 | 1 | 0 | 0 | 0 | 0 | 0 | 0 | 0 |
|  | Low-risk men | 0 | 0 | 0 | 0 | 0 | 0 | 0 | 0 | 0 |
|  | NYSA women | 0 | 0 | 0 | 0 | 0 | 0 | 0 | 0 | 0 |
|  | NYSA men | 0 | 0 | 0 | 0 | 0 | 0 | 0 | 0 | 0 |
|  | Former FSW not in Cot | 0 | 0 | 0 | 0 | 0 | 0 | 0 | 0 | 0 |

$$M_{ij}^{N}=$$

|  | | j | | | | | | | | |
| --- | --- | --- | --- | --- | --- | --- | --- | --- | --- | --- |
|  |  | Pro FSW | Part-time FSW | Low-risk women | Former FSW in Cotonou | Client | Low-risk men | NYSA women | NYSA men | Former FSW not in Cotonou |
| i | Pro FSW | 0 | 0 | 0 | 0 | 1 | 0 | 0 | 0 | 0 |
|  | Part-time FSW | 0 | 0 | 0 | 0 | 1 | 0 | 0 | 0 | 0 |
|  | Low-risk women | 0 | 0 | 0 | 0 | 1 | 1 | 0 | 0 | 0 |
|  | Former FSW in Cot | 0 | 0 | 0 | 0 | 1 | 1 | 0 | 0 | 0 |
|  | Client | 1 | 1 | 1 | 1 | 0 | 0 | 0 | 0 | 0 |
|  | Low-risk men | 0 | 0 | 1 | 1 | 0 | 0 | 0 | 0 | 0 |
|  | NYSA women | 0 | 0 | 0 | 0 | 0 | 0 | 0 | 0 | 0 |
|  | NYSA men | 0 | 0 | 0 | 0 | 0 | 0 | 0 | 0 | 0 |
|  | Former FSW not in Cot | 0 | 0 | 0 | 0 | 0 | 0 | 0 | 0 | 0 |

The probabilities of commercial and non-commercial sexual contact of an individual from group *i* with an individual from group *j* ($p_{ij}^{C}$ and $p_{ij}^{C}$ respectively) is determined by the proportion of partnerships declared by group *j* out of the total number of partnerships, calculated as follows:

$$p_{ij}^{C}\left( t \right)=\left\{ \begin{aligned} \frac{c_{j}^{C}N_{j}}{\sum_{j=1}^{9} c_{j}^{C} N_{j}M_{ij}^{C}}, M_{ij}^{C}=1 \\ 0, M_{ij}^{C}=0 \end{aligned} \right.$$

$$p_{ij}^{N}\left( t \right)=\left\{ \begin{aligned} \frac{c_{j}^{N}N_{j}}{\sum_{j=1}^{9} c_{j}^{N} N_{j}M_{ij}^{N}}, M_{ij}^{N}=1 \\ 0, M_{ij}^{N}=0 \end{aligned} \right.$$

Where $c_{j}^{C}$ and $c_{j}^{N}$ represent the commercial and non-commercial partner change rates of group *j* respectively, *after balancing.*

#### S2j: HIV transmission probability per sex act

From Boily et al. 2009, we obtain ranges for the baseline probability of HIV transmission from men to women ($\beta$) ^18^. We estimate the per sex act probability of transmission from men to women (and vice versa) which depends on: relative risk of HIV transmission probability (RR) with concurrent HSV-2 infection in the acquirer, female-to-male transmission, circumcision, and HSV-2 prevalence in the acquirer. We assume all males are circumcised ^1,2,5^, which confers lower risk of HIV acquisition in men but not transmission from men.

*HIV transmission probability per sex act from men to women (*$\beta_{MF}$*):*

$$\beta_{MF}=\beta*(1+(RR_{j}^{HSV2}-1)*\pi_{j}^{HSV2})$$

Where $RR_{j}^{HSV2}$ is the RR of transmission probability if acquirer group *j* is HSV-2 infected; $\pi_{j}^{HSV2}$ is the prevalence of HSV-2 in group *j*.

*HIV transmission probability per sex act from women to men (*$\beta_{FM}$*):*

$$\beta_{FM}=\beta*RR^{\beta FM}* RR^{\beta circum}*\left( 1+(RR_{j}^{HSV2}-1)*\pi_{j}^{HSV2} \right)$$

Where $RR^{\beta FM}$ is the RR of female-to-male transmission; $RR^{\beta circum}$ is RR of male circumcision.

#### S2k: Relative risk of transmission

Transmission probability per sex act will also depend on the care state (*r*) and the stage of infection (*s*) of the transmitter.

| Relative risk of transmission ($R_{j}^{r,s}$) | | Stage of infection (s) of the transmitter (j) | | | | |
| --- | --- | --- | --- | --- | --- | --- |
|  |  | 1. Primary | 2. CD4 >500 | 3. CD4 350 - 500 | 4. CD4 200 - 349 | 5. CD4 <200 |
| Care state (r) of the transmitter (j) | 0. Off PrEP | RR^primary^ | 1 | 1 | 1 | RR^AIDS^ |
|  | 1. On PrEP | RR^primary^ | - | - | - | - |
|  | 2. Diagnosed off ART | - | 1 | 1 | 1 | RR^AIDS^ |
|  | 3. On ART | - | RR_j_^ART^ | RR_j_^ART^ | RR_j_^ART^ | RR_j_^ART^ |
|  | 4. ART Drop out | - | 1 | 1 | 1 | RR^AIDS^ |

Where $RR^{primary}$ is the relative risk of infection per sex act if the transmitter is in primary stage of infection; RR^AIDS^ is the relative risk of infection	per sex act if the transmitter is in AIDS stage; RR^ART^ is the relative risk of infection if the transmitter is on ART. RR_j_^ART^ is calculated as follows:

$$\mathrm{RR}_{j}^{ART}=1-VS_{j}*eff^{ART}$$

Where $VS_{j}$ is the proportion of individuals that are virally suppressed in group *j,* and $eff^{ART}$ is the efficacy of ART when virally suppressed.

#### S2l: Disease progression and AIDS-related mortality

Progression through the stages of infection occurs at rate $\gamma^{r,s}$ for each care state *r* and current stage *s*.

The duration of the entire infection ($dur_{SCdeath}$, seroconversion to death) is estimated, as well as the duration of primary phase ($\frac{1}{\gamma^{01}}$), infection stage CD4 200-349 ($\frac{1}{\gamma^{04}}$) and CD4 <200 ($\frac{1}{\alpha^{05}}$). The durations of stages CD4 >500 ($\frac{1}{\gamma^{02}}$) and CD4 350-500 ($\frac{1}{\gamma^{03}}$) are calculated as follows:

$$\frac{1}{\gamma^{02}}=\frac{1}{\gamma^{03}}=dur_{SCdeath}-\frac{1}{\gamma^{01}}-\frac{1}{\gamma^{04}}-\frac{1}{\alpha^{05}}$$

Progression happens at the same rate when undiagnosed off PrEP, undiagnosed on PrEP, diagnosed off PrEP or in ART dropout (r = 0, 1, 2, 4); progression is slowed by a factor of ${prog}_{ART}$ for those on ART (r = 3) such that:

$$\gamma^{32}=\frac{\alpha^{03}}{{prog}_{ART}}*VS_{j}+ \gamma^{02}\left( 1-VS_{j} \right)$$

$$\gamma^{33}=\frac{\alpha^{04}}{{prog}_{ART}}*VS_{j}+ \gamma^{03}\left( 1-VS_{j} \right)$$

$$\gamma^{34}=\frac{\alpha^{05}}{{prog}_{ART}}*VS_{j}+ \gamma^{04}\left( 1-VS_{j} \right)$$

Where $VS_{j}$ is the proportion of individuals that are virally suppressed in group *j.*

If an individual on PrEP is infected (stage $I_{i}^{1,1}$), they progress at the same rate as if not on PrEP:

$$\gamma^{1,1}=\gamma^{0,1}$$

There is no AIDS-related mortality for those who are susceptible, primary phase of infection, or CD4 >500 (s = 0, 1, 2). It occurs for CD4 350-500, CD4 200-249, CD4 <200 (s = 3, 4, 5). AIDS related mortality for those on ART (r = 3) is slowed by a factor of ${mort}_{ART}$, such that:

$$\alpha^{33}=\frac{\alpha^{03}}{{mort}_{ART}}*VS_{j}+ \alpha^{03}\left( 1-VS_{j} \right)$$

$$\alpha^{34}=\frac{\alpha^{04}}{{mort}_{ART}}*VS_{j}+ \alpha^{04}\left( 1-VS_{j} \right)$$

$$\alpha^{35}=\frac{\alpha^{05}}{{mort}_{ART}}*VS_{j}+ \alpha^{05}\left( 1-VS_{j} \right)$$

### Section 3: Outcomes

Outcomes estimated for each scenario to evaluate impact:

| **Statistic** | **Calculation** |
| --- | --- |
| Cumulative HIV infections (${CI}_{i}$) | Cumulative HIV infections (${CI}_{i}$) are calculated as follows:  $\frac{d{CI}_{i}}{dt}=\sum_{s=0}^{s=1d} S_{i}^{s}(t)\sum_{j=1}^{j=9} \lambda_{ij}^{s}(t)$  Where $S_{i}^{s}(t)$ represents susceptible inidividuals in group $i$ and PrEP category $s$ at time $t$, and $\lambda_{ij}^{s}(t)$ represents the force of infection from group $j$ to group $i$ in PrEP category s at time $t$. |
| HIV-related deaths ($H_{i}$) | Cumulative HIV-related deaths ($H_{i}$) are calculated as follows:  $\frac{dH_{i}}{dt}=\sum_{r=0}^{r=4} \sum_{s=1}^{s=5} \alpha_{i}^{r,s} I_{i}^{r,s}(t)$  Where $I_{i}^{r,s}(t)$ represents infected individuals in group i, in care-state r, stage of infection s at time t, and $\alpha_{i}^{r,s}$ represents the AIDS-related mortality rate in group i, care state r, stage of infection s. |
| Life years ($LY_{i}$) | Life years ($LY_{i}$) is calculated by summing the number of individuals in each group ($N_{i}(t)$) at mid-year points each year:  $LY_{i}= \sum_{t=t1}^{t=t2} N_{i}(t)$ |
| Disability-adjusted life years ($DALY_{i}$) | DALYs are calculated by weighting the life years of each disease state by quality of life. Susceptible individuals have a “normal” life year attributed, while those in more diseased states (lower CD4 count) are attributed smaller DALY weights.  D^0^: Susceptible  D^1^: Any of the following: primary phase, CD4 > 500, CD4 350-500, on ART  D^2^: CD4 200-349 NOT on ART  D^3^: CD4 < 200 NOT on ART  D^0^, D^1^, D^2^, D^3^ are attributed DALY weights W^0^, W^1^, W^2^, W^3^ respectively  $DALY_{i}= \sum_{t=t1}^{t=t2} \left( D_{i}^{0}\left( t \right)*W^{0}+D_{i}^{1}\left( t \right)*W^{1}+D_{i}^{2}\left( t \right)*W^{2}+D_{i}^{3}\left( t \right)*W^{3} \right)$ |

**Estimating intervention impact**

The following outcomes were estimated for each scenario: cumulative infections ($CI$), life years ($LY$) and disability adjusted life years ($DALY$) as calculated in Table 3.

Infections averted ($IA_{i}^{W}$) was calculated as the relative difference in cumulative infections between intervention $w$ ($CI_{i}^{W}$) and the counterfactual scenario (Scenario C0, Table 1) ($CI_{i}^{C}$).

$$IA_{i}^{W}=CI_{i}^{W}-CI_{i}^{C}$$

Life years gained and DALYs averted were calculated in a similar fashion as above.

The fraction of infections averted ($fIA_{w})$ was calculated as the proportion of cumulative infections in the counterfactual that were averted by intervention w:

$$fIA_{i}^{W}=\frac{IA_{i}^{W}}{CI_{i}^{C}}$$

Impact was calculated among professional FSW only ($i=1$), clients ($i=5$) and in the total population of Grand Cotonou (e.g. $\sum_{i=1}^{i=8} IA_{i}^{W}$), measured over 2 year (2015-2017) and 20 year (2015-2035) time horizons; the latter will allow observation of long term impact such as accumulated life-years and prevention of secondary HIV transmissions.

### Section 4: Parameters

Table 1 below displays full details of the parameters in the model: demographic, sexual behaviour, biological, intervention, PrEP and TasP. Some parameters were sampled in a Latin Hypercube from a uniform distribution of the range given in the table, others were fixed and not sampled. Some parameter values were sampled at certain timepoints (e.g. commercial partner change range); we assume that parameter values between sampled timepoints are linearly interpolated between them.

Model parameters are derived from several sources. Where there were multiple sources estimating the same parameter, a prior range was defined encompassing all estimates, and sampled in the Latin Hypercube. Parameter estimates sourced from the published literature assumed the 95% CI as the prior range.

#### Table S1A: Demographic parameters

| **Name (units)** | **Risk group** | **Year** | **Symbol** | **Value/Range** | **Source** |
| --- | --- | --- | --- | --- | --- |
| Year of seeding of epidemic | NA | NA | t_0_ | 1986 | ^19^ |
| Total population aged 15-59 at seed | All | NA | N_0_ | 286114 | ^20,21^ |
| Population growth rate (per year) | All | 1979-1992 | ε | 0.059 | ^20-23^ |
|  | All | 1993-2002 |  | 0.048 - 0.058 |  |
|  | All | 2002-2013 |  | 0.027 |  |
|  | All | 2014- |  | 0.027 |  |
| Proportion of population that is female at seeding | NA | 1986-2035 | prop_F_ | 0.512 - 0.520 | ^20-23^ |
| Proportion of women that are professional FSW at seeding | Pro FSW | 1986-2035 | prop_PFSW_ | 0.0024 - 0.00715 | ^20-25^ |
| Ratio of women that are part-time FSW to Pro FSW at seeding | Part-time FSW | 1986-2035 | ratio_LFSW_ | 1 - 2 | ^25^ |
| Proportion of women that are former FSW at seeding | Former FSW in Cotonou | 1986-2035 | prop_FFSW_ | Same as proportion that are professional FSW | Assumption |
| Proportion of men that are clients at seeding | Client | 1986-2035 | prop_C_ | 0.066 - 0.30 | ^20-23,26,27^ |
| Fraction of the 15-59 population that is not yet sexually active at seeding | Low-risk women | 1986-2035 | prop_FV_ | 0.079 - 0.20 | ^2,5^ |
|  | Low-risk men | 1986-2035 | prop_MV_ | 0.070 - 0.17 | ^2,5^ |
| Prevalence of HIV at seeding | FSW | NA | π_FSW_ | 0.0132 - 0.0659 | ^19^ |
|  | Client | NA | π_C_ | 0.000313 - 0.00294 | ^28^ |
|  | Low-risk women | NA | π_F_ |  |  |
|  | Low-risk men | NA | π_M_ |  |  |
| Rate of leaving by ageing (per year) | All | 1986-2035 | ν | 0.022 | Assumption |
| Background mortality rate (per year) | Women | 1986-2035 | μ | 0.0187 - 0.0200 | ^29^ |
|  | Men |  |  | 0.0194 - 0.0220 |  |
| Rate of leaving of sex work (per year) | Pro FSW & part-time FSW | 1986-2035 | 1 / dur_FSW_ | 0 - 0.55 | ^3,4,9,11^ |
| Proportion of FSW that are non-Beninese | Pro FSW | 1986-2035 | frac_FSWforeign_ | 0.5 - 0.9 | ^3,4,9,11^ |
| Prevalence of incoming professional FSWs | Pro FSW | 1986 - 1993 |  | 0 - 0.163 | ^3,4,9,11,30^ |
|  |  | 1986 - 2015 |  | Linearly interpolated |  |
|  |  | 2015 - |  | 0 - 0.046 |  |
| Rate of leaving of client category (per year) | Client | 1986-2035 | 1 / dur_client_ | 0 - 0.295 | ^8,10,12^ |
| Rate of entering the sexual population (per year) | Not yet sexually active female | 1986-2035 | χ_F_ | 0.2 - 0.5 | ^2,5^ |
|  | Not yet sexually active male | 1986-2035 | χ_M_ | 0.2 - 0.5 | ^2,5^ |
| Proportion of people that are sexually active at 15 | Women | 1986-2035 | frac_WSA_ | 0.12 – 0.17 | ^2,5^ |
|  | Men | 1986-2035 | frac_MSA_ | 0.18 – 0.35 | ^2,5^ |

#### Table S1B: Sexual behaviour parameters

| **Name** | **Group** | **Year** | **Symbol** | **Value** | **Source** |
| --- | --- | --- | --- | --- | --- |
| Commercial partner change rate (per year) | Professional FSW | 1986 - 1993 | c^C^_PFSW_ | 192 - 1277 | ^3,4,9,11^ |
|  |  | 1993 - 2005 |  | Linearly interpolated |  |
|  |  | 2005 - |  | 81 - 562 |  |
|  | Part-time FSW | 1986-2035 | c^C^_LFSW_ | 26 - 78 | Personal communication |
|  | Client | 1986 - 1998 | c^C^_C_ | 8.4 - 32 | ^8,10,12^ |
|  |  | 1998 - 2002 |  | Linearly interpolated |  |
|  |  | 2002 - |  | 11.1 - 19.8 |  |
| Non-commercial partner change rate (per year) | Professional FSW | 1986-2035 | c^N^_PFSW_ | 0.31 – 0.86 | ^3,4,9,11^ |
|  | Part-time FSW | 1986-2035 | c^N^_LFSW_ | 0.41 – 1.04 | ^9,11^ |
|  | Client | 1986-2035 | c^N^_C_ | 1.6 – 3.3 | ^8,10,12^ |
|  | GPF | 1986 - 1998 | c^N^_F_ | 0.93 - 0.99 | ^2,5^ |
|  |  | 1998 - 2008 |  | Linearly interpolated |  |
|  |  | 2008 - |  | 0.77 - 0.82 |  |
|  | GPM | 1986 - 1998 | c^N^_M_ | 1.25 - 1.43 | ^2,5^ |
|  |  | 1998 - 2008 |  | Linearly interpolated |  |
|  |  | 2008 - |  | 0.73 - 0.84 |  |
| Commercial number of sex acts per partnership (per year) | Professional FSW | 1986-2035 | n^C^_PFSW_ | 1 - 3 | ^3,4,9,11^ |
|  | Part-time FSW | 1986-2035 | n^C^_LFSW_ | 1 | Assumption |
| Non-commercial number of sex acts per partnership (per year) | Professional and part-time FSW | 1986 - 2002 | n^N^_PFSW_ | 13.0 - 20.0 | ^3,4,9,11^ |
|  |  | 2002 - 2015 |  | Linearly interpolated |  |
|  |  | 2015 - |  | 38.2 - 60.0 |  |
|  | Clients | NA | n^N^_C_ | Matching the partner in their respective partnerships | Assumption |
|  | GPF - GPM | 1998 | n^N^_F_ | 35 - 44 | ^2,5^ |
|  |  | 1998 - 2011 |  | Linearly interpolated |  |
|  |  | 2011 |  | 29 - 38 |  |
| Proportion of commercial sex acts protected by condom | Professional FSW & clients | 1986 | fc^C^_PFSWclient_ | 0 - 0.18 | ^3,4,8,10,12^ |
|  |  | 1986 - 1993 |  | Linearly interpolated |  |
|  |  | 1993 |  | 0.18 - 0.33 |  |
|  |  | 1993 - 1998 |  | Linearly interpolated |  |
|  |  | 1998 |  | 0.4 - 0.73 |  |
|  |  | 1998 - 2002 |  | Linearly interpolated |  |
|  |  | 2002 |  | 0.61 - 0.99 |  |
|  |  | 2002 - 2008 |  | Linearly interpolated |  |
|  |  | 2008 |  | 0.86 - 0.99 |  |
|  | Part-time FSW & clients | 1986 | fc^C^_LFSWclient_ | 0 | Assumption |
|  |  | 1986 - 2015 |  | Linearly interpolated |  |
|  |  | 2015 |  | 0.25 – 0.52 | ^8-12^ |
| Proportion of non-commercial sex acts protected by condom | Professional FSW & clients | 1986 | fc^N^_FSW_ | 0 | ^3,4,8,10,12^ |
|  |  | 1986 - 2002 |  | Linearly interpolated |  |
|  |  | 2002 - |  | 0.19 - 0.62 |  |
|  | Part-time FSW & clients | 1986 | fc^C^_LFSWclient_ | 0 | Assumption |
|  |  | 1986 - 2015 |  | Linearly interpolated |  |
|  |  | 2015 |  | 0.138 – 0.383 | ^8-12^ |
|  | Clients | 1986-2035 | fc^N^_C_ | Same as FSW or GPF in respective partnerships | Assumption |
|  | GPF | 1986 - 1998 |  | 0.033 - 0.05 | ^2,5^ |
|  |  | 1998 - 2011 |  | Linearly interpolated |  |
|  |  | 2011 - |  | 0.16 - 0.26 |  |
|  | GPM | Same as GPF | fc^N^_M_ | Same as GPF |  |

#### Table S1C: Biological parameters

| **Name** | **Group** | **Year** | **Symbol** | **Value** | **Source** |
| --- | --- | --- | --- | --- | --- |
| HIV baseline transmission probability male to female per sex act | All | 1986-2035 | β | 0.0006 - 0.00109 | ^18^ |
| RR HIV transmission risk female to male | All | 1986-2035 | RR^βFM^ | 0.53 - 2 | ^18^ |
| RR transmission probability with HSV-2 infection in acquirer commercial | Professional FSW | 1986-2035 | RRj^HSV2^ | 0.9 – 2.3 | ^31^ |
| RR transmission probability with HSV-2 infection in acquirer commercial | Women who are not professional FSW | 1986-2035 | RRj^HSV2^ | 1.8 – 3.4 |  |
| RR transmission probability with HSV-2 infection in acquirer commercial | Clients | 1986-2035 | RRj^HSV2^ | 1.5 – 2.2 |  |
| RR transmission probability with HSV-2 infection in acquirer commercial | Men who are not clients | 1986-2035 | RRj^HSV2^ | 2.2 – 4.3 |  |
| Prevalence of HSV-2 | Pro FSW & part-time FSW | 1986-2035 | πj^HSV2^ | 0.87 - 0.94 | ^26^ |
| Prevalence of HSV-2 | Clients | 1986-2035 | πj^HSV2^ | 0.18 - 0.28 | ^32^ |
| Prevalence of HSV-2 | GPF | 1986-2035 | πj^HSV2^ | 0.27 - 0.32 | ^26^ |
| Prevalence of HSV-2 | GPM | 1986-2035 | πj^HSV2^ | 0.098 - 0.14 | ^26^ |
| RR transmission circumcision | All | 1986-2035 | RR^βcircum^ | 0.34 - 0.72 | ^33,34^ |
| RR of transmission during primary phase | All | 1986-2035 | RR^primary^ | 4.5 - 18.8 | ^18^ |
| RR of transmission during AIDS (CD4 <200) phase | All | 1986-2035 | RR^AIDS^ | 4.5 - 11.9 | ^18^ |
| Efficacy of ART if virally suppressed regarding reduction in onward transmission | All | 1986-2035 | eff^ART^ | 0.96 - 0.99 | ^35^ |
| Fraction of other groups virally suppressed | Low-risk | 1986-2035 | VS_2-9_ | 0.42 - 0.85 | ^36,37^ |
| Fraction of professional FSW virally suppressed | Professional FSW | 1986 - 2015 | VS_1_ |  |  |
| Condom efficacy | All | 1986-2035 | ec | 0.58-0.95 | ^38^ |
| Duration between seroconversion and death (years) | All | 1986-2035 | dur_SCdeath_ | 8.7 - 12.3 | ^39-46^ |
| Duration of primary phase (years) | All | 1986-2035 | 1/γ_01_ | 0.25 - 0.42 | ^18,40^ |
| Duration of CD4 200-349 (years) | All | 1986-2035 | 1/γ_04_ | 2.3 - 4.4 | ^41,43,47^ |
| Duration in CD4 <200 (years) | All | 1986-2035 | 1/α_05_ | 0.58 - 3.17 | ^41,42,46,48-51^ |
| Duration of CD4 >500 (years) | All | 1986-2035 | 1/γ_02_ | Derived from other sampled parameters | Derived: (dur_SCdeath_ - γ_01_ - γ_04_ - α_05_) gives the time after primary phase until CD4 200-349. Dividing this time by 2 gives the duration of both CD4 >500 and CD4 350-500, assuming they have the same duration |
| Duration of CD4 350-500 (years) | All | 1986-2035 | 1/γ_03_ |  |  |
| Death rate CD4 350-500 (per year) | All | 1986-2035 | α_03_ | 0.01 - 0.05 | ^29,48^ |
| Death rate CD4 200-349 (per year) | All | 1986-2035 | α_04_ | 0.03 - 0.10 |  |
| Reduction in progression for individuals on ART | All | 1986-2035 | prog_ART_ | 4.82 - 10.2 | ^52^ |
| Reduction in mortality for individuals on ART | All | 1986-2035 | mort_ART_ |  |  |

#### Table S1D: Intervention parameters

| **Name** | **Group** | **Year** | **Symbol** | **Value** | **Source** |
| --- | --- | --- | --- | --- | --- |
| Probability of having been tested in last year* | Professional FSW | 1986 - 2005 | τ | 0 | Personal communication |
|  |  | 2005 - 2013 |  | Linearly interpolated |  |
|  |  | 2013 |  | 0.65 | ^53^ |
|  |  | 2013 - 2015 |  | Linearly interpolated |  |
|  |  | 2015 - |  | 0.68 | ^11^ |
|  | Low-risk women | 1986 - 2001 |  | 0 | Personal communication |
|  |  | 2001 - 2006 |  | Linearly interpolated |  |
|  |  | 2006 |  | 0.14 | ^6,7^ |
|  |  | 2006 - 2008 |  | Linearly interpolated |  |
|  |  | 2008 |  | 0.21 | ^5^ |
|  |  | 2008 - 2011 |  | Linearly interpolated |  |
|  |  | 2011 - |  | 0.33 | ^2^ |
|  | Low-risk men | 1986 - 2001 |  | 0 | Personal communication |
|  |  | 2001 - 2006 |  | Linearly interpolated |  |
|  |  | 2006 |  | 0.098 | ^6,7^ |
|  |  | 2006 - 2008 |  | Linearly interpolated |  |
|  |  | 2008 |  | 0.1 | ^5^ |
|  |  | 2008 - 2011 |  | Linearly interpolated |  |
|  |  | 2011 - |  | 0.058 | ^2^ |
| Relative increase in testing at CD4 stage <200 | All | 1986-2035 | RR_test200_ | 1 - 5.4 | ^54^ |
| ART initiation rate (per year) | Professional FSW | 1986-2035 | ρ | 0.25 - 6 | ^55^ |
|  | Rest of population | 1986-2035 |  | 6 - 12 |  |
| ART dropout rate (per year) | Professional FSW | 1986-2035 | ϕ_PFSW_ | 0.023 - 0.11 | ^54,56^ |
|  | Client | 1986-2035 | ϕ_C_ | 0.023 - 0.11 |  |
|  | Low-risk women | 1986-2035 | ϕ_F_ |  |  |
|  | Low-risk men | 1986-2035 | ϕ_M_ |  |  |
| ART re-initiation rate (per year) | Professional FSW | 1986-2035 | ι_FSW_ | 0.25 - 1.5 | ^55^ |
|  | Low-risk | 1986-2035 | ι_rest_ | 0.25 - 1.5 |  |
| PrEP/TasP study testing rate (per year) | Professional FSW | 2015 - 2017 |  | 0.5 - 2 | Assumption |

#### Table S1E: PrEP parameters

| **Name** | **Group** | **Year** | **Symbol** | **Value** | **Source** |
| --- | --- | --- | --- | --- | --- |
| Testing rate when on PrEP | Professional FSW | 2015 - 2017 | τ_i_^11^ & τ_i_^00^ | 4 | ^57,58^ |
| PrEP offering rate (per year) | Professional FSW | 2015 - 2017 | ξ_i_ | 0.1 - 0.2 |  |
| PrEP acceptance rate (per year) | Professional FSW | 2015 - 2017 | σ_i_ | 86% |  |
| Fraction in perfect PrEP adherence category | Professional FSW | 2015 - 2017 | ζ_a_ | 36.9% |  |
| Fraction in intermediate PrEP adherence category | Professional FSW | 2015 - 2017 | ζ_b_ | 16.5% |  |
| Fraction in PrEP non-adherence category | Professional FSW | 2015 - 2017 | ζ_c_ | 46.6% |  |
| PrEP drop out rate (per year) | Professional FSW | 2015 - 2017 | κ | 0.45 - 0.64 |  |
| PrEP efficacy | Daily adherence | 2015 - 2017 | eP^1a^ | 0.9 - 0.95 | ^59^ |
|  | Intermediate adherence | 2015 - 2017 | eP^1b^ | 0.3 - 0.49 |  |
|  | Non-adherence | 2015 - 2017 | eP^1c^ | 0 | ^60,61^ |

#### Table S1F: TasP parameters

| **Name** | **Group** | **Year** | **Symbol** | **Value** | **Source** |
| --- | --- | --- | --- | --- | --- |
| TasP initiation rate (per year) | Professional FSW | 1986 - 2015 |  | 0 | No TasP intervention |
|  |  | 2015 - 2017 |  | 0.5 - 5 | ^58^ |
| Fraction of professional FSW virally suppressed | Professional FSW | 2015 - |  | 0.75 - 0.87 | ^37,58^ |

### Section 5: Cross validation data

**Table S2:** **Cross validation data**

| **Outcome** | **Year** | **Value** | **Source** |
| --- | --- | --- | --- |
| HIV prevalence professional FSW (%) | 1995 | 43.0 – 54.4 | ^3,4,9,11^ |
|  | 1998 | 36.6 – 44.7 |  |
|  | 2005 | 30.4 – 39.4 |  |
|  | 2012 | 23.0 – 32.2 |  |
| HIV prevalence clients (%) | 1998 | 5.9 – 11.6 | ^8,10,12^ |
|  | 2002 | 6.8 – 11.6 |  |
|  | 2008 | 3.5 – 9 |  |
|  | 2012 | 1.3 – 5.2 |  |
|  | 2015 | 0.6 – 3.5 |  |
| HIV prevalence all women (%) | 1998 | 2.4 – 4.8 | ^1,2,5^ |
|  | 2008 | 3.0 – 5.3 |  |
| HIV prevalence all men (%) | 1998 | 2.3 – 4.7 |  |
|  | 2008 | 1.2 – 3.0 |  |
| Deaths due to AIDS (all groups) | 1990 | 100 – 300 | ^30^ |
|  | 1991 | 100 – 300 |  |
|  | 1992 | 100 – 300 |  |
|  | 1993 | 100 – 300 |  |
|  | 1994 | 100 – 300 |  |
|  | 1995 | 100 – 400 |  |
|  | 1996 | 100 – 400 |  |
|  | 1997 | 100 – 500 |  |
|  | 1998 | 200 – 600 |  |
|  | 1999 | 200 – 900 |  |
|  | 2000 | 200 – 1200 |  |
|  | 2001 | 500 – 1500 |  |
|  | 2002 | 500 – 1500 |  |
|  | 2003 | 500 – 1500 |  |
|  | 2004 | 500 – 1500 |  |
|  | 2005 | 500 – 2000 |  |
|  | 2006 | 500 – 2000 |  |
|  | 2007 | 500 – 2000 |  |
|  | 2008 | 500 – 2000 |  |
|  | 2009 | 200 – 900 |  |
|  | 2010 | 200 – 900 |  |
|  | 2011 | 200 – 800 |  |
|  | 2012 | 200 – 1300 |  |
|  | 2013 | 200 – 1300 |  |
|  | 2014 | 200 – 1300 |  |
|  | 2015 | 200 – 1300 |  |
|  | 2016 | 100 – 700 |  |
| Proportion of HIV+ on ART and virally suppressed (all groups combined) | 2015 | 14 – 30 | ^30^ |
|  | 2016 | 15 – 32 |  |
|  | 2017 | 28 – 60 |  |

### Section 6: Model fits

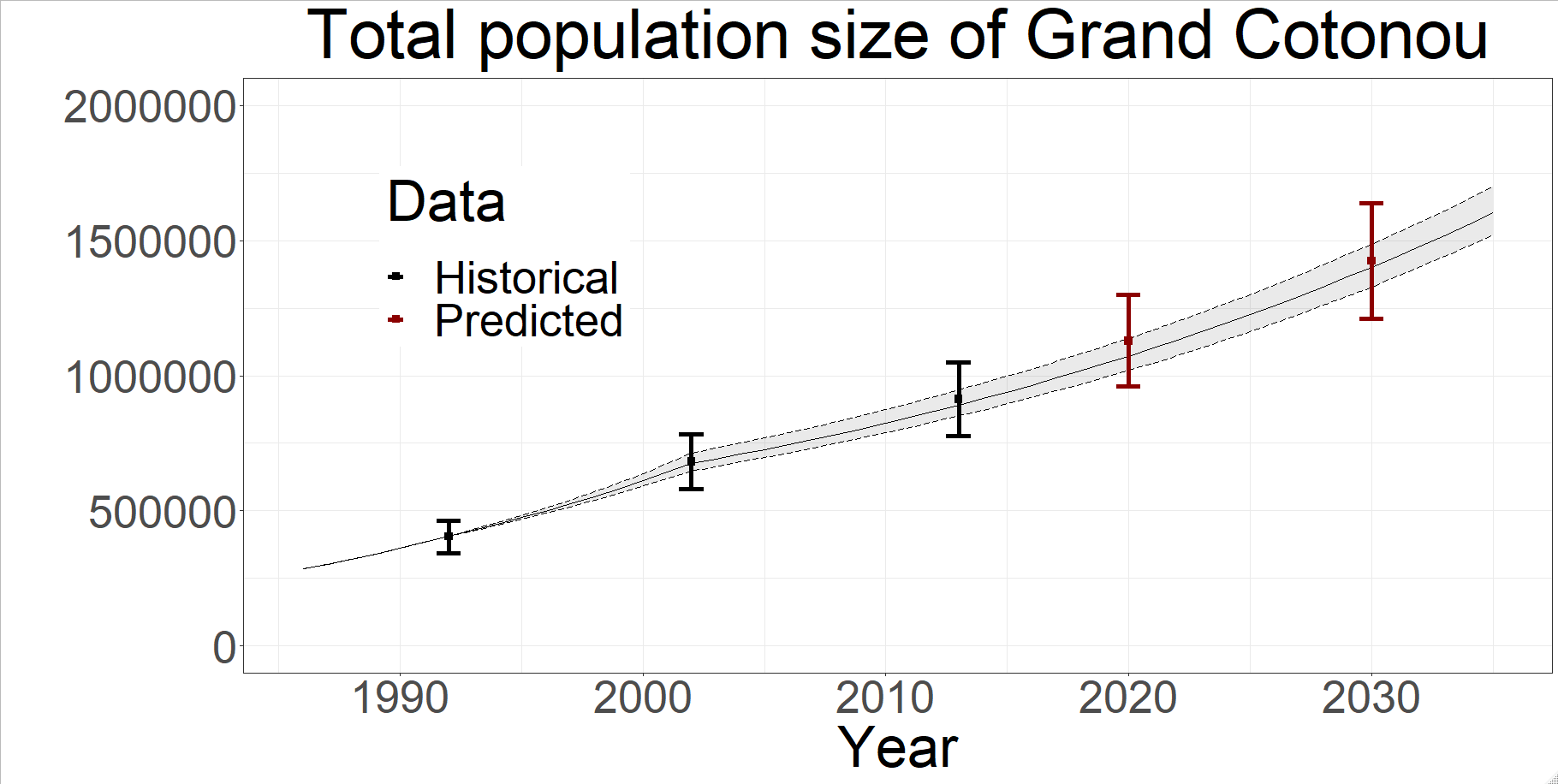

**Figure S3.** Posterior model predictions for total population size of Grand Cotonou from the 111 posterior parameter sets, showing median (solid line) and 95%UI across all fits (shaded regions) of counterfactual (no PrEP or TasP) scenario, compared to available fitting historical demographic data shown in thick bars (black) and estimated future prediction assuming constant population growth rate (red).

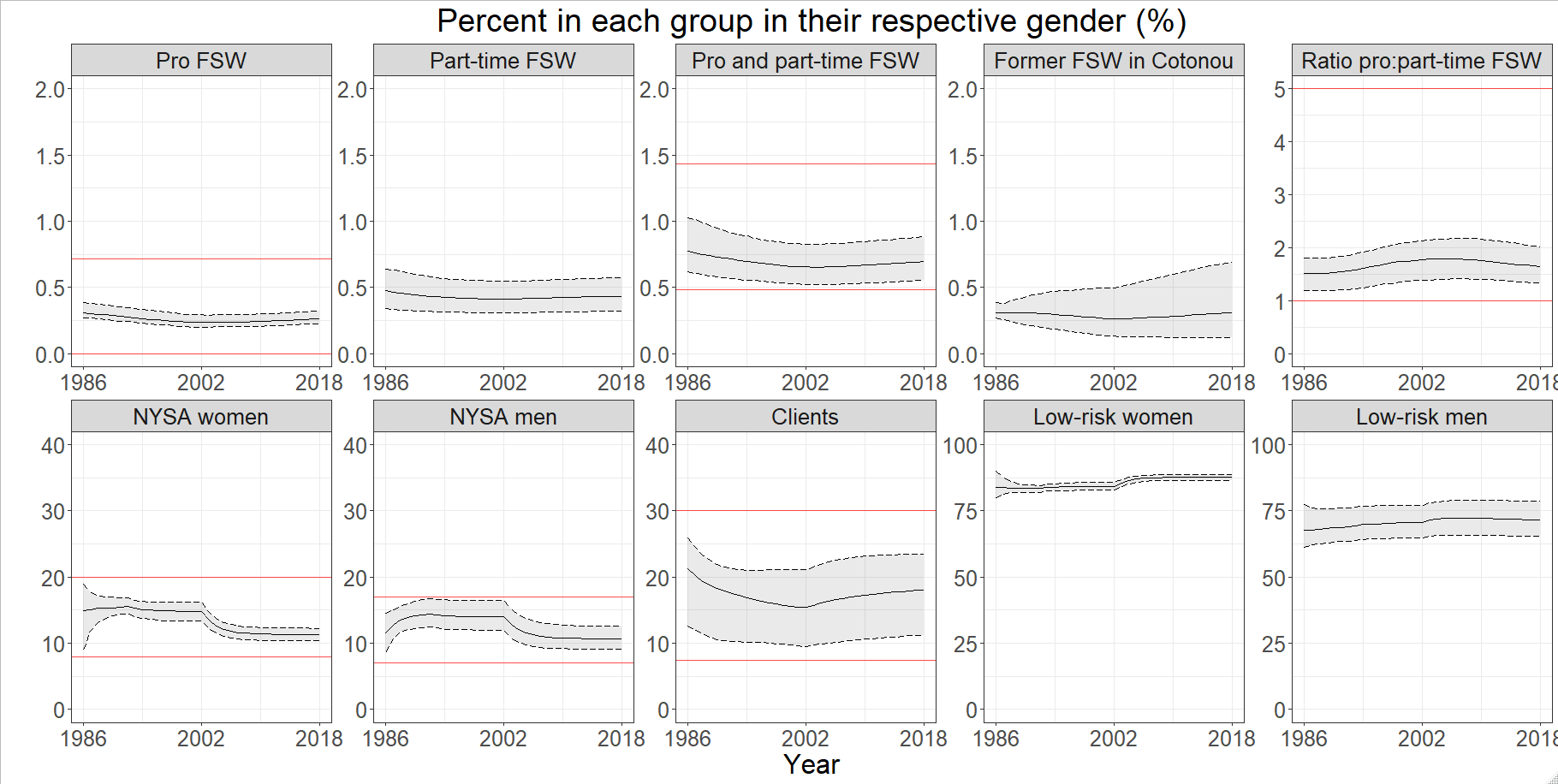

**A**

**B**

**C**

**D**

**E**

**F**

**G**

**H**

**I**

**J**

**Figure S4.** Posterior model predictions for the percent of each group in their respective gender (e.g. the percent of women who are professional FSW), from the 111 posterior parameter sets, showing median (solid line) and 95%UI across all fits (shaded regions) of counterfactual (no PrEP or TasP) scenario, compared to available fitting historical demographic data shown in red lines.

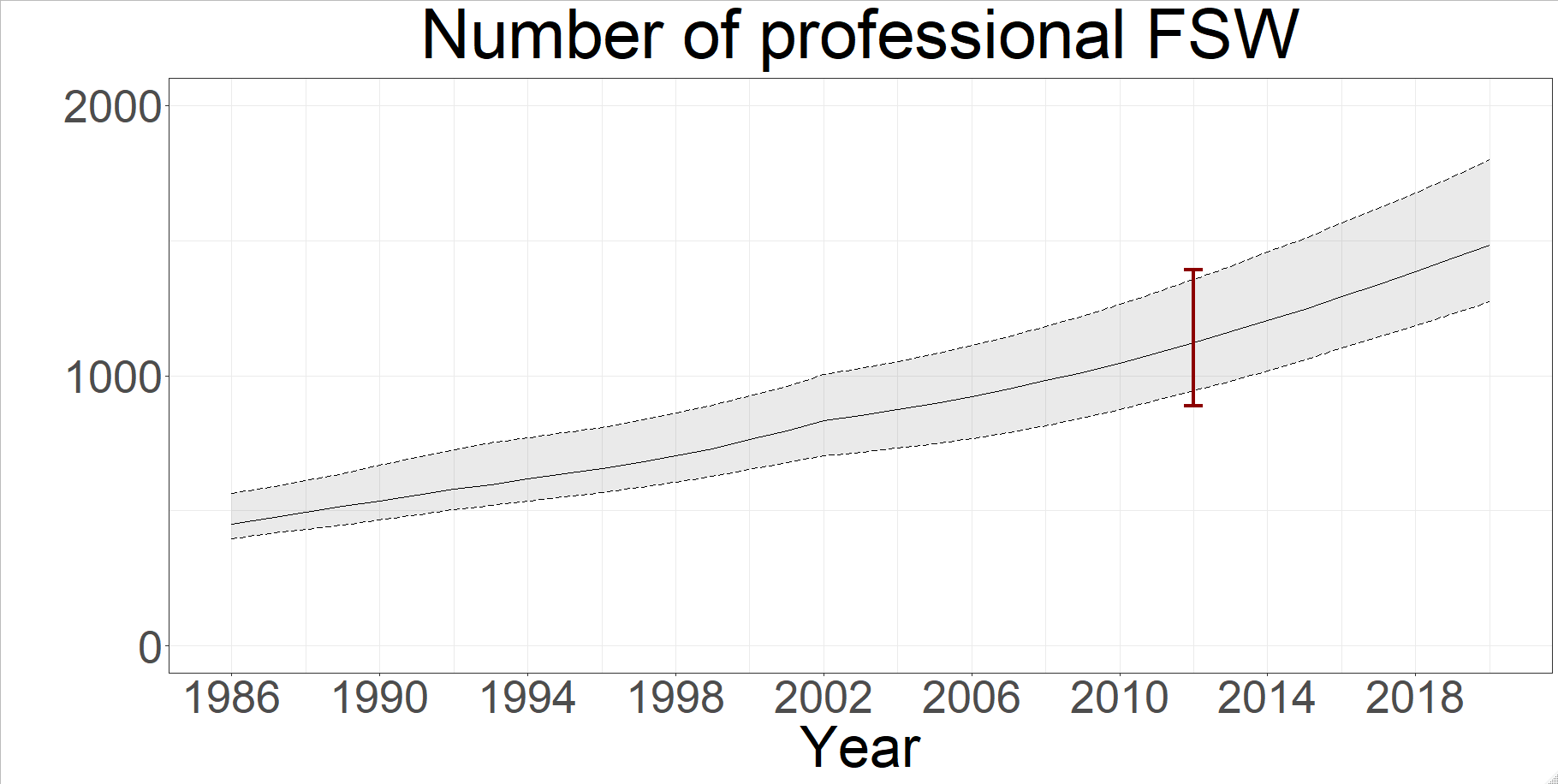

**Figure S5.** Posterior model predictions of number of professional FSW from the 111 posterior parameter sets, showing median (solid line) and 95% UI across all fits (shaded regions) of counterfactual (no PrEP or TasP) scenario, compared to available fitting historical demographic data shown with a red bar.

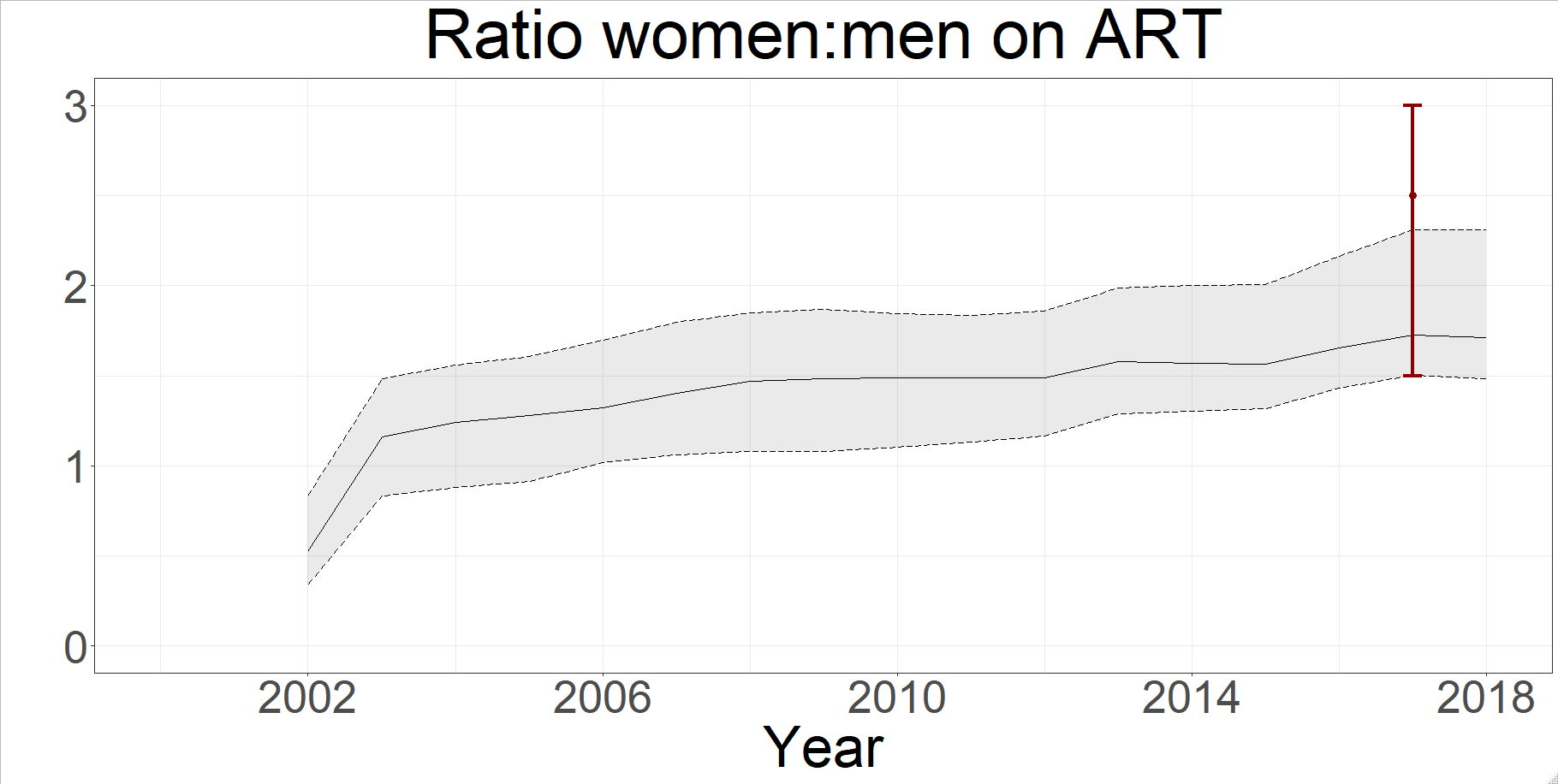

**Figure S6.** Posterior model predictions of the ratio of women to men on ART from the 111 posterior parameter sets, showing median (solid line) and 95% UI across all fits (shaded regions) of counterfactual (no PrEP or TasP) scenario, compared to available fitting intervention data shown with a red bar.

### Section 7: Model cross-validation

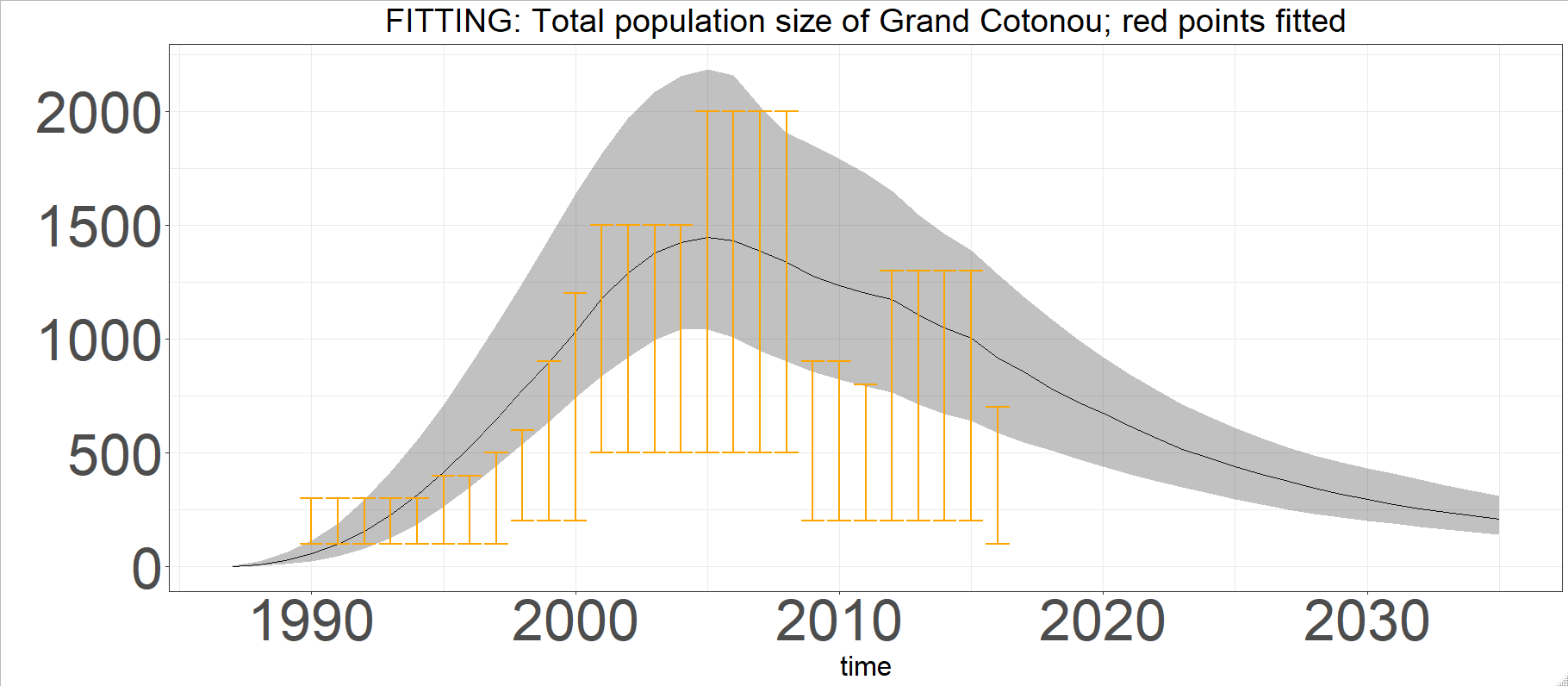

**Figure S7.** Posterior model predictions from the 111 posterior parameter sets, showing median (solid line) and 95% UI across all fits (shaded regions) of counterfactual (no PrEP or TasP) scenario, of annual deaths due to HIV infection compared to available cross-validation UNAIDS Reference Group Spectrum model estimates (orange bars) in Cotonou (equivalent to “Littoral”)^30^.

### Section 8: Posterior parameters

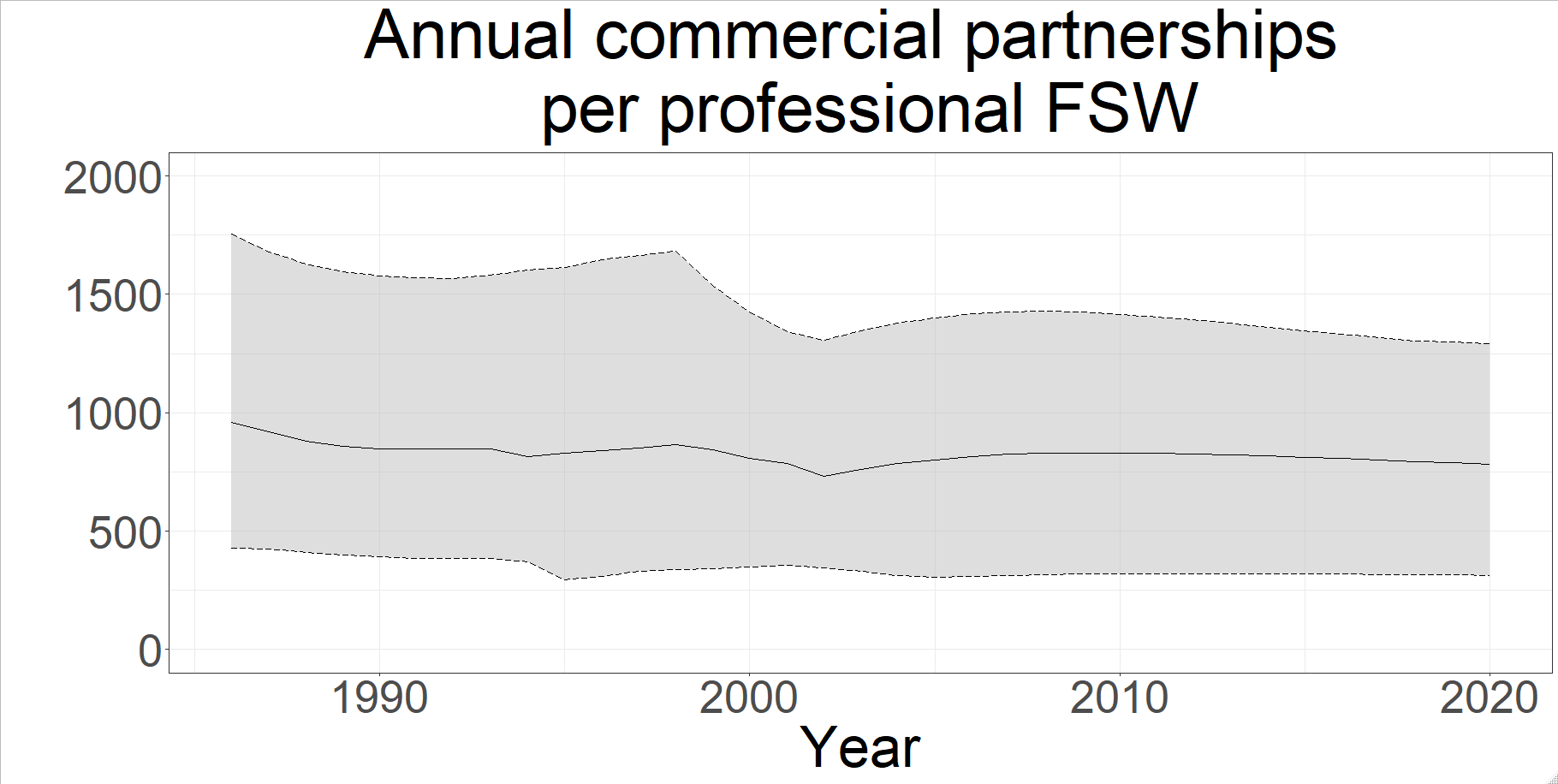

**Figure S8.** Posterior annual number of commercial partnerships between professional FSW and clients from the 111 posterior parameter sets, showing median (solid line) and 95% UI across all fits (shaded regions) of counterfactual (no PrEP or TasP) scenario.

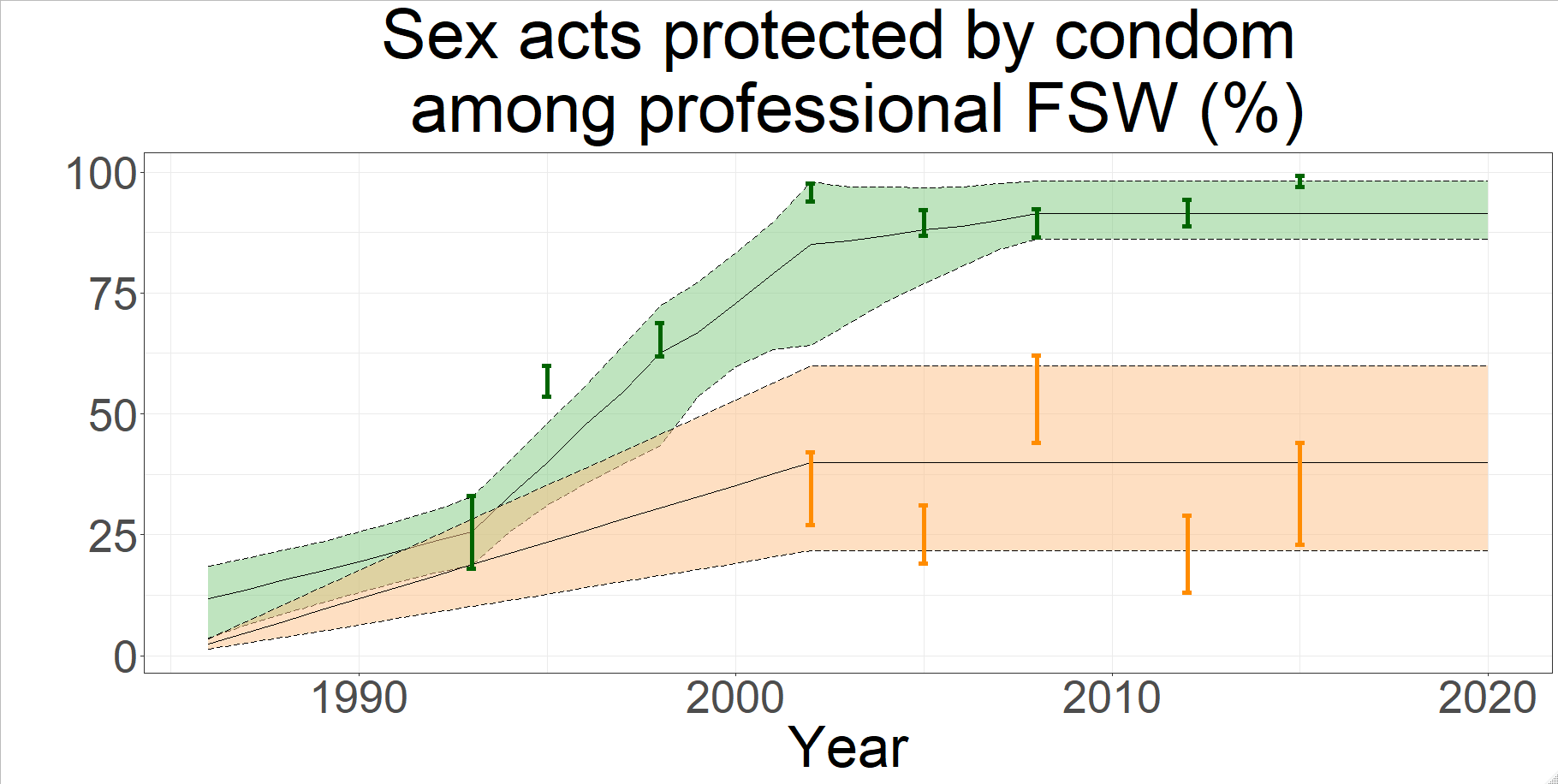

**Figure S9.** Posterior percent of commercial and non-commercial partnerships (shown in green and orange respectively) protected by condoms between professional FSW and clients from the 111 posterior parameter sets, showing median (solid line) and 95% UI across all fits (shaded regions) of counterfactual (no PrEP or TasP) scenario, compared to available historical behavioural data in bars.

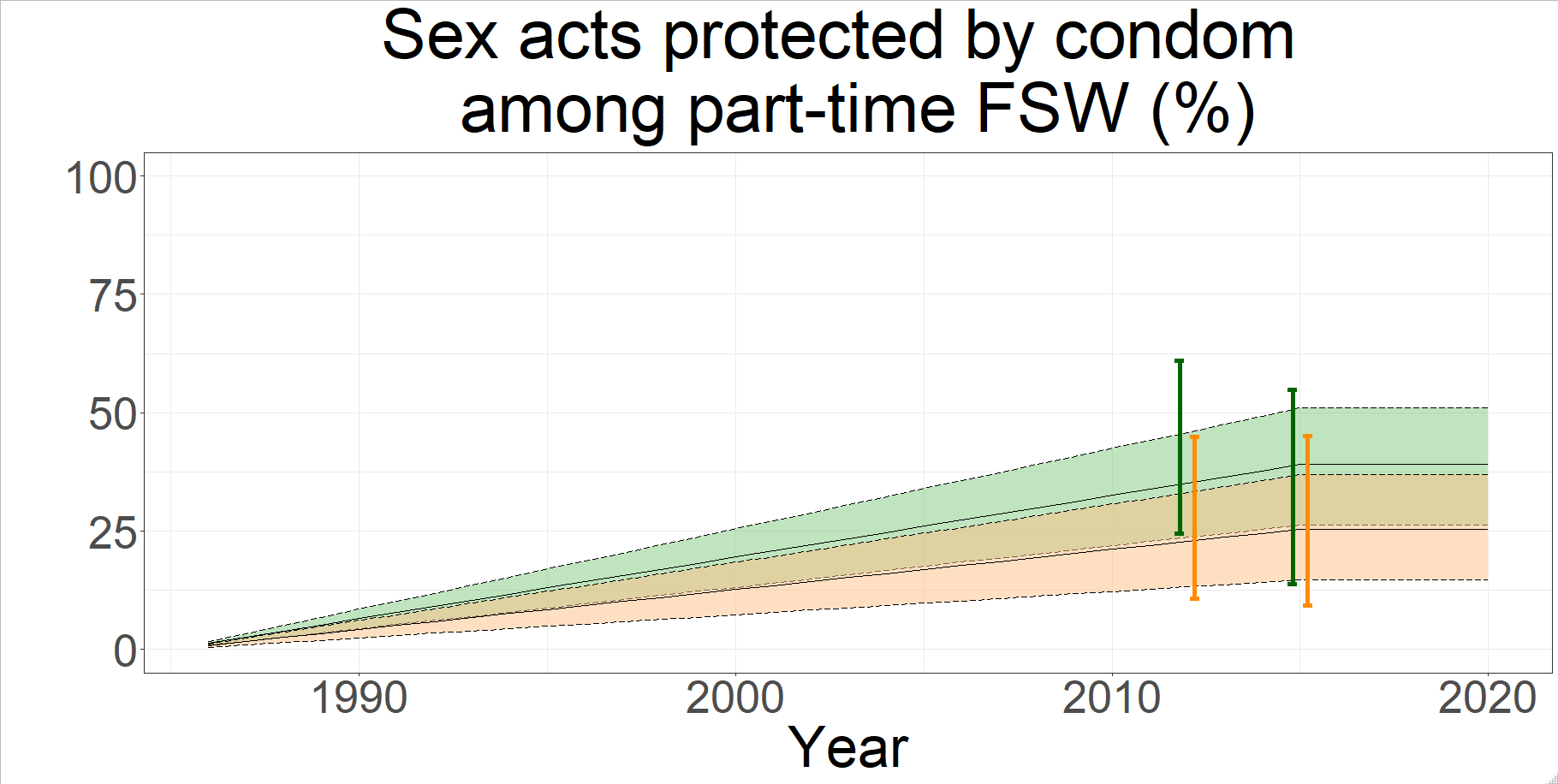

**Figure S10.** Posterior percent of commercial and non-commercial partnerships (shown in green and orange respectively) protected by condoms between part-time FSW and clients from the 111 posterior parameter sets, showing median (solid line) and 95% UI across all fits (shaded regions) of counterfactual (no PrEP or TasP) scenario, compared to available historical behavioural data in bars.

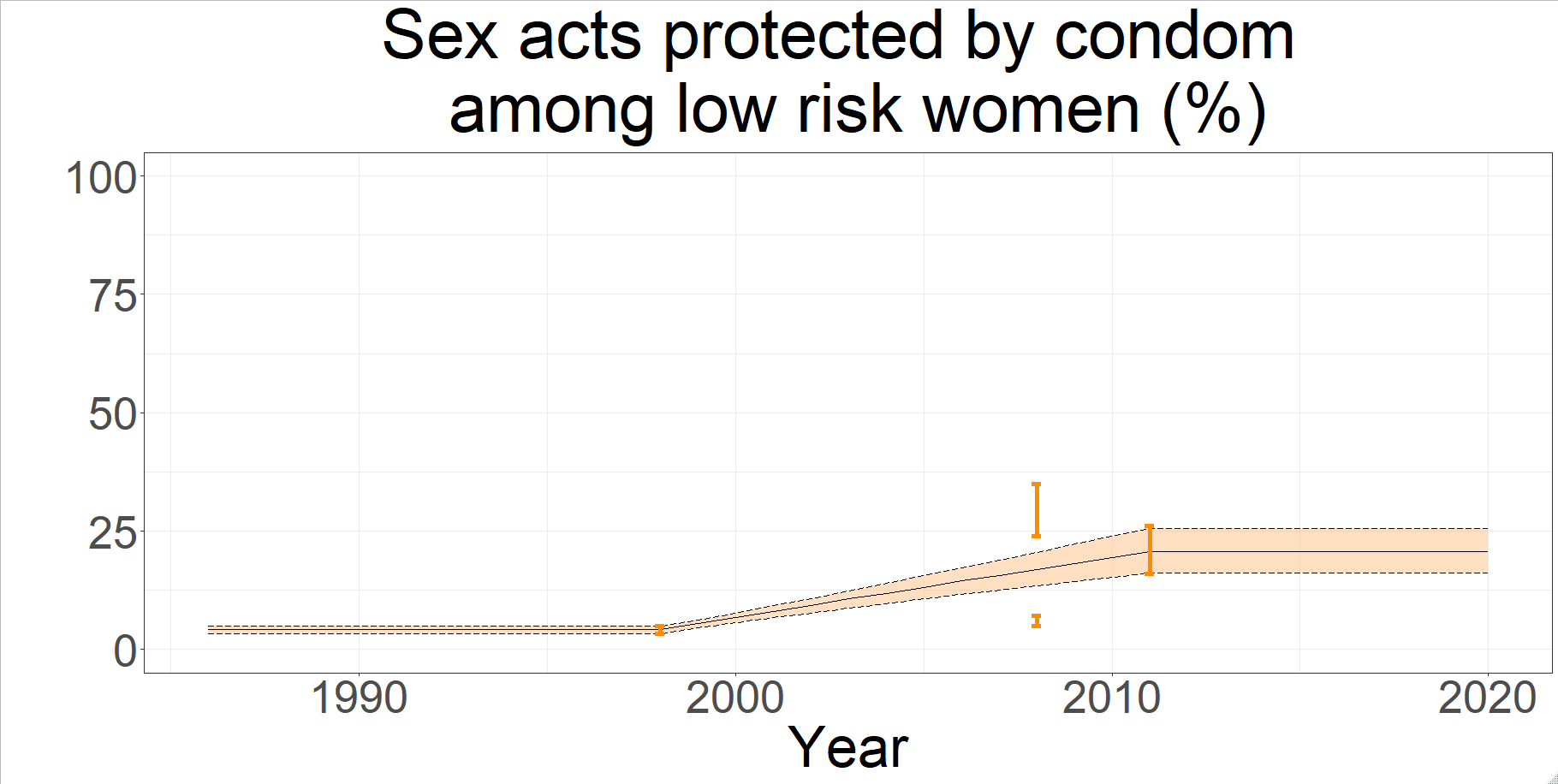

**Figure S11.** Posterior percent of non-commercial partnerships between low-risk women and their partners (clients and low-risk men) protected by condoms from the 111 posterior parameter sets, showing median (solid line) and 95% UI across all fits (shaded region) of counterfactual (no PrEP or TasP) scenario, compared to available historical behavioural data in orange bars.

### Section 9: Impact sensitivity Analyses

#### Increasing PrEP coverage

We explored an increased PrEP scale-up scenario to 69% among HIV- pFSW. This was achieved by allowing PrEP re-initiations as in Scenario B2, but increasing PrEP initiation rate five-fold. We also simulated this scenario with i) PrEP adherence as observed, and ii) perfect PrEP adherence.

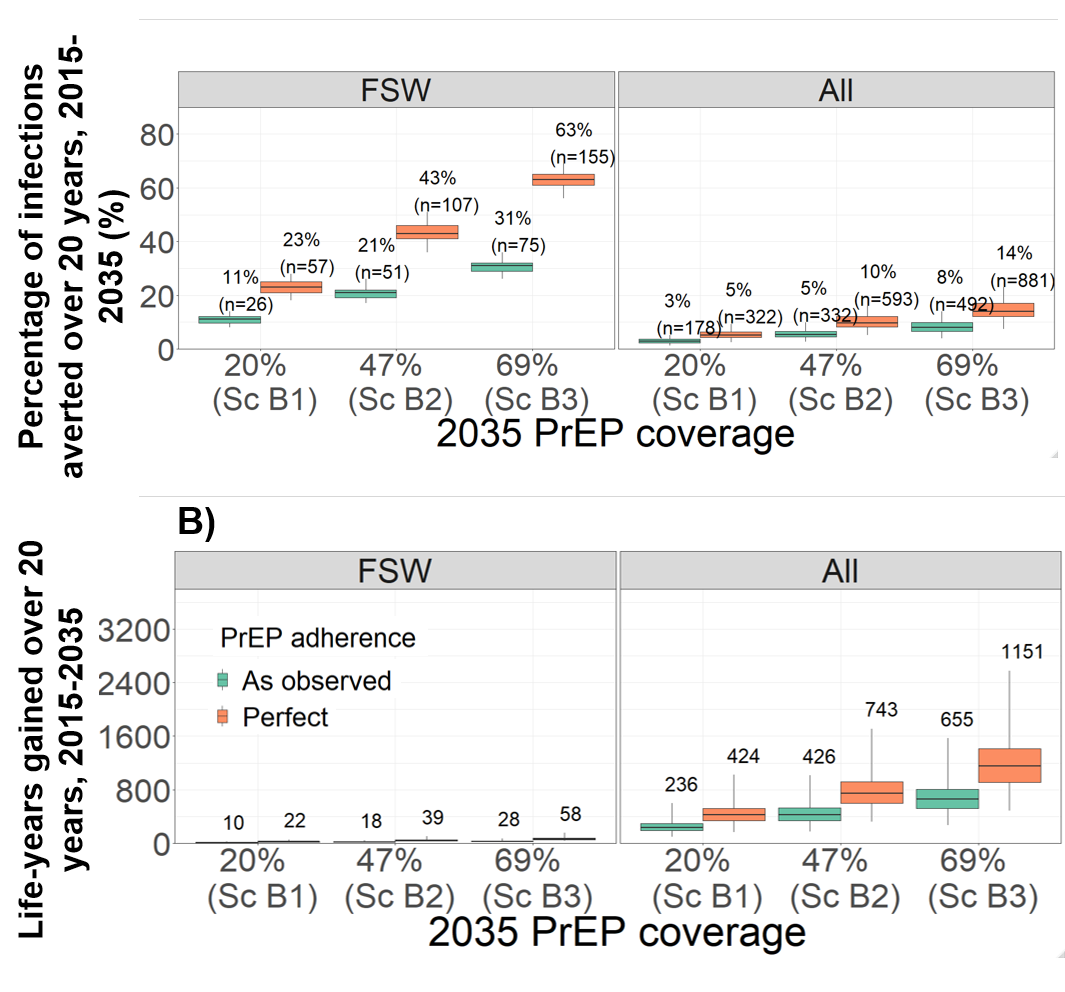

**Figure S12.** Impact of long-term 20-year PrEP intervention scenarios in terms of percentage of infections prevented (A) and life-years gained (B) among professional FSWs (“FSW”) and the whole population (“All”), compared to the counterfactual scenario (no PrEP). Boxplots represent the median impact (central horizontal line), 25^th^ to 75^th^ percentiles (box) and 95% UI (whiskers) across 111 parameter sets. The PrEP coverages represent median posterior values in 2035 (among HIV- pFSW) across all model fits for each scenario as shown in Table 3, as well as a scenario of 69% PrEP coverage. Adherence to PrEP assumed either as observed in the 2-year study (green), or perfect (orange). Labels in panels A shows median percentage (and number in brackets) of infections prevented; panel B shows median life-years gained.

#### Exploring different assumptions on PrEP efficacy

The Partners-in-PrEP efficacy trial (Donnell et al. 2014 ^59^) was conducted among serodiscordant African couples, which evaluated the efficacy of tenofovir disoproxil fumarate and TDF with emtricitabine (FTC/TDF), the same prophylactic component in the PrEP/TasP demonstration study.

They observed that most study participants displayed blood plasma tenofovir concentrations that were either below the assay limit of quantification (< 0.3 ng/ml), or consistent with daily adherence (>40 ng/ml). Few participants had a concentration between 0.3 – 40 ng/ml, from as little as 8% to at most 14%, depending on the visit. The study authors calculated PrEP efficacy at different thresholds of adherence: >0.3 ng/ml, >10 ng/ml and >40 ng/ml. Because so few participants had an intermediate adherence, those that had detectable tenofovir were almost always also daily adherers and as a result the estimated efficacy of PrEP for each threshold were very similar.

We assumed PrEP efficacy for daily adherers in our study was 90 – 95% (modelled as per sex act reduction in HIV transmission), consistent with the adjusted risk reduction for women on FTC/TDF at tenofovir concentration >40 ng/ml estimated by Donnell et al. We assumed that undetectable tenofovir conferred 0% PrEP efficacy. Based on expert opinion, we assumed that intermediate adherence (0.3 – 40 ng/ml) was associated with 30 – 49% efficacy.

Figure S14 demonstrates the results of the PrEP extension (Sc B1) and PrEP scale-up (Sc B2) scenarios, exploring different assumptions for PrEP efficacy for the intermediate (0.3 – 40 ng/ml) adherence category:

i) As assumed in the main scenarios (30 – 49%)
ii) 0% (pessimistic)
iii) 92% (optimistic)

Even though we explored the effect of varying efficacy of intermediate PrEP adherence from 0 to 92%, we found that the impact of PrEP interventions did not substantially vary. This is because intermediate PrEP adherence was observed in only 17% of visits, with the vast majority of blood plasma tenofovir concentration samples being consistent either with daily adherence or non-adherent (undetectable).

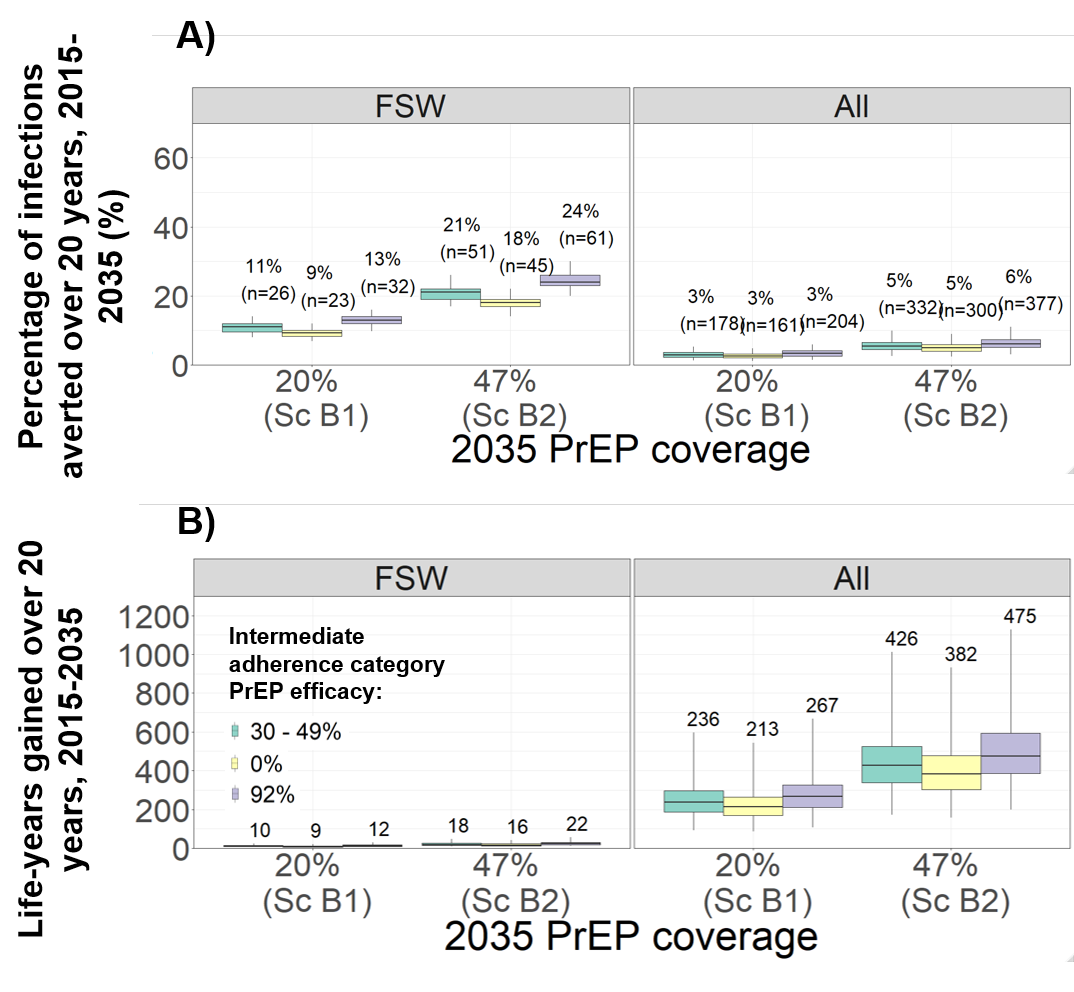

**Figure S13.** Impact of long-term 20-year PrEP intervention scenarios in terms of percentage of infections prevented (A) and life-years gained (B) among professional FSWs (“FSW”) and the whole population (“All”), compared to the counterfactual scenario (no PrEP). Boxplots represent the median impact (central horizontal line), 25^th^ to 75^th^ percentiles (box) and 95% UI (whiskers) across 111 parameter sets. The PrEP coverages represent median posterior values in 2035 (among HIV- pFSW) across all model fits for each scenario as shown in Table 3. The per sex-act reduction in transmission (efficacy) of PrEP in the intermediate adherence category (0.3 – 40 ng/ml) is assumed to be 30 – 49% (green), 0% (yellow) or 92% (purple). Labels in panels A shows median percentage (and number in brackets) of infections prevented; panel B shows median life-years gained.

#### Exploring ART eligibility

As described in the main text, the counterfactual scenario (Sc C0) assumed that only HIV+ pFSW who had a count CD4 T-cell count < 500 cells/ml blood were eligible for ART. This was the standard of care before the start of the PrEP/TasP study in 2015.^62^ The TasP arm of the PrEP/TasP study involved three components: increased HIV testing, increased ART initiation and increased eligibility to ART of all pFSW HIV+ regardless of CD4 count. The main objective of our analysis (indeed also an objective of the PrEP/TasP study) was to evaluate the full impact of the PrEP/TasP study compared to the standard of care at the time. In order to include the impact of all three components of TasP, the counterfactual scenario had to maintain the 2015 eligibility criteria for ART (CD4 < 500) from 2015 onwards.

However, following the PrEP/TasP study’s completion in 2017, the eligibility criteria changed such that all HIV+ pFSW were eligible for ART. Had we assumed this change in the counterfactual scenario, then the full impact of TasP, including increased ART eligibility, could not have been estimated. In this section we describe a second counterfactual scenario (Sc C1 in Table S3), whereby the ART eligibility is expanded to all HIV+ pFSW in 2015. We compare incident infections and life-years between this new counterfactual scenario with the long-term 20-years TasP scenarios (TasP extension and scale-up), to evaluate the added impact of increased HIV testing and initiation, but without the difference in ART eligibility.

**Table S3. Description of new counterfactual scenario.**

| **Scenario** | | **Increased HIV testing among FSW†** | **Eligibility criteria (from 2015)** | **TasP initiation & dropout rates^‡^** | **Median coverage (%)** | **2017** | | **2035** |
| --- | --- | --- | --- | --- | --- | --- | --- | --- |
| C0 | Main counterfactual  (no PrEP or TasP) | - | CD4 < 500 | - | PrEP | 0% | | 0% |
|  |  |  |  |  | ART | 49% | | 52% |
| C1 | Second counterfactual  (no PrEP or TasP) | - | Any CD4 | - | PrEP  ART | 0%  64% | 0%  67% | |
| B3 | TasP extension | As observed | Any CD4 | As observed | PrEP | As counterfactual C0 | | |
|  |  |  |  |  | ART | 83% | | 81% |
| B4 | TasP scale-up | As observed | Any CD4 | Initiation 2x higher & dropout 4x lower than observed | PrEP | As counterfactual C0 | | |
|  |  |  |  |  | ART | 89% | | 88% |

† Increased testing among pFSW applied over the duration of the intervention (PrEP/TasP study scenarios: 2015-2017; long-term scenarios: 2015-2035)

‡ PrEP and TasP initiation rates among HIV- and HIV+ pFSW (PrEP/TasP study scenarios as observed: 2015-2016; long-term scenarios: 2015-2035)

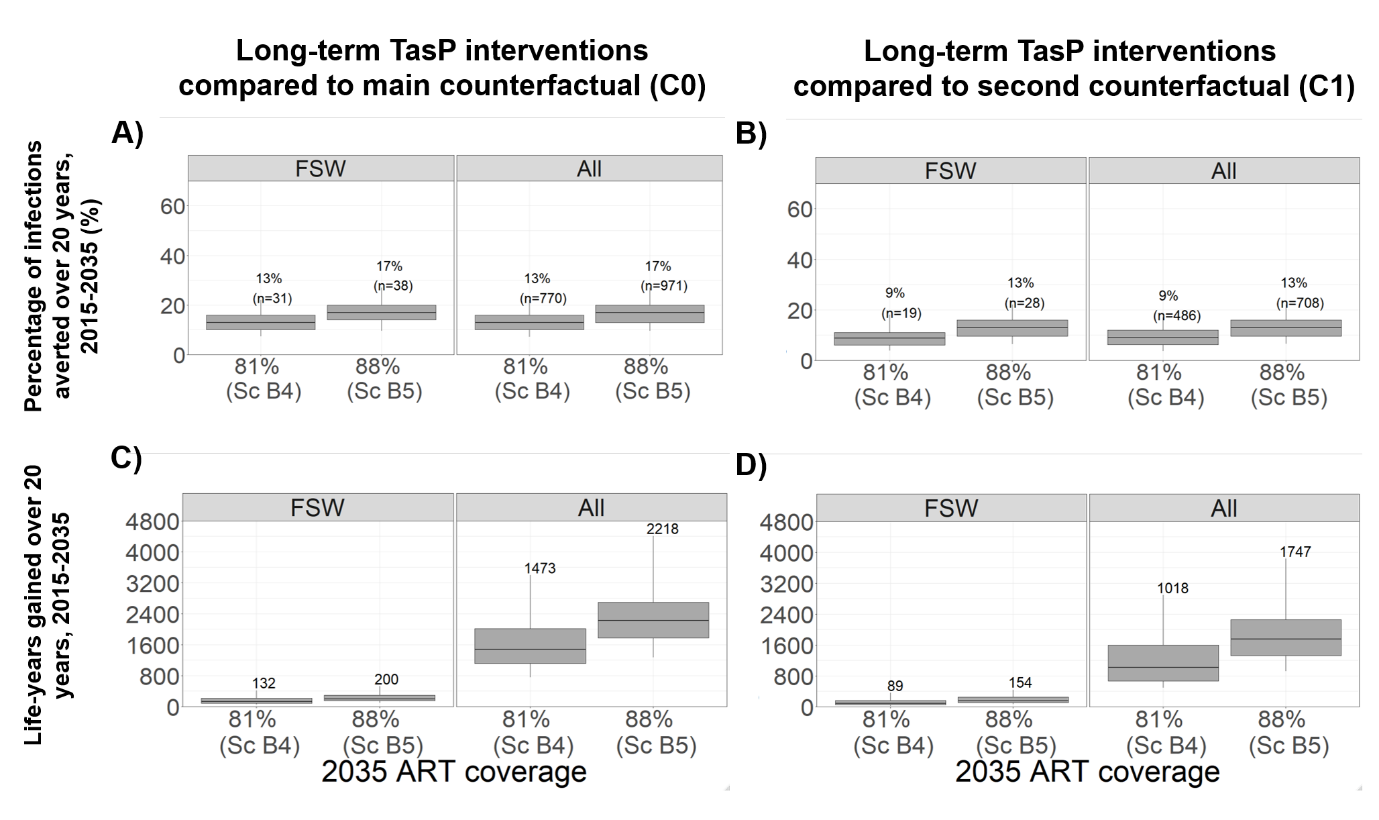

**Figure S14.** Impact of long-term 20-year TasP intervention scenarios in terms of percentage of infections prevented (A, B) and life-years gained (C, D) among professional FSWs (“FSW”) and the whole population (“All”), compared to counterfactual scenario C0 (A, C) and second counterfactual scenario C1 (B, D). Boxplots represent the median impact (central horizontal line), 25^th^ to 75^th^ percentiles (box) and 95% UI (whiskers) across 111 parameter sets. The ART coverages represent median posterior values in 2035 (among HIV- pFSW) across all model fits for each TasP scenario as shown in Table S3.

We estimate that expanding CD4 count to any HIV+ pFSW in 2015 (second counterfactual scenario C1) will lead to a 2035 ART coverage of 67% of HIV+ pFSW, compared to 52% in the main counterfactual scenario. The impact of TasP extension and scale-up (to median 2035 ART coverage of 81% and 88% respectively) is shown in Figure S15, panels B and D. TasP extension would prevent 9% (4-18) and 9% (4-17) infections in FSW and overall respectively, and a TasP scale-up would prevent 13% (6-22) and 13% (7-22). Compared to the impact of TasP in main analysis (against a counterfactual with no change in ART eligibility), the reduced impact is around 30%. This can be interpreted as the relative contribution (in HIV infections prevented) of increased ART eligibility to all HIV+ FSW in the TasP intervention, and 70% as attributable to increased HIV testing and ART initiation.

In terms of life-years gained, the relative contribution of increased ART eligibility is 20-25%.

### Section 10: Cost and resource data

^[1]^Table S4 A Costs and resources included in the intervention and in the government scenario.

| **Cost category** (*allocation rule*) | **Intervention** (Source) | **Ministry of Health Scenario** |
| --- | --- | --- |
| **Fixed costs** | | |
| **Start-up**  (*cost allocated* *proportion of activities related to intervention only as determined through interview and review of training materials*) | - PI and PM time spent designing intervention study protocols. - In house training of staff - Travel costs associated with start-up activities - Consultants salary   (Project Records) | - PI costs removed - Travel costs removed |
| **Capital costs**  (*cost allocated proportion of study visits at the clinic as determined by project records*) | - Buildings - Laboratory - Equipment   (Project records) | - As per intervention |
| **Recurrent costs**   - Utility costs (allocated according to proportion of study visits at the clinic) - Staff costs (time spent on intervention determined through interview, detailed timesheets and time in motion studies) | - Electricity, water and waste (project records) - Administrative staff salaries - Clinical staff costs spent on administrative tasks - Four community workers and receptionist   (project records) | - As per intervention - Project manager and PI replaced with full time project manager - All other staff retained except for one receptionist and local staff salaries used for similar roles or equivalent (Ministry of Health) |
| **Variable costs** | | |
| Micro-costing of visits *(all variable costs allocated according to resources used as documented in clinical records and detailed time sheets)* | - Medications - Hepatitis B vaccine - Condoms - Clinical staff time - Laboratory test   (project records) | - Medications - No hepatitis B vaccine included - Clinical staff time with local salaries - Reduced laboratory test schedule but same cost |

**Table S4B Key unit laboratory and medication costs (USD 2020)**

| Laboratory test* | Unit cost (2020 USD) |
| --- | --- |
| Rapid Duo Syphilis/HIV test | 6.02 |
| Confirmatory HIV test | 4.55 |
| Hepatitis B serology | 29.84 |
| Syphilis RPR | 3.54 |
| Screening NG/CT by TAN | 8.91 |
| Suite of test for TasP (Full blood count, Glucose, transaminase and creatinine) 3000 | 6 |
| Suite of tests for PreP (Full blood count, creatinine, transaminases) 6000 | 11 |
| CD4 count | 8.55 |
| Viral load | 54.32 |
| Medications per month |  |
| ART | 17.28 |
| Co-trimazole | 2.58 |
| PrEP | 17.06 |

*Laboratory costs include consumables and staff time to perform test

**Table S4C Visit schedule in intervention and government scenario**

| **Visit** | **Intervention** | **Ministry of Health scenario** |
| --- | --- | --- |
| At each visit | Medical history and exam, Pre-test HIV and adherence counselling,  Phlebotomist,  One month medication dispensation | Medical history and exam, Pre-test HIV and adherence counselling,  Phlebotomist,  Three month medication dispensation |
| Screening visit | All women:   - Hepatitis B - HIV - Syphilis - FBC - Creatinine - Glucose - Transaminases - Pregnancy test - CD4 as required - Viral load as required | - Rapid HIV test - Syphilis RPR   TasP women:   - FBC - Creatinine - Glucose - Transaminases - Pregnancy test   PrEP women:   - Creatinine - Pregnancy test |
| Recruitment visit | TasP women:   - Chest X-ray - Pregnancy test - Gram stain   PrEP women:   - Rapid HIV test - Pregnancy test - Gram stain | TasP women:   - Chest X-ray   PrEP women:   - Rapid HIV test |
| Day 14 visit | No laboratory test | No day 14 visit |
| Month 3 | TasP women:   - Pregnancy test - CD4 - Gram stain   PrEP women:   - Rapid DUO test - RPR as required - Pregnancy test - Gram stain | TasP women: No visit  PrEP women:   - Rapid HIV test |
| Month 6 | TasP women:   - RPR as required - Pregnancy test - CD4 - Gram stain - FBC - Creatinine - Glucose - Transaminases - Viral load   PrEP women:   - Rapid DUO test - RPR as required - Pregnancy test - Gram stain - FBC - Creatinine - Glucose - Transaminases - PreP levels | TasP women:   - Pregnancy test - CD4 - FBC   PrEP women:   - Rapid HIV test - Creatinine |
| Month 9 | TasP women:   - Pregnancy test - CD4 - Gramstain   PrEP women:   - Rapid DUO test - RPR as required - Pregnancy test - Gram stain | TasP women: No visit  PrEP women:   - Rapid HIV test |
| Month 12 | TasP women:   - RPR as required - Pregnancy test - CD4 - Gram stain - FBC - Creatinine - Glucose - Transaminases - Viral load   PrEP women:   - Rapid DUO test - RPR as required - Pregnancy test - Gram stain - FBC - Creatinine - Glucose - Transaminases - PrEP levels | TasP women:   - Pregnancy test - CD4 - FBC - Viral load   PrEP women:   - Rapid HIV test - Creatinine |

**Table S4D. Sensitivity analysis**

| **Variable (Base case value)** | **Best case scenario**  **(Minimum)** | **Worst case scenario (Maximum)** |
| --- | --- | --- |
| Discount rate (4.5%) | 10% | 1% |
| Useful life of:   - Building (30 years) - Laboratory equipment (20 years) - IT equipment (5 years) - Start-up activities (4 years) | - 35 years - 25 years - 8 years - 5 years | - 25 years - 15 years - 2 years - 3 years |
| Allocation rule for capital and recurrent utility costs (proportion of intervention visits out of all visits conducted at the clinic) | -20% | +20% |
| Administrative staff time allocation to intervention (proportion of time spent on intervention) | -20% | +20% |
| Proportion of clinical time spent on intervention-only as determined by time sheets | Minimum proportion | Maximum proportion |
| Unit lab costs (median value of estimated resources used for each laboratory tests) | Minimum value of estimated resources used | Maximum value of estimated resources used |
| Medication costs (as per project records) | -20% | +20% |

### Section 11: Impact results

**Figure S14. Impact of Tasp/PreP implementation for 2 years of study period compared to standard ART care for FSW.**

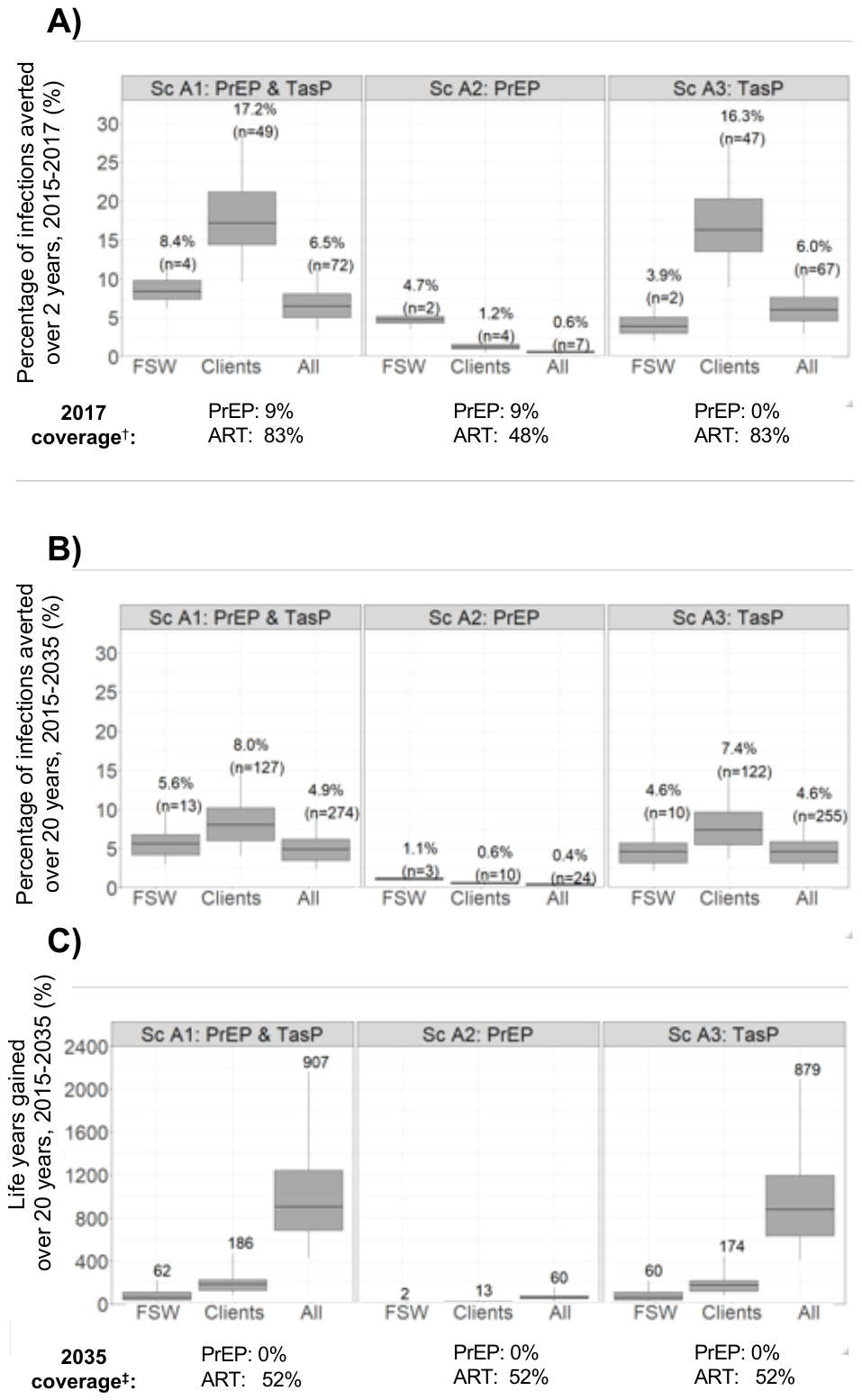

**Figure S15. Impact of implementing TasP and PreP for 20 years compared to standard ART care for FSW**

**Figure S16. Impact implementing both TasP and PrEP combined for 20 years when compared to standard ART for FSW.**

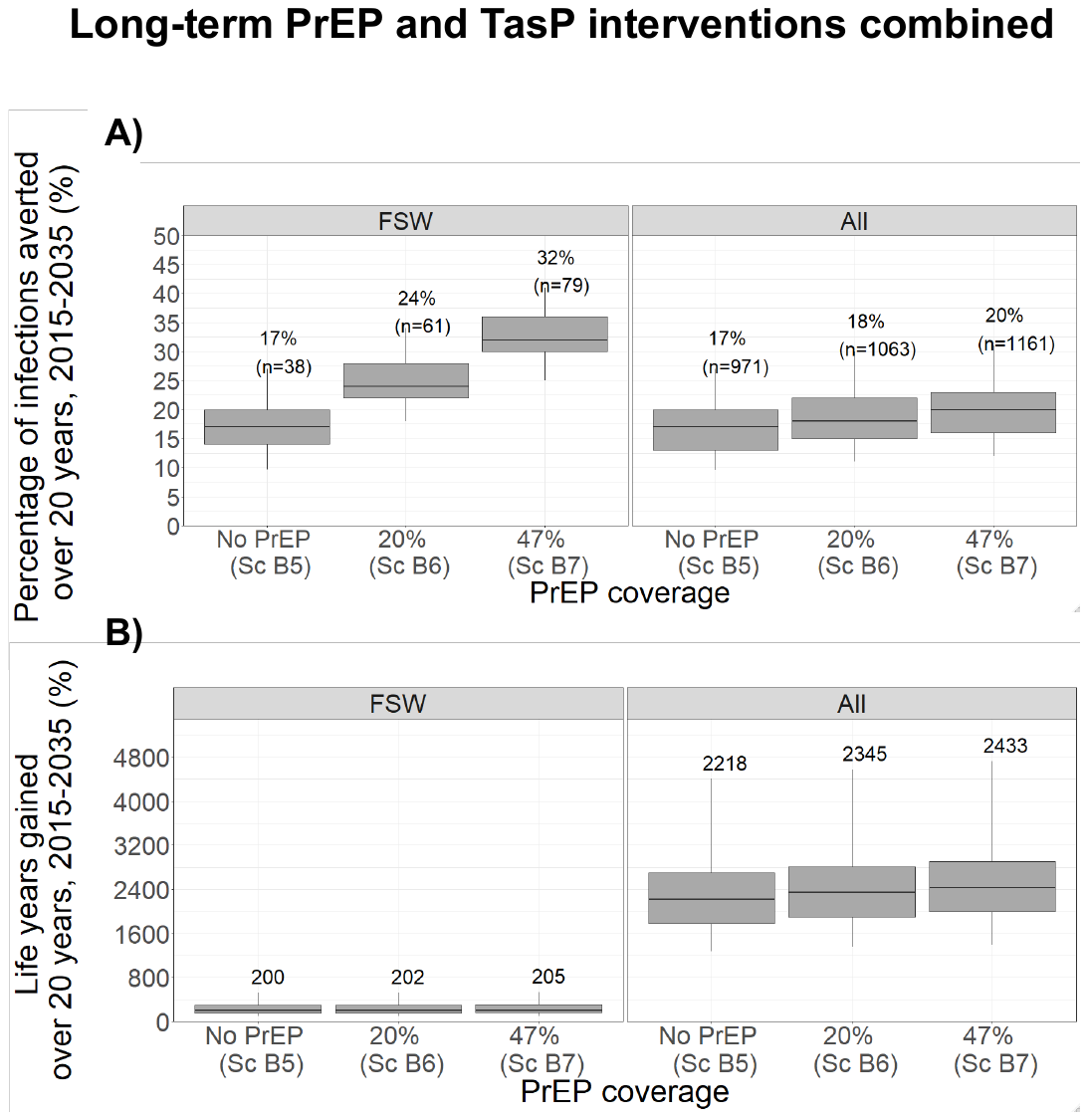

### Section 12: Cost-effectiveness results

**Figure S17** **Cost and cost-effectiveness of delivering PrEP at different coverage levels among HIV-negative FSW by Ministry of Health over 20 years compared to 80% coverage of TasP among HIV + positive FSW (USD2020)**

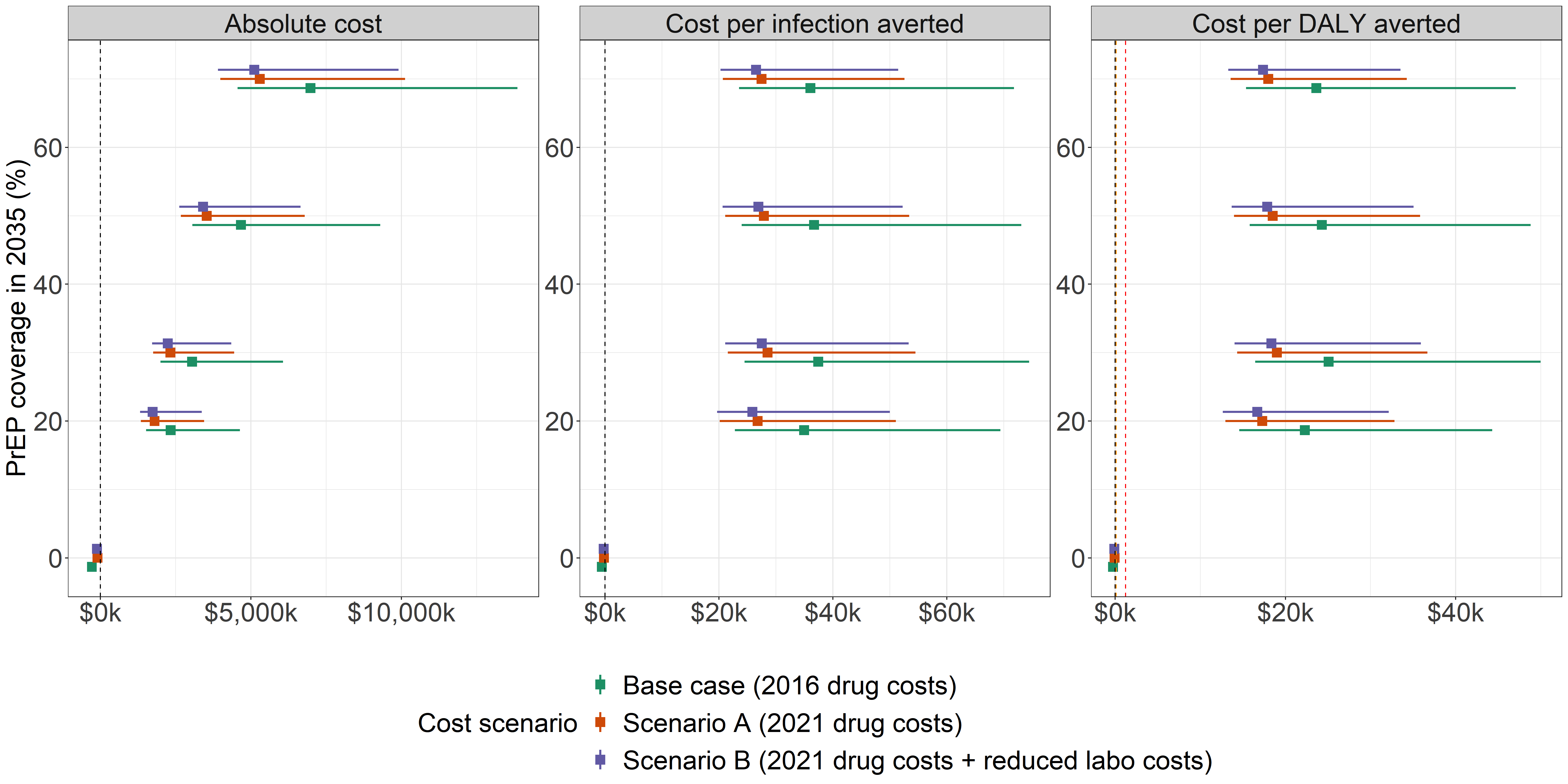

**Figure S18 Cost and cost-effectiveness of delivering PrEP at different coverage levels among HIV-negative FSW by Ministry of Health over 20 years compared to no PREP and standard ART care for HIV positive FSW (US2020).**

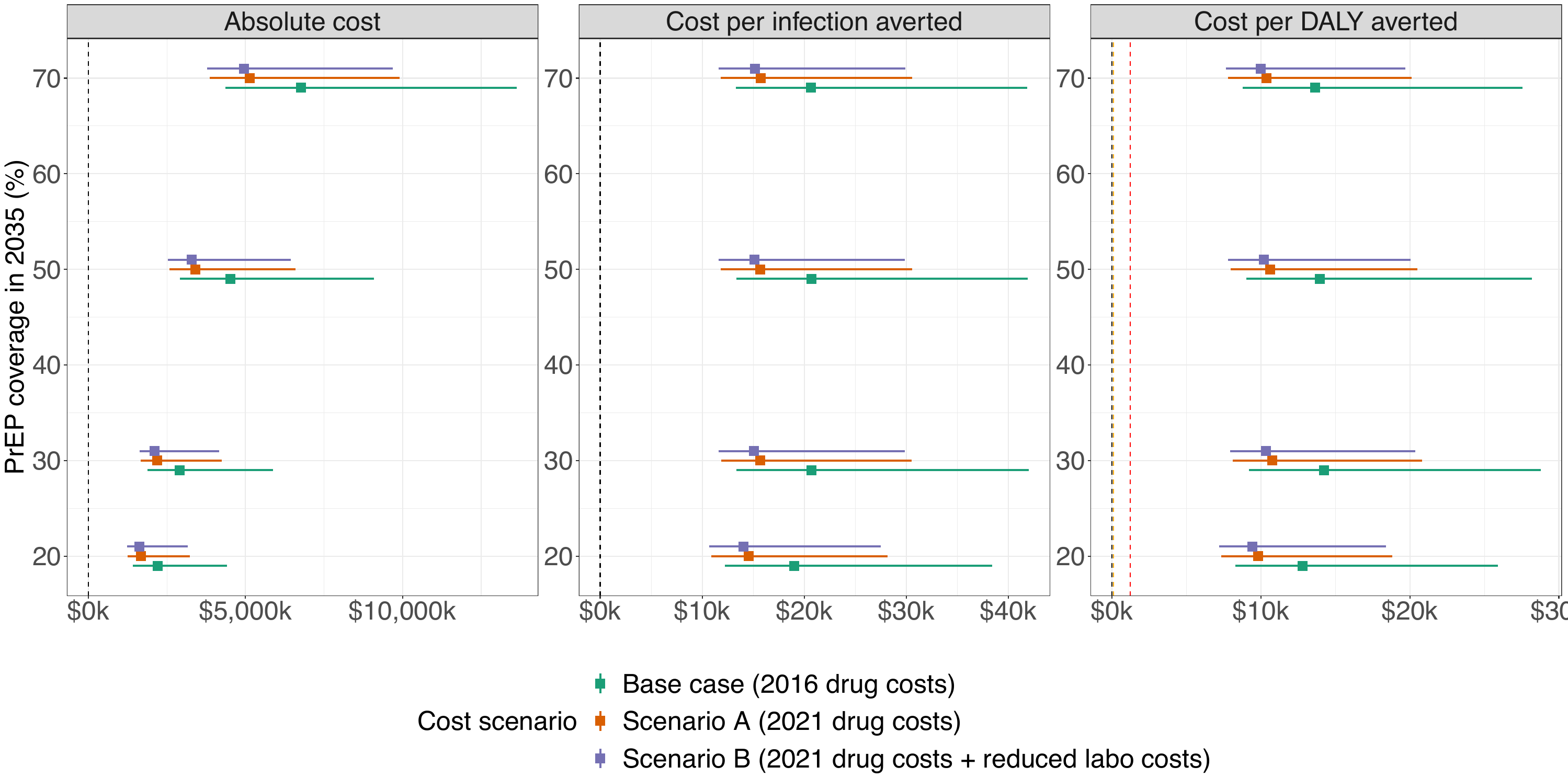

**Figure S19 Cost and cost-effectiveness of of PrEP at different adherence levels (4B) as delivered by Ministry of Health over 20 years compared to no PrEP and standard ART care for HIV+ positive FSW (USD 2020).**

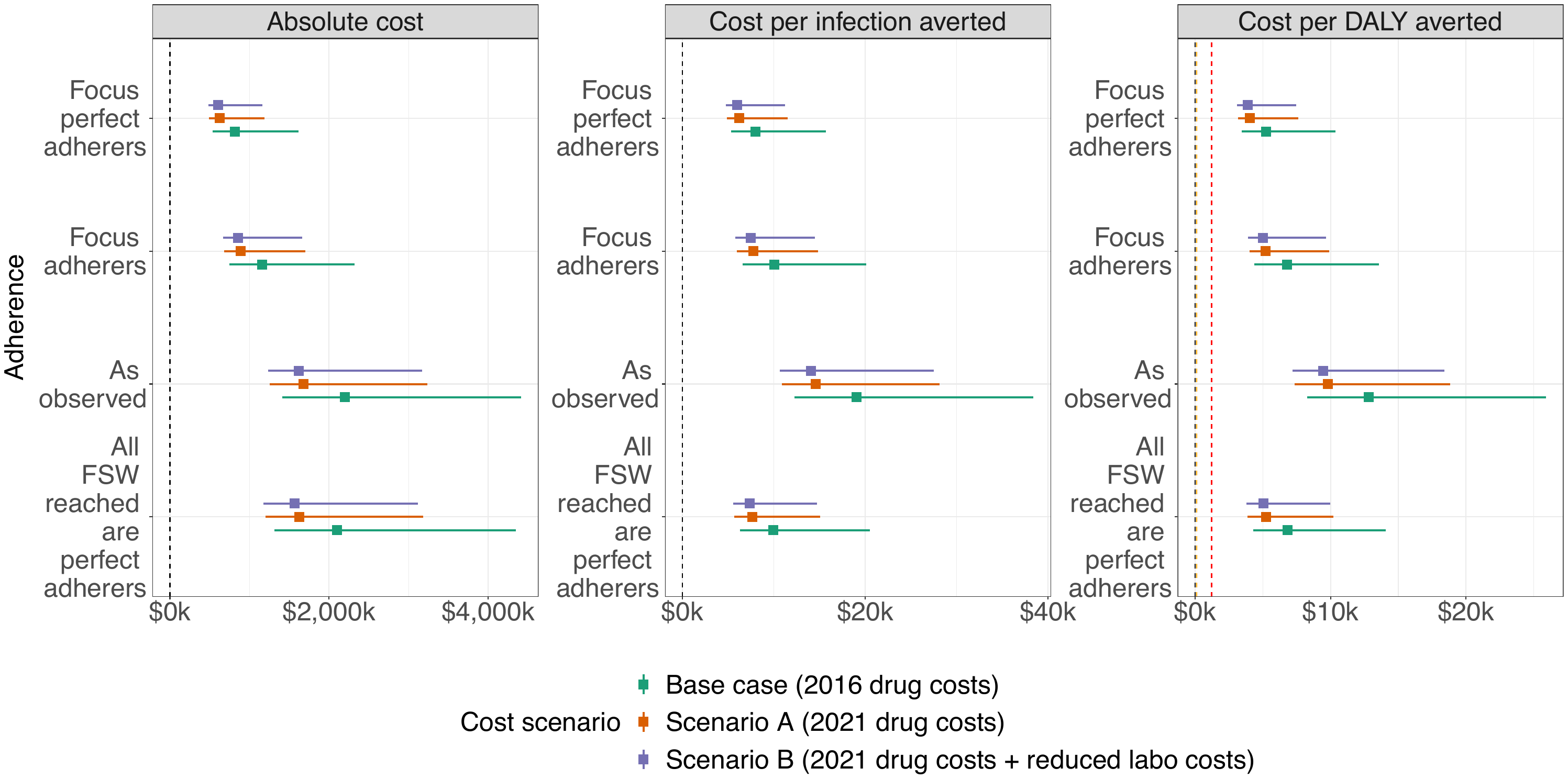

### References

**1.**	Behanzin L, Diabate S, Minani I, et al. Decline in HIV prevalence among young men in the general population of Cotonou, Benin, 1998-2008. *PLoS One.* 2012;7(8):e43818. doi: 10.1371/journal.pone.0043818.

**2.**	Institut National de la Statistique et de l’Analyse Économique (INSAE). Institut National de la Statistique et de l’Analyse Économique - INSAE/Bénin and ICF International. 2013. Enquête Démographique et de Santé du Bénin 2011-2012. Calverton, Maryland, USA: INSAE and ICF International. 2011.

**3.**	Alary M, Mukenge-Tshibaka L, Bernier F, et al. Decline in the prevalence of HIV and sexually transmitted diseases among female sex workers in Cotonou, Benin, 1993-1999. *AIDS.* 2002;16(3):463-470. <https://www.ncbi.nlm.nih.gov/pubmed/11834959>.

**4.**	Behanzin L, Diabate S, Minani I, et al. Decline in the prevalence of HIV and sexually transmitted infections among female sex workers in Benin over 15 years of targeted interventions. *J Acquir Immune Defic Syndr.* 2013;63(1):126-134. doi: 10.1097/QAI.0b013e318286b9d4.

**5.**	Behanzin L, Diabate S, Minani I, et al. Assessment of HIV-related risky behaviour: a comparative study of face-to-face interviews and polling booth surveys in the general population of Cotonou, Benin. *Sexually Transmitted Infections.* 2013;89(7):595-601. doi: 10.1136/sextrans-2012-050884.

**6.**	Institut National de la Statistique et de l’Analyse Économique (INSAE). Institut National de la Statistique et de l’Analyse Économique - INSAE/Bénin and ORC Macro. 2002. Enquête Démographique et de Santé au Bénin 2001. Calverton, Maryland, USA: INSAE/Bénin and ORC Macro. Available at <http://dhsprogram.com/pubs/pdf/FR133/FR133.pdf>. 2001.

**7.**	Institut National de la Statistique et de l’Analyse Économique (INSAE). Institut National de la Statistique et de l’Analyse Économique - INSAE/Bénin and ICF International. Enquête Démographique et de Santé du Bénin 2006. Calverton, Maryland, USA: INSAE and ICF International. 2006.

**8.**	Lowndes CM, Alary M, Gnintoungbe CA, et al. Management of sexually transmitted diseases and HIV prevention in men at high risk: targeting clients and non-paying sexual partners of female sex workers in Benin. *AIDS.* 2000;14(16):2523-2534.

**9.**	Ministry of Health Benin. *Enquête de surveillance de deuxième génération des IST/VIH/SIDA au Bénin (ESDG-2012): Travailleuses du sexe et Serveuses de bars/restaurants.* 2012.

**10.**	Ministry of Health Benin. *Enquête de surveillance de deuxième génération des IST/VIH/SIDA au Bénin (ESDG-2012): Camionneurs et Clients des Travailleuses du sexe.* 2012.

**11.**	Ministry of Health Benin. *Enquête de surveillance de deuxième génération relative aux IST, VIH et SIDA au Bénin (ESDG‐2015): Professionnelles de Sexe & Serveuses de Bar et Restaurants.* 2015.

**12.**	Ministry of Health Benin. Enquête de surveillance de deuxième génération relative aux IST, VIH et SIDA au Bénin (ESDG‐2015): Clients des TS, Camionneurs et Personnes Privées de Liberté. 2015.

**13.**	UNAIDS; Ministry of Health Benin; CNLS. *Rapport de suivi de la déclaration de politique sur le VIH/SIDA au Bénin 2016 (Country progress report, Benin).* 2016.

**14.**	UNAIDS; PNLS. *Rapport de suivi de la déclaration de politique sue le VIH/SIDA au Bénin 2010 (Country progress report, Benin).* 2010.

**15.**	Hindmarsh AC, Petzold LR. Algorithms and software for ordinary differential equations and differential- algebraic equations, Part I: Euler methods and error estimation. *Computers in Physics.* 1995;9(1). doi: 10.1063/1.168536.

**16.**	Fitzjohn R. "odin": ODE Generation and Integration. 2019; github.com/mrc-ide/odin.

**17.**	*"deSolve": Solvers for Initial Value Problems of Differential Equations ('ODE', 'DAE', 'DDE')* [computer program]. R-project, CRAN; 2019.

**18.**	Boily M-C, Baggaley RF, Wang L, et al. Heterosexual risk of HIV-1 infection per sexual act: systematic review and meta-analysis of observational studies. *The Lancet Infectious Diseases.* 2009;9(2):118-129. doi: 10.1016/s1473-3099(09)70021-0.

**19.**	Bigot A, I. Zohoun, M. De Bruyere. *Premiers cas de seropositivite anti-HIV-1 au Benin.* Vol 161987.

**20.**	Institut National de la statistique et d’analyse économique du Bénin (INSAE). *Recensement General de la Population et de l'Habitation 1er Edition.* 1979.

**21.**	Institut National de la statistique et d’analyse économique du Bénin (INSAE). *Recensement General de la Population et de l'Habitation 2eme Edition.* 1992.

**22.**	Institut National de la statistique et d’analyse économique du Bénin (INSAE). *Quatrième Recensement Général de la Population et de l’Habitation* 2013.

**23.**	Institut National de la statistique et d’analyse économique du Bénin (INSAE). *Recensement General de la Population et de l'Habitation 3eme Edition.* 2002.

**24.**	Vandepitte J. Estimates of the number of female sex workers in different regions of the world. *Sexually Transmitted Infections.* 2006;82(suppl_3):iii18-iii25. doi: 10.1136/sti.2006.020081.

**25.**	Projet Équité en Santé Sexuelle et Santé. *Cartographie des Travailleuses de Sexe des communes de Cotonou, Abomey-Calavi, Sèmè-Podji et Parakou: Rapport Synthèse du Mapping, Benin, 2013-2014.*

**26.**	Lowndes CM, Alary M, Meda H, et al. Role of core and bridging groups in the transmission dynamics of HIV and STIs in Cotonou, Benin, West Africa. *Sexually Transmitted Infections.* 2002;78(Supplement 1):i69-i77. doi: 10.1136/sti.78.suppl_1.i69.

**27.**	The Global HIV/AIDS Program World Bank. *West Africa HIV/AIDS Epidemiology and Response Synthesis: Implications for prevention.* 2008.

**28.**	Boyle Torrey B MM, Way P. *Blood donors and AIDS in Africa: The gift relationship revisited.* US Bureau of the Census1990.

**29.**	UNDP. World Population Prospects: The 2015 revision—key findings and advance tables. New York, NY: Population Division of the Department of Economic and Social Affairs of the United Nations Secretariat, 2015 Contract No.: ESA/P/WP.241. 2019.

**30.**	The UNAIDS Reference Group on Estimates, Modelling and Projections;. Spectrum model. 2019; <https://aidsinfo.unaids.org/>.

**31.**	Looker KJ, Elmes JAR, Gottlieb SL, et al. Effect of HSV-2 infection on subsequent HIV acquisition: an updated systematic review and meta-analysis. *Lancet Infect Dis.* 2017;17(12):1303-1316. doi: 10.1016/S1473-3099(17)30405-X.

**32.**	Weiss HA, Buvé A, Robinson NJ, et al. The epidemiology of HSV-2 infection and its association with HIV infection in four urban African populations. *AIDS.* 2001;15:S97-S108. doi: 10.1097/00002030-200108004-00011.

**33.**	Mills E, Cooper C, Anema A, Guyatt G. Male circumcision for the prevention of heterosexually acquired HIV infection: a meta-analysis of randomized trials involving 11050 men. *HIV Medicine.* 2008;9(6):332-335. doi: 10.1111/j.1468-1293.2008.00596.x.

**34.**	Siegfried N, Muller M, Deeks JJ, Volmink J. Male circumcision for prevention of heterosexual acquisition of HIV in men. *Cochrane Database of Systematic Reviews.* 2009. doi: 10.1002/14651858.cd003362.pub2.

**35.**	Cohen MS, Chen YQ, McCauley M, et al. Antiretroviral therapy for the prevention of HIV-1 transmission. *N Engl J Med.* 2016;375(9):830-839. doi: 10.1056/NEJMoa1600693.

**36.**	Ministry of Health Benin. *Serology report by department, Benin.* 2015.

**37.**	Ministry of Health Benin. *Rapport de l’audit de la file active des personnes vivant avec le VIH au Bénin (Audit report).* 2017.

**38.**	Weller SC, Davis-Beaty K. Condom effectiveness in reducing heterosexual HIV transmission. *Cochrane Database of Systematic Reviews.* 2002. doi: 10.1002/14651858.cd003255.

**39.**	Glynn JR, Sonnenberg P, Nelson G, Bester A, Shearer S, Murray J. Survival from HIV-1 seroconversion in Southern Africa: a retrospective cohort study in nearly 2000 gold-miners over 10 years of follow-up. *AIDS.* 2007;21(5):625-632. doi: 10.1097/qad.0b013e328017f857.

**40.**	Lavreys L, Baeten JM, Chohan V, et al. Higher set point plasma viral load and more-severe acute HIV type 1 (HIV-1) illness predict mortality among high-risk HIV-1-infected African Women. *Clinical Infectious Diseases.* 2006;42(9):1333-1339. doi: 10.1086/503258.

**41.**	May M, Wood R, Myer L, et al. CD4+T cell count decreases by ethnicity among untreated patients with HIV infection in South Africa and Switzerland. *The Journal of Infectious Diseases.* 2009;200(11):1729-1735. doi: 10.1086/648096.

**42.**	Morgan D, Mahe C, Mayanja B, Okongo JM, Lubega R, Whitworth JAG. HIV-1 infection in rural Africa: is there a difference in median time to AIDS and survival compared with that in industrialized countries? *AIDS.* 2002;16(4):597-603. doi: 10.1097/00002030-200203080-00011.

**43.**	Pantazis N, Morrison C, Amornkul PN, et al. Differences in HIV natural history among African and non-African seroconverters in Europe and seroconverters in Sub-Saharan Africa. *PLoS ONE.* 2012;7(3):e32369. doi: 10.1371/journal.pone.0032369.

**44.**	Peters PJ, Karita E, Kayitenkore K, et al. HIV-infected Rwandan women have a high frequency of long-term survival. *AIDS.* 2007;21(Suppl 6):S31-S37. doi: 10.1097/01.aids.0000299408.52399.e1.

**45.**	Todd J, Glynn JR, Marston M, et al. Time from HIV seroconversion to death: a collaborative analysis of eight studies in six low and middle-income countries before highly active antiretroviral therapy. *AIDS.* 2007;21(Suppl 6):S55-S63. doi: 10.1097/01.aids.0000299411.75269.e8.

**46.**	Van der Paal L, Shafer LA, Todd J, Mayanja BN, Whitworth JAG, Grosskurth H. HIV-1 disease progression and mortality before the introduction of highly active antiretroviral therapy in rural Uganda. *AIDS.* 2007;21(Suppl 6):S21-S29. doi: 10.1097/01.aids.0000299407.52399.05.

**47.**	Bakari M, Urassa W, Mhalu F, Biberfeld G, Pallangyo K, Sandström E. Slow progression of HIV-1 infection in a cohort of antiretroviral naïve hotel workers in Dar es Salaam, Tanzania as defined by their CD4 cell slopes. *Scandinavian Journal of Infectious Diseases.* 2008;40(5):407-413. doi: 10.1080/00365540701708285.

**48.**	Badri M, Lawn SD, Wood R. Short-term risk of AIDS or death in people infected with HIV-1 before antiretroviral therapy in South Africa: a longitudinal study. *The Lancet.* 2006;368(9543):1254-1259. doi: 10.1016/s0140-6736(06)69117-4.

**49.**	French N, Mujugira A, Nakiyingi J, Mulder D, Janoff EN, Gilks CF. Immunologic and clinical stages in HIV-1–infected Ugandan adults are comparable and provide no evidence of rapid progression but poor survival with advanced disease. *JAIDS Journal of Acquired Immune Deficiency Syndromes.* 1999;22(5):509. doi: 10.1097/00126334-199912150-00013.

**50.**	Lawn SD, Myer L, Orrell C, Bekker L-G, Wood R. Early mortality among adults accessing a community-based antiretroviral service in South Africa: implications for programme design. *AIDS.* 2005;19(18):2141-2148. doi: 10.1097/01.aids.0000194802.89540.e1.

**51.**	Schim van der Loeff MF, Jaffar S, Aveika AA, et al. Mortality of HIV-1, HIV-2 and HIV-1/HIV-2 dually infected patients in a clinic-based cohort in The Gambia. *AIDS.* 2002;16(13):1775-1783. doi: 10.1097/00002030-200209060-00010.

**52.**	Keiser O, Taffé P, Zwahlen M, et al. All cause mortality in the Swiss HIV cohort study from 1990 to 2001 in comparison with the Swiss population. *AIDS.* 2004;18(13):1835-1843. doi: 10.1097/00002030-200409030-00013.

**53.**	Batona G, Gagnon M-P, Simonyan DA, Guedou FA, Alary M. Understanding the intention to undergo regular HIV testing among female sex workers in Benin. *JAIDS Journal of Acquired Immune Deficiency Syndromes.* 2015;68:S206-S212. doi: 10.1097/qai.0000000000000452.

**54.**	Ekouevi DK, Balestre E, Ba-Gomis F-O, et al. Low retention of HIV-infected patients on antiretroviral therapy in 11 clinical centres in West Africa. *Tropical Medicine & International Health.* 2010;15:34-42. doi: 10.1111/j.1365-3156.2010.02505.x.

**55.**	Ministry of Health Benin. "File Active": anonymised database of current female sex workers on ART in Benin. Ministry of Health, Benin; 2017.

**56.**	UNAIDS. *Rapport de suivi de la declaration de politique sur le vih/sida au Benin* 2016.

**57.**	Giguère K, Béhanzin L, Guédou FA, et al. PrEP use among female sex workers: no evidence for risk compensation. *JAIDS Journal of Acquired Immune Deficiency Syndromes.* 2019;82(3):257-264.

**58.**	Mboup A, Béhanzin L, Guédou FA, et al. Early antiretroviral therapy and daily pre-exposure prophylaxis for HIV prevention among female sex workers in Cotonou, Benin: a prospective observational demonstration study. *Journal of the International AIDS Society.* 2018;21(11):e25208. doi: 10.1002/jia2.25208.

**59.**	Donnell D, Baeten JM, Bumpus NN, et al. HIV protective efficacy and correlates of tenofovir blood concentrations in a clinical trial of PrEP for HIV prevention. *JAIDS Journal of Acquired Immune Deficiency Syndromes.* 2014;66(3):340-348. doi: 10.1097/qai.0000000000000172.

**60.**	Marrazzo JM, Ramjee G, Richardson BA, et al. Tenofovir-based preexposure prophylaxis for HIV infection among African women. *New England Journal of Medicine.* 2015;372(6):509-518. doi: 10.1056/nejmoa1402269.

**61.**	Van Damme L, Corneli A, Ahmed K, et al. Preexposure prophylaxis for HIV infection among African women. *New England Journal of Medicine.* 2012;367(5):411-422. doi: 10.1056/nejmoa1202614.

**62.**	Dovonou Albert Comlan AAC, Attinsounon Cossi Angelo,Saké Kadidjatou, Adè Serge, Ahoui Séraphin, Degla Jivather, Tchégnonsi Tognon Francis, Zannou Djimon Marcel, Adè Gabriel, Houngbé Fabien. Clinical and immunological characteristics in HIV-infected patients at the treatment initiation at the University Hospital of Parakou (Benin). *Open Journal of Immunology.* 2017;7.
